# Clinical characteristics with inflammation profiling of Long-COVID and association with one-year recovery following hospitalisation in the UK: a prospective observational study

**DOI:** 10.1101/2021.12.13.21267471

**Authors:** Writing Group (on behalf of the PHOSP-COVID Collaborative Group), Rachael A Evans, Olivia C Leavy, Matthew Richardson, Omer Elneima, Hamish J C McAuley, Aarti Shikotra, Amisha Singapuri, Marco Sereno, Ruth M Saunders, Victoria Claire Harris, Raminder Aul, Paul Beirne, Charlotte E Bolton, Jeremy S Brown, Gourab Choudhury, Nawar Diar Bakerly, Nicholas Easom, Carlos Echevarria, Jonathan Fuld, Nicholas Hart, John R Hurst, Mark Jones, Dhruv Parekh, Paul Pfeffer, Najib M Rahman, Sarah Rowland-Jones, Ajay M Shah, Dan G Wootton, Trudie Chalder, Melanie J Davies, Anthony De Soyza, John R Geddes, William Greenhalf, Neil J Greening, Liam G Heaney, Simon Heller, Luke Howard, Joseph Jacob, R Gisli Jenkins, Janet M Lord, William D-C Man, Gerry P McCann, Stefan Neubauer, Peter J M Openshaw, Joanna Porter, Jennifer Quint, Matthew J Rowland, Janet T Scott, Malcolm G Semple, Sally J Singh, David Thomas, Mark Toshner, Keir Lewis, Ryan S Thwaites, Andrew Briggs, Annemarie B Docherty, Steven Kerr, Nazir I Lone, Aziz Sheikh, Mathew Thorpe, Bang Zheng, James D Chalmers, Ling-Pei Ho, Alex Horsley, Michael Marks, Krisnah Poinasamy, Betty Raman, Ewen M Harrison, Louise V Wain, Christopher E Brightling

**Author notes:** Correspondence to: Prof Christopher Brightling, Institute for Lung Health, Department of Respiratory Sciences, NIHR Leicester BRC, University of Leicester, Leicester, United Kingdom. Joint first and last authors and contributed equally. Details of the PHOSP-COVID Collaborative Group membership is provided as a supplementary file. **Funding** Jointly funded by UK Research and Innovation and National Institute of Health Research (grant references: MR/V027859/1 and COV0319). **Ethics Approval** Ethics Ref: 20/YH/0225.

## Abstract

**Background:** There are currently no effective pharmacological or non-pharmacological interventions for Long-COVID. To identify potential therapeutic targets, we focussed on previously described four recovery clusters five months after hospital discharge, their underlying inflammatory profiles and relationship with clinical outcomes at one year.

**Methods:** PHOSP-COVID is a prospective longitudinal cohort study, recruiting adults hospitalised with COVID-19 across the UK. Recovery was assessed using patient reported outcomes measures (PROMs), physical performance, and organ function at five-months and one-year after hospital discharge. Hierarchical logistic regression modelling was performed for patient-perceived recovery at one-year. Cluster analysis was performed using clustering large applications (CLARA) k-medoids approach using clinical outcomes at five-months. Inflammatory protein profiling from plasma at the five-month visit was performed.

**Findings:** 2320 participants have been assessed at five months after discharge and 807 participants have completed both five-month and one-year visits. Of these, 35·6% were female, mean age 58·7 (SD 12·5) years, and 27·8% received invasive mechanical ventilation (IMV). The proportion of patients reporting full recovery was unchanged between five months 501/1965 (25·5%) and one year 232/804 (28·9%). Factors associated with being less likely to report full recovery at one year were: female sex OR 0·68 (95% CI 0·46-0·99), obesity OR 0·50 (95%CI 0·34-0·74) and IMV OR 0·42 (95%CI 0·23-0·76).

Cluster analysis (n=1636) corroborated the previously reported four clusters: ‘very severe’, ‘severe’, ‘moderate/cognitive’, ‘mild’ relating to the severity of physical, mental health and cognitive impairments at five months in a larger sample. There was elevation of inflammatory mediators of tissue damage and repair in both the ’very severe’ and the ’moderate/cognitive’ clusters compared to the ’mild’ cluster including interleukin-6 which was elevated in both comparisons. Overall, there was a substantial deficit in median (IQR) EQ5D-5L utility index from pre-COVID (retrospective assessment) 0·88 (0·74-1·00), five months 0·74 (0·60-0·88) to one year: 0·74 (0·59-0·88), with minimal improvements across all outcome measures at one-year after discharge in the whole cohort and within each of the four clusters.

**Interpretation:** The sequelae of a hospital admission with COVID-19 remain substantial one year after discharge across a range of health domains with the minority in our cohort feeling fully recovered. Patient perceived health-related quality of life remains reduced at one year compared to pre-hospital admission. Systematic inflammation and obesity are potential treatable traits that warrant further investigation in clinical trials.

**Funding:** UKRI & NIHR

**Research in Context:** *Evidence before this study:* We systematically searched PubMed and Embase databases for large studies reporting one-year follow-up data for hospitalised COVID-19 patients published between January 1, 2021 and November 7, 2021, without language restrictions. Search terms related to COVID-19, hospitalisation and long-term follow-up were used. A large prospective cohort study from Wuhan, China (n = 1276) showed that 49% of patients reported at least one persistent symptom during a follow-up clinic visit at 12 months post COVID-19; no significant improvement in exercise capacity was observed between six- and 12-month visits. Another two large cohort studies in China (n = 2433) and Spain (n = 1950) with one-year follow-up data from telephone interviews showed that 45% and 81% of patients reported at least one residual COVID-19 symptom, respectively. However, no previous studies have compared the trajectories of COVID-19 recovery in patients classified by different clinical phenotypes, and there are no large studies investigating the relationship between systemic inflammation and ongoing health impairments post COVID-19.

*Added value of this study:* In a diverse population of adults post-hospital admission with COVID-19, our large UK prospective multi-centre study reports several novel findings: the minority felt fully recovered at one year with minimal recovery from five months across any health domain; female sex and obesity are associated with being less likely to feel fully recovered at one year; several inflammatory mediators were increased in individuals with the most severe physical, mental health, and cognitive impairments compared to individuals with milder ongoing impairments.

*Implications of all the available evidence:* Both pharmacological and non-pharmacological interventions are urgently needed to improve the ongoing burden following hospitalisation for COVID-19 both for individuals and healthcare systems; our findings support the use of a precision medicine approach with potential treatable traits of systemic inflammation and obesity.

## Introduction

To date (December, 2021), there have been over 260 million cases of SARS-CoV-2 reported worldwide,^1^ 10 million cases in the UK^2^ and over half a million patients in the UK admitted to hospital for COVID-19. We, and others, have shown that this sizeable population are at high risk of persisting health impairments six months after discharge associated with reduced physical function and health-related quality of life.^3–4^ It is essential to understand the longer-term trajectory of recovery to identify ongoing healthcare needs, and the required response by healthcare systems and policy makers for this already large and ever-increasing population.

Much remains unknown about the longer-term sequelae of COVID-19. In the largest cohort study to date from Wuhan, China, nearly half of patients experienced persistent symptoms 12 months post-discharge from hospital for COVID-19.^5^ Between six- and 12-months post-discharge, there was no change in six-minute walk distance, but some improvement in the results of pulmonary imaging.^5^

The mechanisms underlying long-term persistence of symptoms are currently unknown. A potential hypothesis is that the hyperinflammation associated with acute COVID-19 leads to a persistent inflammatory state post COVID-19, associated with dysregulated immunity and multiorgan dysfunction. Although multiple studies have highlighted elevated inflammatory markers, including IL-6, associated with severity of acute illness,^6, 7^ there are no large studies investigating the relationship between systemic inflammation and ongoing health impairments after COVID-19.

Currently, there are no effective treatments for Long-COVID/ post-COVID-19 condition. Long-COVID is defined by National Institute for Health and Care Excellence (NICE) as on-going symptoms beyond four to 12 weeks after COVID-19 and post-COVID-19 condition by the World Health Organisation as “occurs in individuals with a history of probable or confirmed SARS CoV-2 infection, usually 3 months from the onset of COVID-19 with symptoms and that last for at least 2 months and cannot be explained by an alternative diagnosis”).^8, 9^ Improved characterisation of this population with an emphasis on elucidating underlying mechanisms is needed to identify potential therapeutic targets. We previously described four clinical recovery clusters of patients: ‘very severe’, ‘severe’, ‘moderate/cognitive’, and ‘mild’ defined by severity of ongoing physical, mental health and cognitive impairment five months after a hospital admission with COVID-19. In this report, using the ongoing PHOSP-COVID longitudinal study cohort, we sought to determine patient-perceived recovery, the associated risk factors for failure to recover at one year after discharge, and to investigate the association between our previously described clusters and multiple inflammatory mediators using a proteomics panel. We report the trajectory of recovery at one year across different health domains and the differences in recovery trajectory by cluster.

## Methods

### Study design and participants

The PHOSP-COVID cohort recruitment strategy including eligibility criteria has been described previously.^3^ In brief, we recruited patients aged 18 years and older who were discharged from over 50 National Health Service (NHS) hospitals across the four UK nations following admission to a medical assessment unit or ward for confirmed or clinician-diagnosed COVID-19 before March 31, 2021. The current analysis involves participants who consented to attend two additional in-person research visits (Tier 2 – see protocol and Table S1) within one-year after discharge alongside routine clinical care.

Written informed consent was obtained from all study participants. The study was approved by the Leeds West Research Ethics Committee (20/YH/0225) and is registered on the ISRCTN Registry (ISRCTN10980107).

### Procedures

Participants were invited to attend research visits at two to seven months post-discharge (‘five-month visit’) and at 10 to 14 months (‘one-year visit’). Participants were also able to attend a one-year only visit if they were outside the time period for a five-month visit at the time of consent and were discharged before November 30, 2020. The core set of data variables collected at each visit and included in this study are listed in the Supplementary materials, Table S2. These included baseline demographics, information about disease severity and treatment during their hospital admission, as well as physical tests, patient reported outcome measures (PROMs), pulmonary physiology and blood test results obtained at their clinical and research visits (see Supplementary methods). Patients were also asked to complete the EQ5D-5L, Washington Group Short Set Functioning (WGSS) and Visual analogue scale for Breathlessness and Fatigue retrospectively to assess their perceived pre-COVID health (Supplementary Table S2). Plasma samples obtained at the five-month visit were analysed using the Olink Explore 384 Inflammation panel. Sample processing and assay details are provided in the Supplementary methods.

The primary outcome for this analysis was patient-perceived recovery assessed using a study-specific questionnaire and the question ‘Do you feel fully recovered?’ and participants could answer ‘Yes’, ‘No’, or ‘Not sure’. Other outcomes included symptoms since their COVID-19 hospital admission collected on the bespoke study-specific questionnaire, validated patient reported outcome questionnaires, and physiological measures (including physical performance and spirometry). See Supplementary Tables S1 and S2 for details of all variables included in this study.

### Statistical analysis

Continuous variables were presented as median and interquartile range (IQR) or mean and standard deviation (SD). Binary and categorical variables were presented as counts and percentages (by row or by column as indicated in table legends). Participants were stratified by patient-perceived recovery: Yes (recovered), Not sure or No (not recovered).

Missing data were reported within each variable and per category. Within visit, a chi-squared test was used to identify differences in proportions across multiple categories. For normally distributed and non-normally distributed continuous data, analysis of variance (ANOVA F-test) and Kruskal Wallis tests respectively, were used to test differences across categories. For paired data between the five-month and one-year visit, a McNemar’s Chi-squared test with continuity correction and a McNemar’s Chi-squared test were used for binary variables and variables with > 2 levels, respectively. For normally distributed and non-normally distributed continuous data, a paired t-test and a Wilcoxon signed-rank test were used, respectively.

As previously described,^3^ univariable and hierarchical multivariable logistic regression models (admission hospital included as random effect) were used to explore risk factors associated with patient-perceived recovery. Missing data was addressed using multiple imputation (10 datasets, 10 iterations, final models combined using Rubin’s Rules), with the outcome used in imputation models, but not itself imputed.

In order to evaluate any potential bias arising due to loss to follow-up in participants attending the one-year visit, we compared characteristics and patient-perceived recovery (measured at five-months) in attendees and non-attendees, restricted to participants discharged between February 01, 2020 and 30 June, 2020. This was because this group had completed at least 14 months of follow-up at the time of analysis (October 6, 2021), allowing an additional month for completing data entry. We also compared characteristics between those attending five-month, one-year and both visits to understand if there were any systematic differences between these groups.

In this larger cohort, we repeated our previous unsupervised cluster analysis^3^ of patient recovery measured using symptom questionnaires (PROMS), physical performance and cognitive assessment data (Questionnaires: Dyspnoea-12, Fatigue - the Functional Assessment of Chronic Illness therapy (FACIT), Generalised Anxiety Disorder (GAD-7), Patient Health Questionnaire (PHQ-9), Post-Traumatic Stress Disorder Checklist for DSM-5 (PCL-5), Short Physical Performance Battery (SPPB) and Montreal Cognitive Assessment (MoCA) as continuous variables) from the five-month visit (discharge dates Feb 2020 – March 2021) using the clustering large applications (CLARA) k-medoids approach.^10^ Scores were centred, normalised and transformed so higher burden of disease represented higher values. A Euclidean distance metric was used, and the optimal number of clusters chosen using a silhouette plot. Cluster membership was determined for each individual using five-month visit data. Characteristics at one-year and change in characteristics between five-months and 12-months, are presented as cluster-stratified tables. All tests were two-tailed and p values <0·05 were considered statistically significant. We did not adjust for multiple testing.

Plasma protein levels were compared between clusters using the mildest recovery cluster as baseline and using multinomial regression with age, body mass index (BMI) and number of comorbidities as covariates (see Supplementary Methods). Significance was defined as p <0·1 after False Discovery Rate adjustment for multiple testing.

We used R (version 3·6·3) with the finalfit, tidyverse, mice, cluster, ggplot2, ggalluvial, radiant, dabestr and recipes packages.

## Results

At the time of analysis October 6, 2021, 2320 participants (discharged from hospital March 7, 2020 to April 18, 2021) had attended a five-month visit (median [IQR] 5 [4-6] months post-discharge), 924 participants (discharged February 28, 2020 to November 28, 2020) had returned for a one-year visit (13 [12-13]) months post-discharge) and 807 participants had attended both visits (Figure 1). The individual and hospital admission characteristics including severity of acute illness between those with a five-month visit, one-year visit, and those who attended both visits were similar (Table 1) except for acute treatment with corticosteroids. 350/1063 (32.9%) of participants discharged between February 01, 2020 and June 30, 2020 had either not attended or not had data entered for a one-year visit at the time of analysis. Age, BMI, co-morbidities, the proportion requiring invasive mechanical ventilation and five-month recovery status were similar between those who had and had not attended (Tables S3a and S3b).

**Figure 1.**
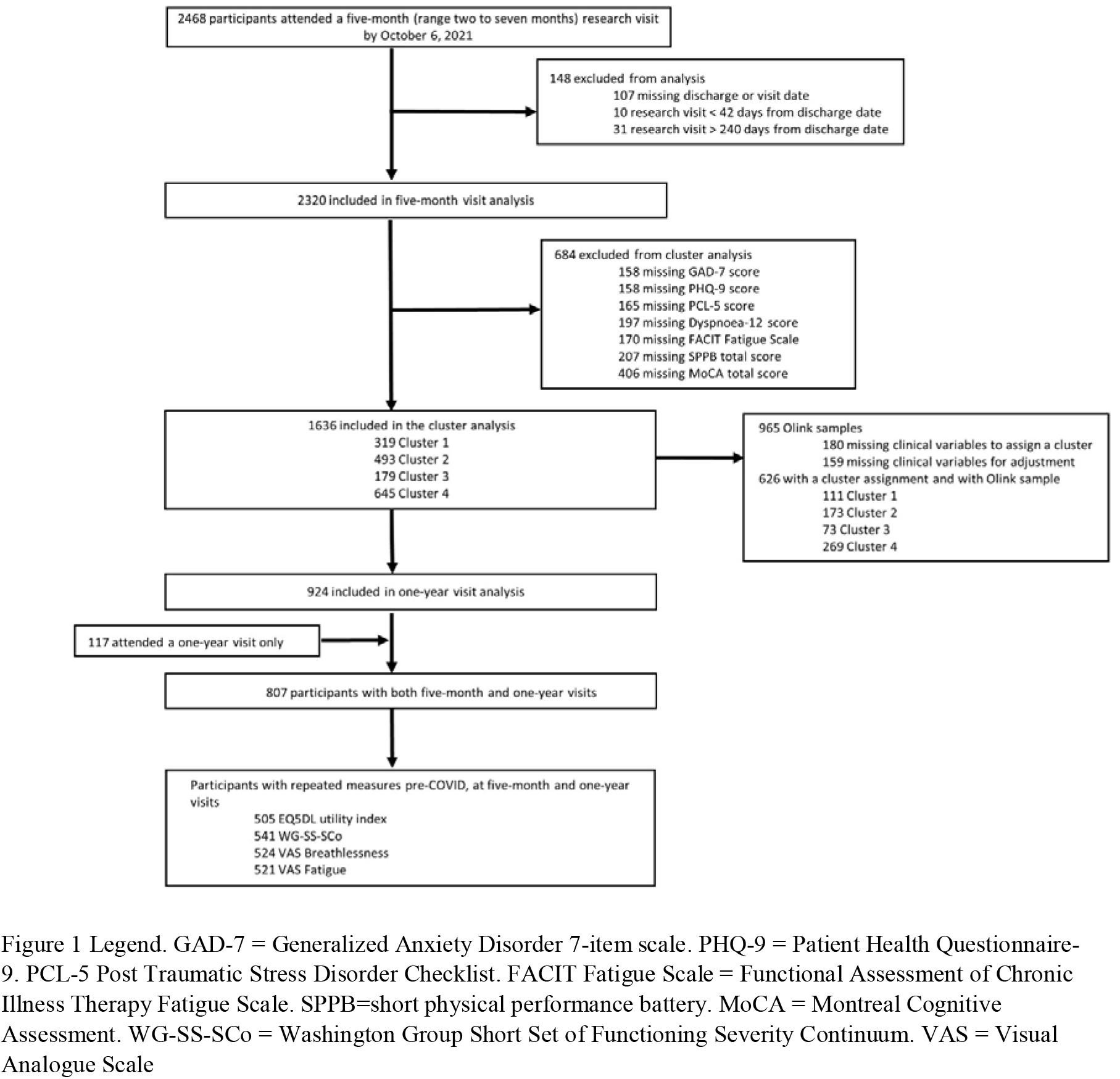
Consort diagram of participants

**Table 1.** Individual and hospital admission characteristics for the participants who had a five-month visit, a one-year visit, and both visits.

|  | Complete five-month visit | Complete one-year visit | Completed both visits |
| --- | --- | --- | --- |
| **Total N (%)** | 2320 (100.0) | 924 (100.0) | 807 (100.0) |
| **Age at admission, years\*** | 58.0 (12.6) | 58.9 (12.5) | 58.7 (12.5) |
| **Female Sex at birth¶** | 855 (39.0) | 319 (35.8) | 279 (35.6) |
| Missing data | 127 | 33 | 23 |
| **Ethnicity¶** |  |  |  |
| White | 1685 (75.4) | 681 (74.7) | 596 (74.5) |
| South Asian | 262 (11.7) | 102 (11.2) | 94 (11.8) |
| Black | 154 (6.9) | 68 (7.5) | 57 (7.1) |
| Mixed | 46 (2.1) | 19 (2.1) | 18 (2.2) |
| Other | 87 (3.9) | 42 (4.6) | 35 (4.4) |
| Missing data | 86 | 12 | 7 |
| **Pre-illness Occupation¶** |  |  |  |
| Full- or Part-time Employment | 1053 (55.5) | 493 (58.9) | 460 (59.7) |
| Other occupation status | 846 (44.5) | 344 (41.1) | 311 (40.3) |
| Missing data | 421 | 87 | 36 |
| **Health-care worker¶** | 344 (16.2) | 163 (19.0) | 153 (20.4) |
| Missing data | 203 | 67 | 58 |
| **Index of Multiple Deprivation (IMD)¶** |  |  |  |
| 1 (most deprived) | 517 (22.6) | 187 (20.4) | 163 (20.4) |
| 2 | 533 (23.3) | 186 (20.3) | 163 (20.4) |
| 3 | 404 (17.7) | 175 (19.1) | 155 (19.4) |
| 4 | 396 (17.3) | 160 (17.5) | 137 (17.2) |
| 5 (least deprived) | 438 (19.1) | 207 (22.6) | 180 (22.6) |
| Missing data | 32 | 9 | 9 |
| **BMI †** | 31.2 (27.7 - 36.1) | 31.5 (27.7 - 35.8) | 31.5 (27.7 - 35.7) |
| < 30 kg/m<sup>2</sup>¶ | 840 (41.1) | 349 (40.3) | 316 (41.2) |
| ≥ 30 kg/m<sup>2</sup>¶ | 1204 (58.9) | 517 (59.7) | 451 (58.8) |
| Missing data | 276 | 58 | 40 |
| **Smoking status¶** |  |  |  |
| Never | 1085 (54.7) | 429 (53.2) | 350 (52.8) |
| Ex-smoker | 864 (43.5) | 369 (45.7) | 301 (45.4) |
| Current smoker | 36 (1.8) | 9 (1.1) | 12 (1.8) |
| Missing data | 335 | 117 | 144 |
| **WHO clinical progression scale¶** |  |  |  |
| WHO – class 3-4 | 385 (16.9) | 171 (18.6) | 145 (18.0) |
| WHO – class 5 | 959 (42.2) | 342 (37.1) | 299 (37.1) |
| WHO – class 6 | 517 (22.7) | 167 (18.1) | 139 (17.2) |
| WHO – class 7-9 | 412 (18.1) | 241 (26.2) | 224 (27.8) |
| Missing data | 47 | < 5 | 0 |
| **Comorbidities¶** |  |  |  |
| Median number of comorbidities † | 2.0 (0.0 to 3.0) | 2.0 (0.0 to 3.0) | 2.0 (0.0 to 3.0) |
| **0** | 642 (27.7) | 251 (27.2) | 213 (26.4) |
| **1** | 468 (20.2) | 172 (18.6) | 154 (19.1) |
| **≥2** | 1210 (52.2) | 501 (54.2) | 440 (54.5) |
| **Cardiovascular** | 1027 (44.3) | 425 (46.0) | 375 (46.5) |
| **Renal and endocrine** | 250 (10.8) | 100 (10.8) | 89 (11.0) |
| **Respiratory** | 611 (26.3) | 235 (25.4) | 215 (26.6) |
| **Type 2 diabetes** | 451 (19.4) | 179 (19.4) | 163 (20.2) |
| **Neuro-psychiatric** | 465 (20.0) | 171 (18.5) | 149 (18.5) |
| **Admission duration, days** | 13.9 (18.2) | 17.0 (24.7) | 17.8 (22.1) |
| **Positive SARS-CoV-2 PCR¶** | 1916 (92.4) | 796 (90.8) | 700 (90.6) |
| **Missing data** | 246 | 47 | 34 |
| **Systemic steroids¶** | 1173 (54.2) | 251 (29.8) | 226 (30.2) |
| **Missing data** | 157 | 81 | 59 |
| **Antibiotic therapy¶** | 1769 (79.5) | 746 (84.5) | 660 (84.5) |
| **Missing data** | 96 | 41 | 26 |
| **Therapeutic dose anti-coagulation¶§** | 941 (43.4) | 320 (37.5) | 282 (37.3) |
| **Missing data** | 154 | 71 | 51 |

¶ = Number (%) with positive response, \*Mean [SD], †Median [IQR]. Percentages are calculated by category after exclusion of missing data for that variable. N= Number. WHO classes are as follows: 3–4 = no continuous supplemental oxygen needed; 5 = continuous supplemental oxygen only; 6 = continuous or bi-level positive airway pressure ventilation or high-flow nasal oxygen; and 7–9 = invasive mechanical ventilation or other organ support. BMI = body-mass index. PCR= Polymerase Chain Reaction. §Therapeutic dose anticoagulation; does not include intermediate doses which were not recorded.

Overall, 25·5% (501/1965) of patients felt fully recovered at five-months post-discharge, and 28·9% (232/804) at one year (Figure 2a, Table S10b) with similar proportions observed in those with paired data (Table 2b). Assuming all the participants who had missing data at one year had either not recovered or fully recovered, the adjusted proportion of participants feeling fully recovered at one year is 212/1063 (19·9%) or 604/1063 (60·2%), respectively (Table 3b). The proportions that reported feeling fully recovered excluding or including those not yet due a one-year visit and missing data are shown in Figure S2a and b, respectively. In multivariable analysis, female sex (OR 0·68 (0·46-0·99)), Body Mass Index (BMI) 30 kg/m^2^ or greater (OR 0·50 (0·34-0·74)) and receiving invasive mechanical ventilation (IMV) World Health Organisation [WHO] category 7-9 (OR 0·42 (0·23-0·76)) were all independent factors associated with being less likely to recover at one-year (Figure 2b and Table S4). There was no effect of receiving systemic corticosteroids (OR 1·05 (0·66-1·65)) during the acute admission on patient-perceived recovery at one-year for the whole cohort (Figure 2b, Table S4). There was also no effect of time from discharge to the research visit (OR 1·00 (1·00-1·01)).

**Figure 2.**
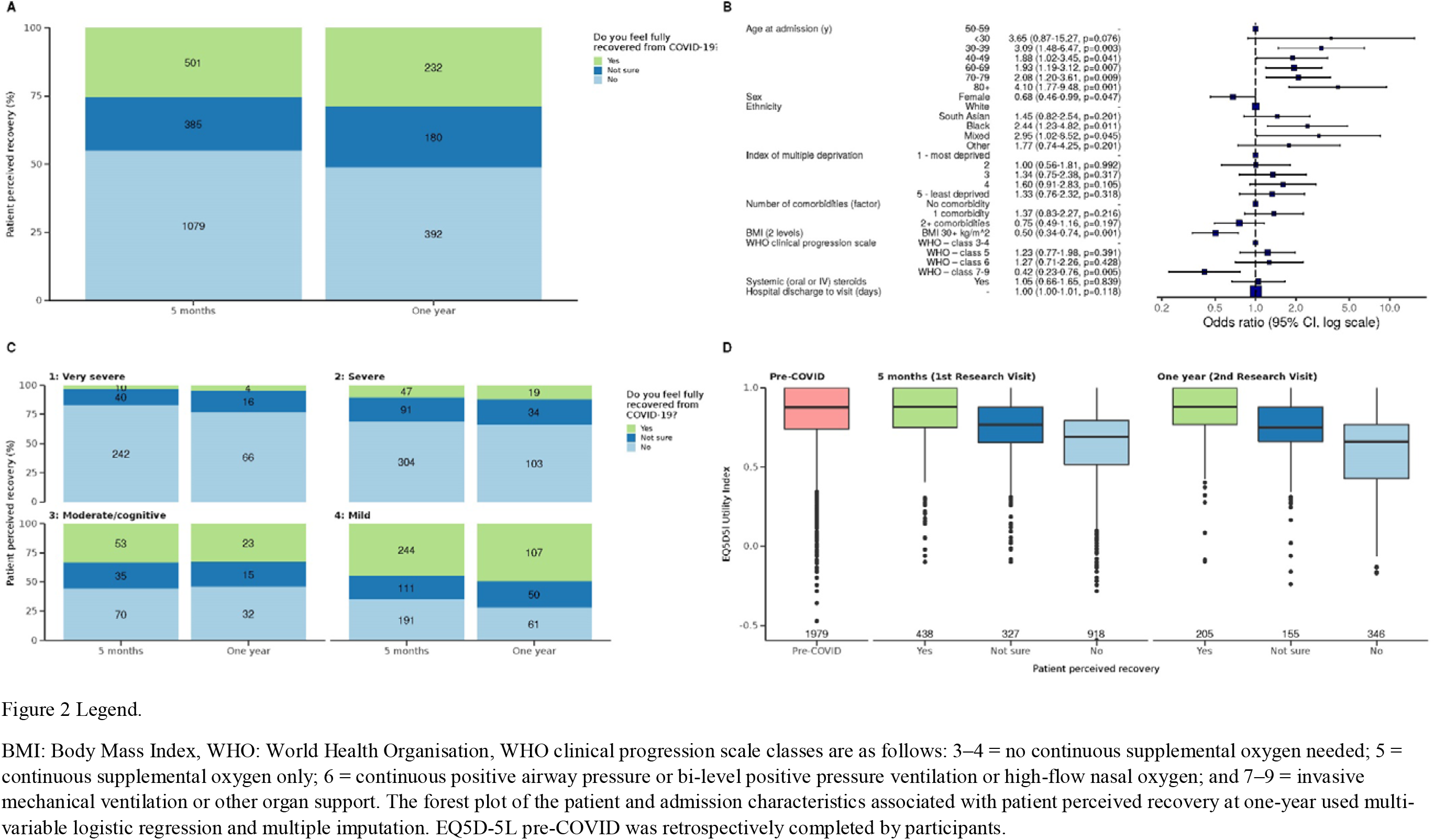
Four panel plot demonstrating patients perceived recovery at one year a) compared with five months, b) risk factors for being less likely to recover, c) compared by the four clusters d) compared to health-related quality of life (assessed by the EQ5D-5L Utility Index)

**Table 2.** A comparison of the change between five-months and one-year post-discharge in patient reported outcome measures including mental health, physical function, cognitive impairment, and organ function

|  | Five-month visit |  |  |  |  | One-year visit |  |  |  |  |
| --- | --- | --- | --- | --- | --- | --- | --- | --- | --- | --- |
|  | Total N (5-months) | Recovered | Not sure | Not recovered | p | Total N (1-year) | Recovered | Not sure | Not recovered | p |
| Total N (%) All participants | 1965 | 501 (25.5) | 385 (19.6) | 1079 (54.9) | - | 804 | 232 (28.9) | 180 (22.4) | 392 (48.8) | - |
| Total N (%) Paired data | 590 | 151 (25.6) | 113 (19.2) | 326 (55.3) | - | 590 | 168 (28.5) | 138 (23.4) | 284 (48.1) | - |

|  | Five-month visit – stratified by patient perceived recovery at five months, n=1965 |  |  |  | Paired data at five-month and one-year visits, n=807 |  |  |  |
| --- | --- | --- | --- | --- | --- | --- | --- | --- |
|  | Recovered | Not sure | Not recovered | P | N pairs with available data | Five-months | One-year | p |
| **Total n (%)** | 501 (25.5) | 385 (19.6) | 1079 (54.9) | N/A | **% recovered**<br>n=590 | 151 (25.6) | 168 (28.5) | 0.12 |
| **Time to review from discharge, days †** | 166 (127- 191) | 165 (122 - 191) | 157 (119 - 189) | 0.040 | 807 | 178 (156 - 197) | 384 (359-409) | <0.0001 |
| **BMI †** | 29.4 (26.6-33.5) | 31.5 (28.0-36.3) | 31.6 (28.0-36.4) | <0.0001 | 602 | 30.7 (27.3-35.0) | 31.1 (27.5-35.5) | <0.0001 |
| **< 30 kg/m<sup>2</sup>** | 230 (54.5) | 131 (40.2) | 360 (38.8) | <0.0001 | 602 | 275 (45.7) | 255 (42.4) | 0.021 |
| **≥ 30 kg/m<sup>2</sup>** | 192 (45.5) | 195 (59.8) | 568 (61.2) |  |  | 327 (54.3) | 347 (57.6) |  |
| **Any symptom at time point¶** | 415 (83.0) | 369 (95.8) | 1066 (99.0) | <0.0001 | 619 | 580 (93.7) | 584 (94.3) | 0.67 |
| **Median number of symptoms †** | 3 (1 - 7) | 8 (4 - 15) | 14 (8 - 20) | <0.0001 | 619 | 9 (4 - 16) | 9 (4 - 17) | 0.010 |
| **Fatigue VAS †** | 0.0 (0.0 - 2.0) | 2.0 (0.0 - 5.0) | 5.0 (2.0 - 8.0) | <0.0001 | 521 | 3.0 (0.0 - 6.0) | 3.0 (0.0 - 6.0) | 0.090 |
| **Breathlessness VAS †** | 0.0 (0.0 - 1.0) | 1.5 (0.0 - 4.0) | 4.0 (1.0 - 6.0) | <0.0001 | 524 | 2.0 (0.0 - 5.0) | 2.0 (0.0 - 5.0) | 0.052 |
| **Anxiety (GAD7 >8)¶** | 53 (11.3) | 81 (22.6) | 339 (33.4) | <0.0001 | 684 | 164 (24.0) | 147 (21.5) | 0.13 |
| **Depression (PHQ-9 ≥10)¶** | 47 (9.9) | 96 (26.7) | 426 (42.0) | <0.0001 | 680 | 181 (26.6) | 169 (24.9) | 0.25 |
| **PTSD (PCL-5 ≥38)¶** | 18 (3.8) | 34 (9.5) | 202 (20.0) | <0.0001 | 680 | 83 (12.2) | 68 (10.0) | 0.055 |
| **Dyspnoea-12 score\*** | 2.1 (4.8) | 5.1 (7.1) | 8.9 (8.8) | <0.0001 | 702 | 6.0 (8.1) | 5.5 (7.7) | 0.040 |
| **FACIT fatigue subscale score\*** | 43.6 (8.8) | 36.5 (11.2) | 29.1 (12.8) | <0.0001 | 679 | 35.7 (12.9) | 36.3 (12.5) | 0.070 |
| **BPI severity\*** | 9.2 (10.1) | 12.0 (9.5) | 15.2 (10.0) | <0.0001 | 455 | 12.9 (9.7) | 12.8 (9.8) | 0.90 |
| **BPI interference\*** | 11.2 (16.4) | 17.6 (17.3) | 24.8 (19.8) | <0.0001 | 432 | 19.6 (19.1) | 19.2 (17.8) | 0.53 |
| **SPPB $\leq 10$¶** | 181 (38.9) | 166 (46.0) | 582 (58.8) | <0.0001 | 685 | 318 (46.4) | 309 (45.1) | 0.53 |
| **ISWT distance, m\*** | 487.6 (274.7) | 431.4 (242.3) | 384.6 (249.4) | <0.0001 | 509 | 453.6 (262.8) | 468.2 (267.8) | 0.017 |
| **ISWT % predicted\*** | 63.5 (30.7) | 57.8 (28.1) | 52.5 (28.7) | <0.0001 | 429 | 60.1 (29.4) | 61.2 (28.7) | 0.22 |
| **Rockwood CFS $\geq 5$¶** | 9 (2.0) | 17 (4.9) | 81 (8.2) | <0.0001 | 651 | 41 (6.3) | 46 (7.1) | 0.53 |
| **MoCA <23¶** | 66 (16.5) | 39 (12.4) | 147 (15.8) | 0.26 | 623 | 89 (14.3) | 62 (10.0) | 0.0013 |
| **MoCA (adjusted) <23¶** | 57 (14.3) | 36 (11.4) | 125 (13.4) | 0.52 | 623 | 72 (11.6) | 55 (8.8) | 0.034 |
| **FEV1 <80% predicted¶** | 43 (21.4) | 43 (27.7) | 131 (27.9) | 0.20 | 287 | 67 (23.3) | 63 (22.0) | 0.64 |
| **FVC <80% predicted¶** | 40 (19.9) | 33 (21.4) | 155 (33.2) | 0.00030 | 281 | 79 (28.1) | 63 (22.4) | 0.018 |
| **FEV1/FVC <70%¶** | 25 (8.8) | 23 (10.6) | 49 (7.5) | 0.35 | 311 | 25 (8.0) | 28 (9.0) | 0.61 |
| **BNP $\geq 100$ ng/L or Pr-NT-BNP $\geq 400$ ng/L¶** | 27 (8.7) | 24 (10.3) | 35 (5.2) | 0.014 | 335 | 30 (9.0) | 29 (8.7) | 1.0 |
| **HbA1C $\geq 6.0\%$ (DCCT/NGSP)¶** | 6.1 (1.2) | 6.2 (1.3) | 6.2 (1.3) | 0.60 | 399 | 140 (35.1) | 130 (32.6) | 0.21 |
| **eGFR < 60 ml/min/1.73 m<sup>2</sup>¶** | 49 (12.0) | 35 (11.3) | 101 (11.4) | 0.94 | 564 | 73 (12.9) | 79 (14.0) | 0.45 |
| **CRP >5 mg/L¶** | 83 (20.9) | 75 (23.9) | 239 (27.1) | 0.052 | 557 | 126 (22.6) | 133 (23.9) | 0.52 |
| **Fibrinogen (g/L)\*** | 3.5 (0.9) | 3.6 (0.8) | 3.6 (0.9) | 0.34 | 379 | 3.5 (0.9) | 3.5 (0.9) | 0.97 |
| **Ferritin (µg/L)\*** | 136.3 (149.7) | 145.3 (150.6) | 140.1 (166.4) | 0.79 | 449 | 141.2 (150.6) | 129.1 (136.2) | <0.0001 |
| **EQ5DL Utility Index †** | 0.88 (0.75 - 1.00) | 0.77 (0.65 - 0.88) | 0.69 (0.52 - 0.80) | <0.0001 | 585 | 0.7 (0.6 - 0.9) | 0.8 (0.6 - 0.9) | 0.95 |
| **EQ5D-5L VAS †** | 85.0 (72.2 - 91.2) | 75.0 (60.0 - 85.0) | 70.0 (50.0 - 80.0) | <0.0001 | 586 | 75.0 (60.0 - 90.0) | 75.0 (60.0 - 90.0) | 0.43 |
| **WG-SS-SCo †** | 0.0 (0.0 - 2.0) | 2.0 (0.5 - 3.0) | 3.0 (1.0 - 8.0) | <0.0001 | 548 | 2.0 (0.0 - 4.0) | 2.0 (0.0 - 4.0) | 0.73 |

For the current five-month dataset with complete data n=1636 (Figure 1), the previously identified four clusters^3^ were confirmed representing ‘very severe’ physical and mental health impairment (n=319), ‘severe’ physical and mental health impairment (n=493), ‘moderate/cognitive’ physical health impairment with cognitive impairment (n=179) and ‘mild’ mental and physical health impairment (n=645) (Table S5, Figure S2). 86·7% (664/766) of individuals included in the previous study^3^ were re-assigned to the same recovery cluster as before; the cognitive cluster had the most assignment alterations (60/127). Characteristics of individuals in each recovery cluster are shown in Table S6. The very severe cluster had a higher proportion of female sex (165/306, 53·9%) and obesity (204/288, 70·8%) compared to the ‘mild’ cluster (177/624, 28·4%) and (288/568, 50·2%), respectively.

After quality control, plasma proteome data for 296 protein features and complete clinical data for cluster assignment were available at five-months for 626 participants: ‘very severe’ cluster n=111, ‘severe’ cluster n=173, ‘moderate/cognitive’ cluster n=73 and ‘mild’ cluster n=269. Age, BMI, and two or more co-morbidities were associated with cluster membership whereas receiving IMV during the acute illness was not (analysis in participants with plasma proteome data and a cluster assignment) (Table S7). After adjustment for age, BMI, and co-morbidity count, 13 proteins were significantly elevated in participants in the ‘very severe’ recovery cluster compared to those in the ‘mild’ cluster (Table S8, Figure 3). These were Trefoil Factor 2 (TFF2), Transforming Growth Factor Alpha (TGFA), Lysosomal Associated Membrane Protein 3 (LAMP3), CD83 molecule (CD83), Galectin-9 (LGALS9), Plasminogen Activator, Urokinase Receptor (PLAUR), Interleukin 6 (IL6), Erythropoietin (EPO), Fms Related Receptor Tyrosine Kinase 3 Ligand (FLT3LG), Agrin (AGRN), Secretoglobin Family 3A Member 2 (SCGB3A2), Follistatin (FST) and C-Type Lectin Domain Family 4 Member D (CLEC4D) (Figure S4). In addition, IL6 and CD70 molecule (CD70) were significantly increased in the ‘moderate/cognitive’ recovery cluster compared to the ‘mild’ cluster.

**Figure 3.**
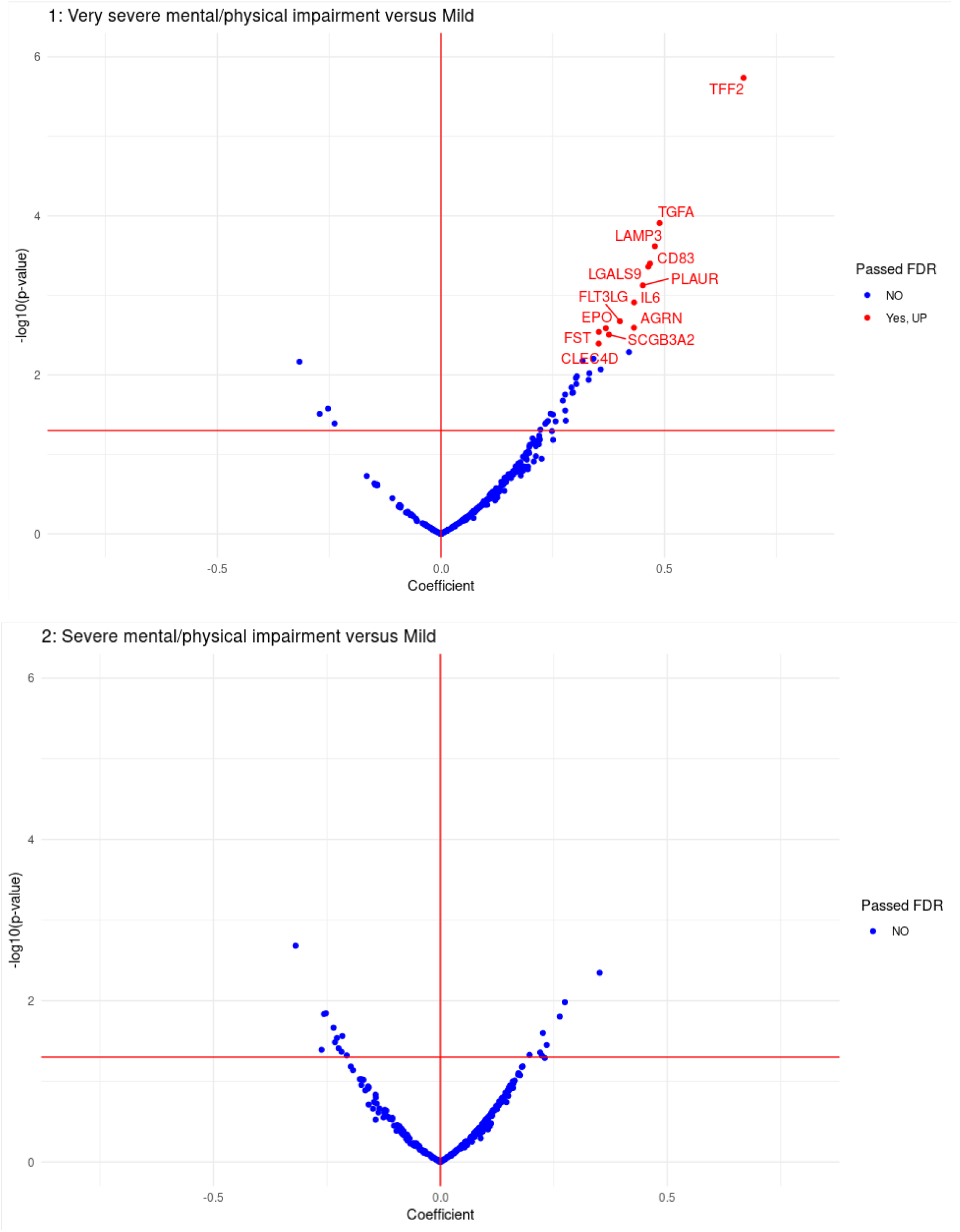
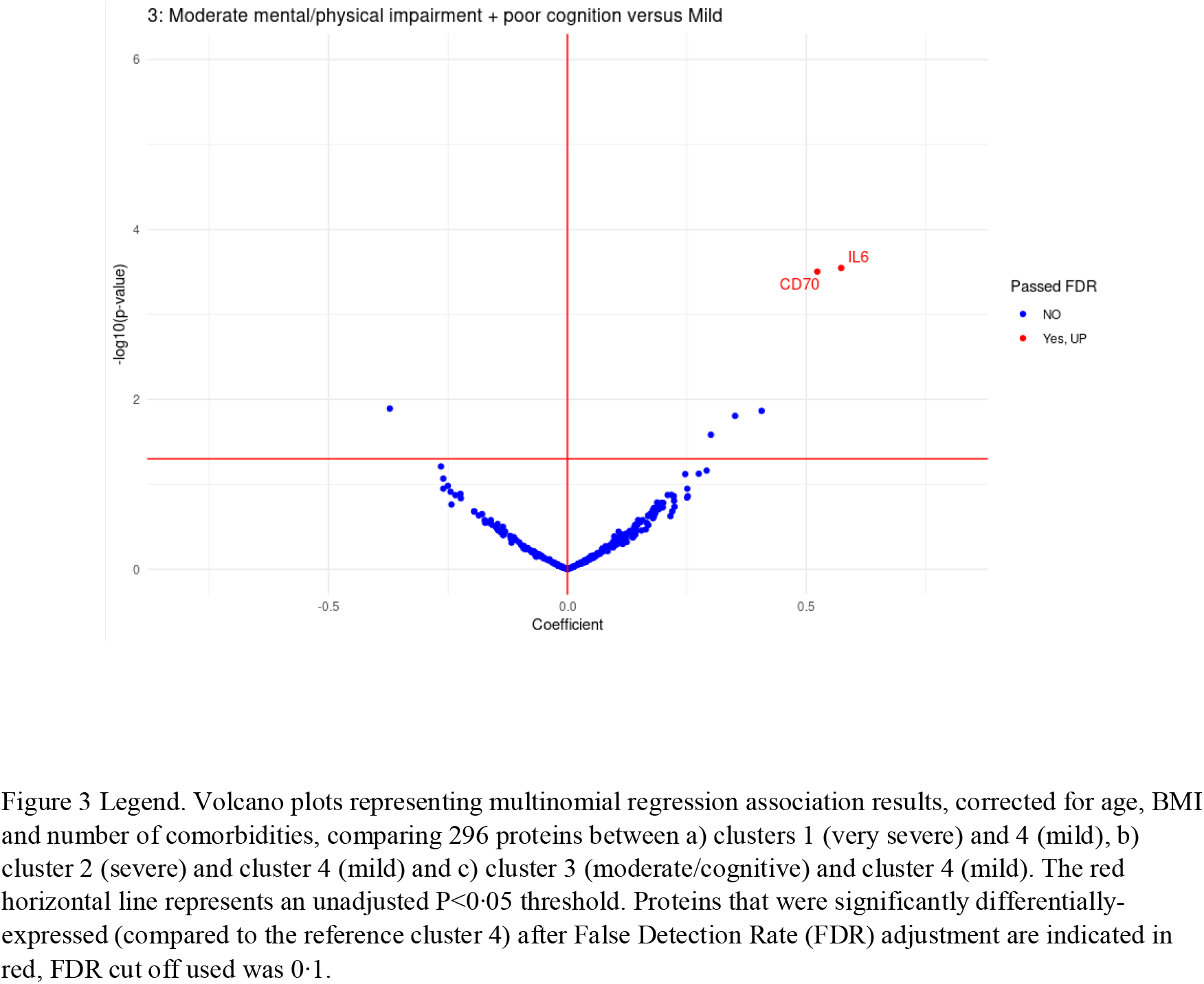
Volcano plots representing multinomial regression association results for comparison of 368 proteins between a) clusters 1 (very severe) and 4 (mild), b) clusters 2 (severe) and 4 (mild) and c) clusters 3 (moderate/cognitive) and 4 (mild). The red horizontal line represents an unadjusted P<0·05 threshold. Proteins that were significantly over-expressed (compared to the reference cluster 4) after FDR adjustment are indicated in red and those significantly under-expressed are indicated in black, FDR cut off used was 0·1.

**Figure 4.**
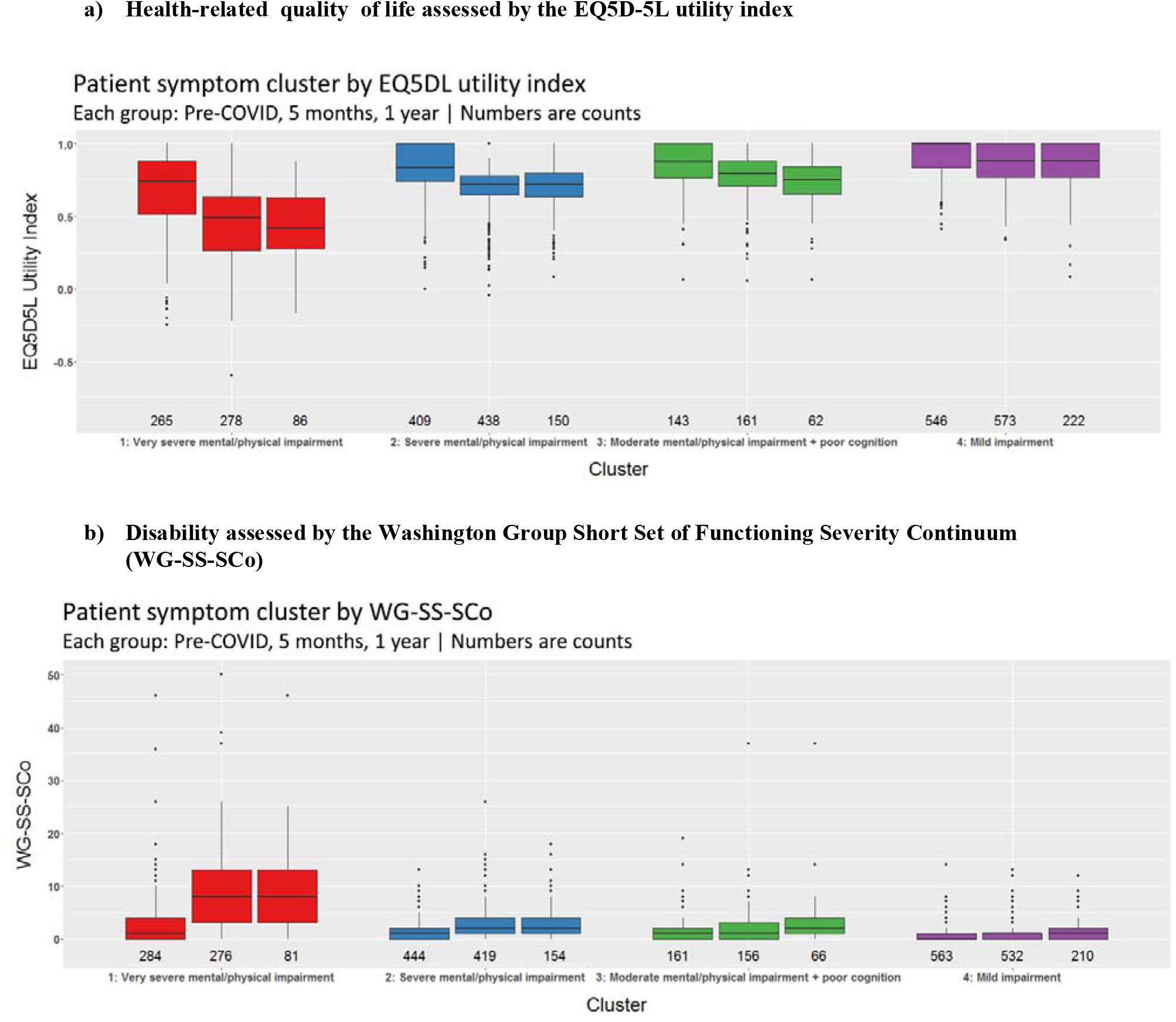
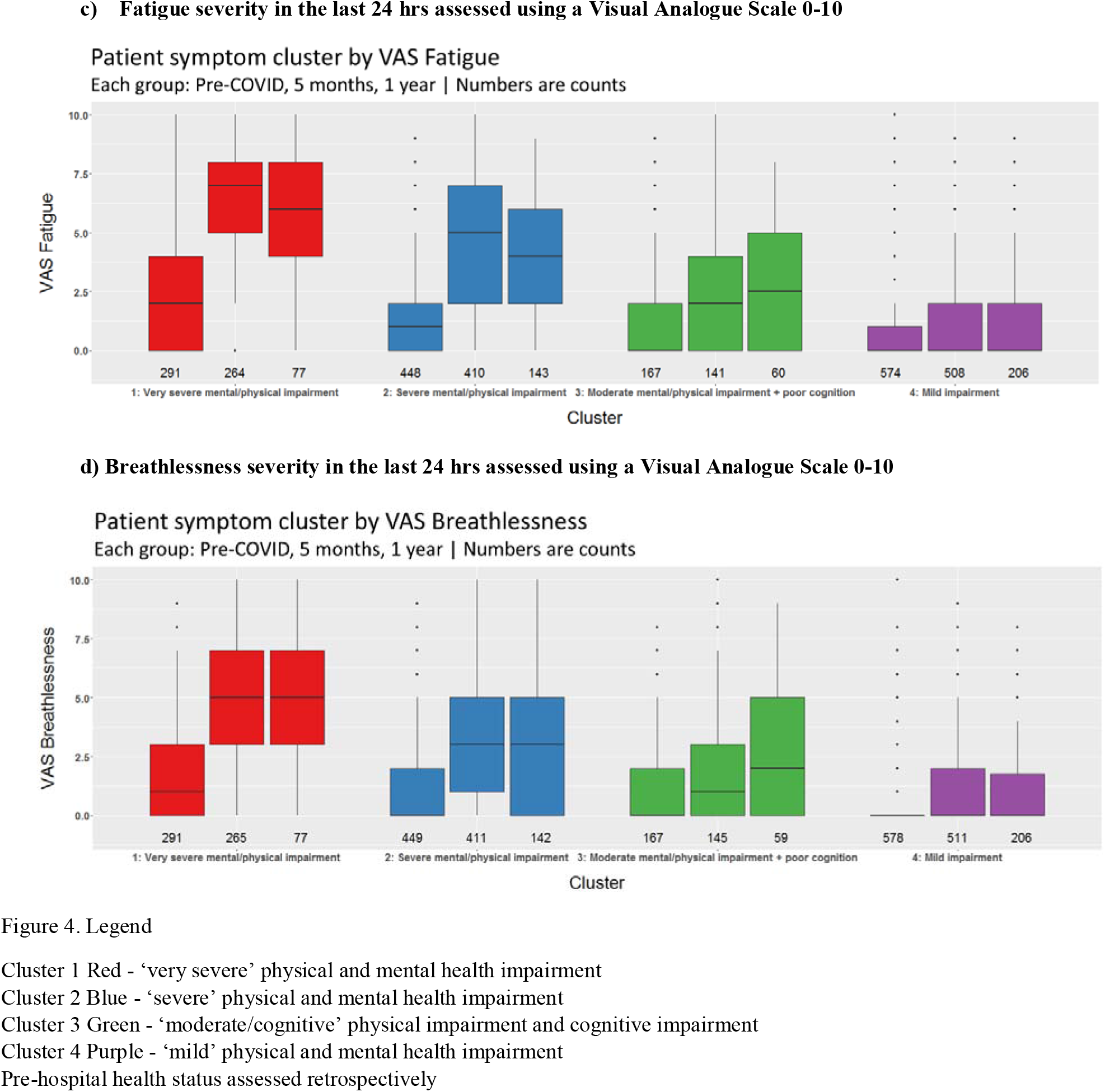
A comparison between the four ‘severity’ cluster phenotypes of health-related quality of life, disability, fatigue and breathlessness across pre-hospitalisation, five-months and one-year after hospital discharge.

The top 10 most common persistent symptoms at one-year post-discharge were fatigue (463/770, 60·1%), aching muscles (442/809, 54·6%), physical slowing down (429/811, 52·9%), poor sleep (402/769, 52·3%), breathlessness (395/769, 51·4%), joint pain or swelling (382/803, 47·6%), slowing down in thinking (377/808, 46·7%), pain (359/770, 46·6%), short term memory loss (360/808, 44·6%) and limb weakness (341/813, 41·9%) (Table S9). Overall, these were essentially unchanged in prevalence from five months to one year, with small reductions in rates of limb weakness (47·6% at five-months vs. 41·7% at one-year, p=0·010), paraesthesia (40·6% vs. 35·2%, p=0·014) and balance problems (34·9% vs. 30·0, p=0·008). There was either no or minimal improvement in PROMS, physical function, cognitive impairment, or organ function at one-year compared to five-months post-discharge (paired data Table 2b and presented by patient-perceived recovery in Tables S10a and b). At one year, 147/684 (21·5%) and 169/680 (24·9%) participants had clinically relevant symptoms of anxiety and/or depression, respectively, 68/680 (10·0%) had symptoms compatible with post-traumatic stress disorder, and 55/623 (8·8%) had significant cognitive impairment (Table 2b). Measures of symptoms and physical function were significantly different across participants feeling fully recovered, not sure, or not fully recovered at five months and one-year but cognitive impairment and measures of organ function (except for Forced Vital Capacity [FVC]) were not (Table 2b). Similarly, health-related quality of life was significantly different across participants reporting being fully recovered, not sure or not recovered at both five months, and one year (Figure 2d and Tables S10a and b).

In addition to higher proportions of female sex and obesity (Table S6), the more severe clusters were associated with lower proportion of feeling fully recovered 11/272 (4·0%) vs. 90/179 (50·3%) (Figure 2c), reduced exercise capacity ISWT 44·4% predicted vs. 72·4% predicted, higher number of symptoms 20 vs 4, and higher proportion of elevated CRP level >5mg/L 38·4% vs. 14·5% compared to the mild cluster (Table 3, Figure S3). A comparison of health outcomes across the four clusters between the five-month and one-year time-points (n=602) shows there was minimal change across the two time-points for the four clusters (Table 3). In the ‘very severe’ cluster, symptoms of anxiety, depression, breathlessness and fatigue significantly improved between five-months and one-year, but with minimal change in physical performance and no overall change in systemic inflammation measured by CRP levels. Cognitive impairment significantly improved at one-year in the ‘moderate/cognitive’ cluster and showed a non-significant trend towards improvement in the ‘severe’ cluster but was unchanged in the other clusters (Table 3). Compared with patient perceived pre-COVID-19 health, decrements were seen at five months and sustained at one-year across health-related quality of life (EQ5D-5L), disability (WG-SS-SCo), and severity of breathlessness and fatigue experienced in the last 24 hrs (Figure 3, Table S11, Figure S5).

**Table 3.**
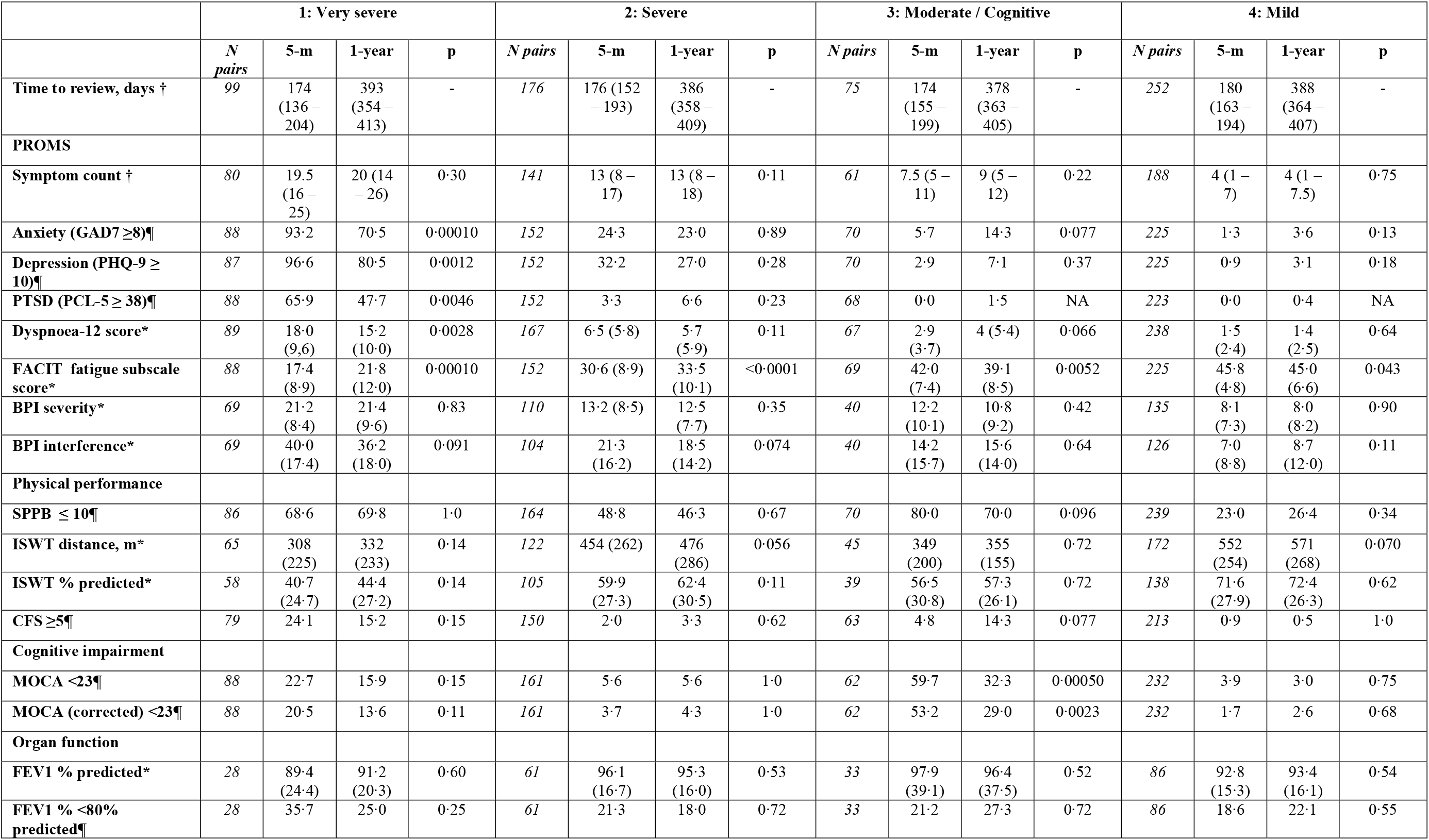
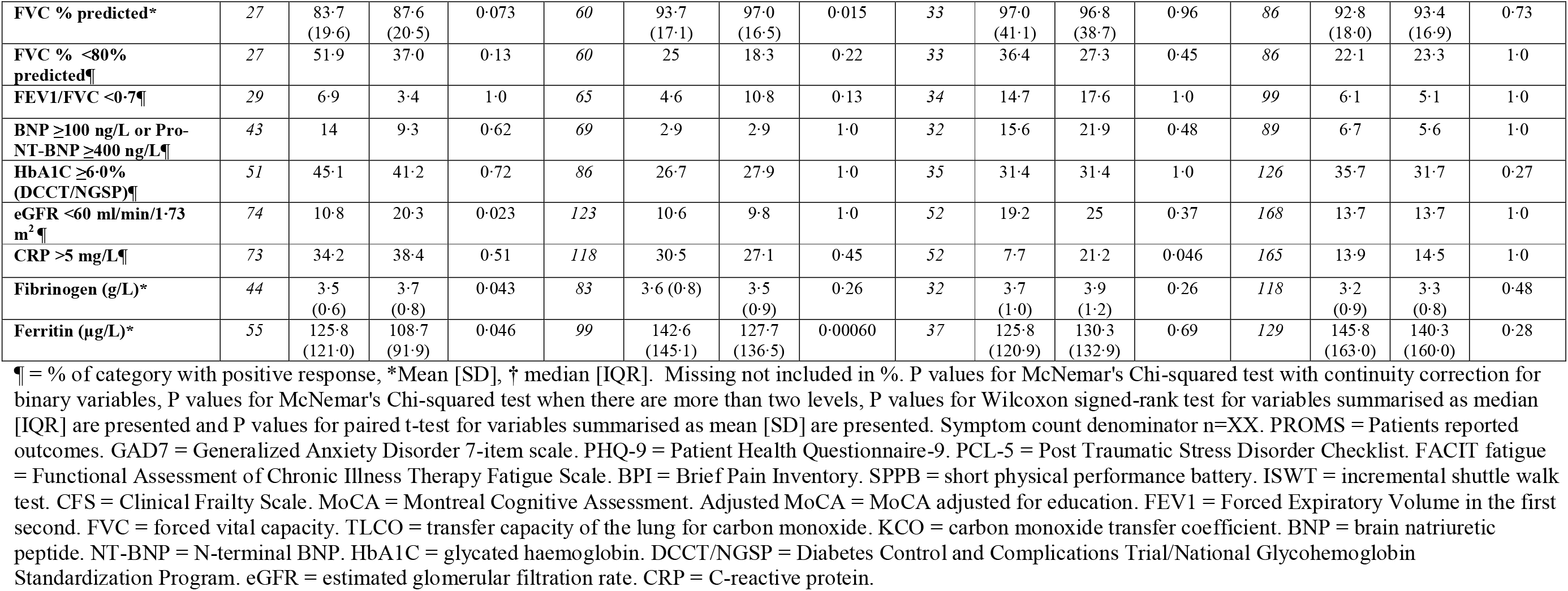
A comparison of the change in patient reported outcome measures including mental health, physical function, cognitive impairment, and organ function between five-month and one-year sub-grouped by the four ‘severity’ cluster phenotypes

## Discussion

In a diverse UK population of adult survivors of COVID-19, we found the minority of participants felt fully recovered one year after hospital discharge with minimal improvement after their five-month assessment. The most common ongoing symptoms were fatigue, muscle pain, physically slowing down, poor sleep and breathlessness. The major risk factors for failure to recover at one-year were female sex, obesity and receiving IMV during the acute illness. There were substantial impairments in health-related quality of life (HRQoL) at five-months and one-year compared to retrospective self-reported pre-infection levels. Cluster analysis using the participants’ five-month assessment corroborated four different clusters: ‘very severe’, ‘severe’, ‘moderate/cognitive’, and ‘mild’ based on the severity of physical, mental and cognitive impairments with similar characteristics to previously reported.^3^ We confirmed that obesity, reduced exercise capacity, a higher number of symptoms and elevated serum CRP level were associated with the more severe clusters.^3^ In the largest post-hospital cohort with systemic inflammatory profiling to date, inflammatory mediators consistent with persistent lung and systemic inflammation were elevated between both the ‘very severe’ and ‘moderate/cognitive’ clusters, and the ‘mild’ cluster. We therefore highlight traits to identify individuals at high risk of non-recovery and potential targetable pathways for interventions.

Comparing the systemic inflammatory profiling at five months after discharge between the ‘very severe’ and ‘mild’ cluster, the most elevated protein, TFF2, is a protein released with mucin from mucosal epithelium including lung and gastric mucosa. It has postulated roles in repair of damaged epithelium^11^ and in combination with IFN-kappa reduced duration of infection in a small open-label randomised controlled trial of acute COVID-19.^11^ In a previous study of patients during acute illness with COVID-19 using Olink proteomics, IL-6 was the most upregulated protein at day seven amongst patients who developed ARDS and subsequently died.^6^ Similarly, other proteins we identified such as LAMP3, Gal-9, CD83 are involved in T-cell macrophage and dendritic cell activation and were associated with increased morbidity and mortality during acute COVID-19 infection.^12–14^ These changes suggest persistent mucosal epithelial abnormalities and inflammatory cell activation. Elevated serum levels of the C-terminal fragment of Agrin have been reported in older adults with sarcopenia, possibly related to break down of the neuromuscular junction.^15^ The raised Agrin levels seen here may therefore contribute to the high prevalence of physical impairment.

Interestingly, in the ‘moderate/cognitive’ cluster versus the ‘mild’ cluster IL6 and CD70 were elevated suggesting possible neuroinflammation contributing to the cognitive impairment as CD70 has been implicated in inflammation in the Central Nervous System (CNS)^16^ via a role in differentiation of proinflammatory pathogenic lymphocytes. There were small improvements in cognition in the ‘moderate/cognitive’ cluster and a trend towards improvement in the ‘very severe’ cluster indicating that some of this deficit was not pre-existing and potentially modifiable; however, considerable deficit persisted at one year. The associations with the inflammatory mediators remained after adjusting for age, BMI and co-morbidities, and the proportion having received invasive mechanical ventilation was similar across the clusters; all factors known to be associated with systemic inflammation.^17^ Taken together, the elevated mediators provide biological plausibility for the persistent severe impairments seen in physical, mental health and cognitive impairment after COVID-19.

The limited recovery from five-months to one-year post-hospitalisation in our study across symptoms, mental health, exercise capacity, organ impairment and quality-of-life is striking. There are limited similar detailed prospective longitudinal studies for patients hospitalised with COVID-19 but in a larger cohort we confirm their findings of minimal recovery.^18–2^

Although the large-scale study from Wuhan, China suggests a greater magnitude of recovery, new onset symptoms persisted in half of the patients.^5^ Notably, the Wuhan cohort included less severe acute illness with only 1% requiring IMV and 7% requiring high flow nasal oxygen and/or continuous positive airway pressure (CPAP), had fewer pre-existing co-morbidities, and a higher proportion of never-smokers. In non-COVID related ARDS survivors, little recovery in health-related quality of life is observed beyond six months, but larger improvements in walking distance^21, 22^ than we report post-COVID-19 in our cohort where over 70% did not receive IMV. In non-hospitalised patients after COVID-19 the proportion that develop Long-COVID appears lower.^23, 24^

The responses for patient perceived recovery were discriminatory across all the PROMS and exercise measures providing additional validity for this outcome measure. We found female sex and obesity were major risk factors for not recovering at one year confirming existing results from smaller cohorts^25^, and non-hospitalised cohorts.^26–28^ Female sex was similarly associated with worse recovery for fatigue, mental health and lung function at 12 months in the Wuhan cohort.^5^ In our clusters, female sex and obesity were also associated with more severe ongoing health impairments including reduced exercise performance and HRQoL at one year, potentially highlighting a group which may need higher intensity interventions such as supervised rehabilitation. Health-related quality of life pre-COVID was substantially greater than at five-months post-discharge across all four clusters indicating that the persistent burden of impaired physical and mental health is not simply explained by pre-existing morbidity. The total number and range of ongoing symptoms at one year was striking, positively associated with the severity of Long-COVID, and emphasises the multi-system nature of Long-COVID. Other studies have shown the number of symptoms during the acute illness were associated with the likelihood of developing Long-COVID.^29^ Whether the number of ongoing symptoms, a simple widely available measure, could underpin a future risk score deserves further attention. We suggest that female sex and obesity should be considered when planning follow-up healthcare services.

Currently, there are no specific therapeutics for Long-COVID and our data highlights effective interventions are urgently required. Critically, our findings of persistent systemic inflammation, particularly in those in the ‘very severe’ and ‘moderate/cognitive’ clusters, suggests these groups might respond to anti-inflammatory strategies. The upregulation of IL6 suggests that anti-IL6 biologics that were successful for acute-hospitalised COVID-19^30^ might also have a place in Long-COVID. Similarly, activation of the uPAR pathway suggests that IL1 activation might play a role, with soluble uPAR a biomarker in acute COVID-19 associated with good response to the recombinant IL-1 receptor antagonist anakinra.^31^ Impaired exercise capacity was also associated with the more severe clusters and showed minimal improvement at one-year (well below the minimum clinically important distance for other long-term conditions).^32–34^ Current therapies available for some adults with Long-COVID include rehabilitation,^35^ but the optimal exercise prescription is contentious due to concerns of post-exertional symptom exacerbation (PESE). Our data confirm high prevalence of musculoskeletal symptoms including muscle ache, fatigue, breathlessness, physically slowing down, and limb weakness.^5, 16^ This supports the need to investigate rehabilitation in combination with other therapies to improve skeletal muscle function such as mitochondrial energetics, mitophagy enhancers and drugs to combat cell senescence (associated with ageing).

The concordance of the severity of physical and mental health impairment in Long-COVID highlights the need for close integration between physical and mental healthcare for Long-COVID including assessment and interventions, but also for healthcare professional knowledge transfer to improve patient care. It also suggests the need for complex interventions that target both physical and mental health impairments in order to ameliorate symptoms. However, specific therapeutic approaches to manage post-traumatic stress disorder may be needed.^36^ With obesity being associated with both non-recovery and severity of Long-COVID, whether weight reduction using pharmacological and non-pharmacological approaches in concert can ameliorate Long-COVID warrants further investigation. Beyond diet and lifestyle interventions, glucagon like peptide-1 (GLP-1) analogues have recently been reported to achieve clinically important weight reduction.^37^

Our cohort study is ongoing, and we report these one-year findings to add novel findings to the limited literature, help direct clinical care and further investigation. However, there are limitations. There will be selection bias for participants returning for a one-year visit, although we have not found overt differences between the demographics, or five-month recovery status between attendees and non-attendees of the one-year visit. Notwithstanding this limitation, even with the assumption of all participants with missing data having fully recovered then the highest estimate is 60% of participants feeling fully recovered at one year demonstrating a substantial proportion with ongoing new morbidity. Our cohort has a higher proportion of patients requiring IMV than typically seen in UK hospitals^38^ and therefore our results may not be directly generalisable to the wider population. To reduce uncertainty of the impact of pre-existing illness, we asked our recruits whether they felt fully recovered i.e., back to their normal. We also asked them retrospectively to estimate their pre-COVID-19 health status including the most prevalent symptoms, disability and health-related quality of life; we recognise there might be recall bias. Data linkage to electronic patient records is in process, but not currently available so in the current report pre-existing co-morbidities were self-reported and data regarding hospital admissions and mortality in the first year are unavailable. Our study suggests that persistent inflammation may be underlying ongoing impairment in some participants; the specific mechanisms underlying this signal require further investigation and replication. We describe several associations with more severe health impairments at one year. Our findings cannot confirm causality but suggest these associations should be further investigated as part of mechanistic studies and clinical trials.

In summary, our study highlights an urgent need for healthcare services to support this large and rapidly increasing patient population where, in our cohort, there is a substantial burden of symptoms, reduced exercise capacity and large decrements in health-related quality of life after one year. Without effective treatments, Long-COVID has the potential to become a highly prevalent new long-term condition. Our study also provides a rationale for investigating treatments for Long-COVID with a precision medicine approach to target treatments to the relevant phenotype, potentially anti-inflammatories, weight reduction and rehabilitation to restore health-related quality of life.

## Contributors

The manuscript was initially drafted by CEB, RAE, LVW further developed by the writing committee. CEB, RAE, LVW, OE, HJCM, AShi, ASi, MJD, ABD, NIL, AShe, JDC, L-PH, AH, MM, KP, BR, made substantial contributions to the conception and design of the work. RAE, ASi, MS, RMS, VCH, RA, PB, CEBo, JSB, GC, NDB, NE, CE, JF, NH, JRH, MJ, DP, PP, NMR, SR-J, AMS, DGW, JDC, L-PH, AH, MM, WD-CM made substantial contributions to the acquisition of data. CEB, RAE, LVW, OCL, MR, OE, HJCM, MS, TC, MJD, ADS, JRG, WG, NJG, LGH, SH, LH, JJ, RGJ, JML, WD-CM, GPM, SN, PJMO, JP, JQ, MJR, JTS, MGS, SJS, MTo, KL, RST, AB, ABD, SK, NIL, AShe, MTh, BZ, JDC, L-PH, AH, MM, KP, BR, EMH, made contributions to the analysis, or interpretation of data for the work. All authors contributed to data interpretation and critical review and revision of the manuscript. Final approval of the version to be published and agreement to be accountable for all aspects of the work in ensuring that questions related to the accuracy or integrity of any part of the work are appropriately investigated and resolved.

## Supporting information

Manuscript Supplement

PHOSP-COVID Collaborative Group

## Data Availability

All data produced in the present work are contained in the manuscript

## Acknowledgments

This study would not be possible without all the participants who have given their time and support. We thank all the participants and their families. We thank the many research administrators, health-care and social-care professionals who contributed to setting up and delivering the study at all of the 69 NHS Trusts and 25 Research Institutions across the UK as well as all the supporting staff at the at the NIHR Clinical Research Network, Health Research Authority, Research Ethics Committee, Department of Health and Social Care Public Health Scotland, Public Health England and support from the ISARIC Coronavirus Clinical Characterisation Consortium (ISARIC4C). At the NIHR Office for Clinical Research Infrastructure (NOCRI) we thank Kate Holmes (for her support in coordinating the charities group) Ivana Poparic and Peter Sargent and Sheuli Porkess at the Association of the British Pharmaceutical Industry for their advice in commercial discussions. We are very grateful to all the charities that have provided insight to the study-Action Pulmonary Fibrosis, Alzheimer’s Research UK, Asthma UK /British Lung Foundation UK/BLF, British Heart Foundation, Diabetes UK, Cystic Fibrosis Trust, Kidney Research UK, MQ Mental Health, Muscular Dystrophy UK, Stroke Association Blood Cancer UK, McPin Foundations, Versus Arthritis. We thank the NIHR Leicester Biomedical Research Centre patient and public involvement group and the Long Covid Support Group.

## Data sharing

The protocol, consent form, definition and derivation of clinical characteristics and outcomes, training materials, regulatory documents, requests for data access and other relevant study materials are available online at www.phosp.org.

## Ethics Approval

Ethics Ref: 20/YH/0225 Trial ID: ISRCTN10980107

## Supplement Tables and Figures

Table S1. Tier 2 outcome measures.

Table S2. Methods and thresholds for processing of variables and outcome measures used in the current analysis.

Table S3. Comparison of individual and admission characteristics, and patient-perceived recovery between the one-year visit attendees and non-attendees for participants discharged between February 2020 – June 2020.

Table S4. Comparison between imputed and non-imputed logistic regression of predictors of failure to recover at 1-year (multi-variable and multi-level).

Table S5. Cluster medoids and characteristics

Table S6. Comparison of participant demographics, clinical characteristics, and admission characteristics stratified by the four clinical recovery clusters.

Table S7: Multinomial logistic regression of clinical characteristics against cluster membership (mild cluster as reference) for participants with plasma proteomic data.

Table S8. Comparison of plasma proteomics between a) cluster 1 (very severe) and cluster 4 (mild), b) cluster 2 (severe) and cluster 4 (mild) and c) cluster 3 (moderate/cognitive) and cluster 4 (mild). Multinomial regression results with unadjusted P<0·05 are presented.

Table S9. Ongoing symptoms recorded at five-months and one-year post-discharge from hospital

Table S10. Patient reported outcomes, physiological and biochemical tests stratified by patient perceived recovery outcomes at five-months and one-year visits a) paired data and b) unpaired data.

Table S11. A comparison between the four clinical recovery phenotypes of health-related quality of life, disability, fatigue and breathlessness across pre-hospitalisation, and five-months and one-year post-discharge

Figure S1. A comparison of participant-perceived recovery between five months and one-year

Figure S2. Clusters of mental, cognitive, and physical health impairments at five-months Figure S3. Characteristics associated with the four ‘recovery’ clusters

Figure S4. Estimation plots for the features significantly upregulated when comparing cluster 1 (very severe) to cluster 4 (mild) (panels a-m) and when comparing cluster 3 (moderate/cognitive) to cluster 4 (mild) (panels n and o).

Figure S5. Health-related quality of life, disability, and symptoms across the four ‘recovery’ clusters assessed for pre-hospitalisation (patient estimated), and at five-months and one-year post-discharge (individual complete data for all three time-points.

