## Supplementary material for "Clinical characteristics with inflammation profiling of Long-COVID and association with one-year recovery following hospitalisation in the UK: a prospective observational study": Manuscript Supplement

Supplemental Methods

**Olink Explore 384 Inflammation panel analysis**

*PHOSP-COVID biological sampling*

PHOSP-COVID recruitment and biological sampling is ongoing. All tier 2 participants were invited to provide a blood sample and consent for biological sample analysis at both their five and one-year research visits. Pre-labelled blood, urine and oral sample kits were provided to all sites from the PHOSP-COVID team at the Liverpool Good Clinical Practice Laboratory (Liverpool GCP Lab Facility, 1st Floor William Henry Duncan Building, 6 West Derby Street, Liverpool, L7 8TX). All sites followed the same laboratory protocol for obtaining, processing and locally storing samples following collection. Processed samples were then shipped back to the Liverpool GCP Laboratory at intervals where they were logged and stored using a customised Matrix Laboratory Information Management system (Autoscribe, Reading).

*Plasma sample processing*

Blood samples were obtained at the first visit for all participants who provided consent for biological sampling. After sampling, the 10ml K2EDTA tubes were inverted 8 to 10 times and then centrifuged at 1500 x g for 10 minutes at room temperature. Plasma supernatant was aliquoted and immediately frozen at –80C at site prior to shipping. Samples were shipped by courier on dry ice to Liverpool GCP Laboratory.

*Sample prioritisation for Olink assay*

Tier 2 participants were considered eligible for Olink assay if they were in one or more of the following ordered priority groups (participants could be in more than one group):

1) Participants included in the clustering analysis in Evans et al. (n=767)

2) Participants who were included in the total 1077 participants^1^ and had also been included in the ISARIC4C Immunology study [UK CRN /CPMS ID 14152 IRAS ID126600] (n=78)

3) Participants who had already been followed up by the ISARIC4C Immunology study but were not within the 1077 participants in Evans et al. (n=142)

4) Participants within the 1077 participants in Evans et al and not already included in groups 1 to 3 above.

5) Participants who had completed a first visit but were not included in the groups above.

Participant samples were ranked according to the priorities above and participants for which plasma aliquots from the first visit 10ml K2EDTA blood sample had been received and logged at the Liverpool GCP Laboratory before end of May 2021 and for which there were no LIMS quality issues identified were selected for Olink assay. A total of 975 unique samples and 24 duplicates were plated with duplicate sample pairs plated across different plates. The latest sample collection date was 13^th^ April 2021.

Four participants withdrew consent during sample picking and processing and were excluded from further analysis. Six samples were subject to kit queries and were excluded from this analysis leaving 965 participants with plasma proteomic data available for further analysis.

*Olink proteomic assay*

Samples were assayed using the Olink(R) Explore 384 Inflammation panel at the Olink laboratory in Uppsala. In total, 364978 out of 376464 (97%) sample-feature assays passed Olink quality control. Samples were provided to Olink on 96 well plates randomised according to site and date of sample collection. Analysis was carried out blind to any patient details (including time points and site). Each plate had at least 4 samples that were duplicates of a patient (and time point) on another plate so that a plate had at least 4 other plates carrying duplicate pairs and any plate could be linked to any other via duplicates. Analysis was carried out blind to the duplicates to ensure inter-plate standardisation was robust.

*Data analysis.*

For this study, only participants included in the cluster analysis (Table 3) were included in the Olink data analysis. Of 368 unique features included in the assay, 296 passed quality control. Features were required to be above the logarithm of the odds (LOD) in 80% of samples, to be above the minimum proportion of samples passing QC which was set at 80% and to have a global assay interquartile range of greater than 0·1. Multivariable regression was undertaken comparing clusters 1 (very severe), 2 (severe) and 3 (moderate/cognitive) with cluster 4 (mild) including age, sex, BMI, ethnicity, number of comorbidities, and WHO severity at admission. Age, BMI and number of comorbidities (categorised as zero, one or two or more) were significantly associated with cluster membership for at least one comparison (P<0·001, Table S7a) and were included as adjustments in a multinomial logistic regression for each feature comparing clusters 1 to 3 with cluster 4 (mild). Prior to fitting the Multinomial model, the expression values for the 296 features that passed quality control were standardised (for each feature subtract mean and divide by standard deviation). The Multinomial regression was applied on a feature-by-feature basis, with cluster as the dependent variable. A list comprising the p-values for each feature was formed and adjusted p-values were obtained from this list using False Discovery Rate (FDR), FDR cut off used was 0·1. Analysis of the Olink data was performed using R 4·0·0. Multinomial logistic regression was performed using the Radiant package, version 1·4·1. Estimation plots were produced using the dabestr package,version 0·3·0 and included protein expression levels for all participants with available data (not limited to those with complete clinical data for adjustment for age, BMI and number of comorbidities). This research used the SPECTRE High Performance Computing Facility at the University of Leicester.

Supplement Tables and Figures

Table S1. Tier 2 outcome measures.

Table S2. Methods and thresholds for processing of variables and outcome measures used in the current analysis.

Table S3. Comparison of individual and admission characteristics, and patient-perceived recovery between the one-year visit attendees and non-attendees for participants discharged between February 2020 – June 2020.

Table S4. Comparison between imputed and non-imputed logistic regression of predictors of failure to recover at 1-year (multi-variable and multi-level).

Table S5. Cluster medoids and characteristics

Table S6. Comparison of participant demographics, clinical characteristics, and admission characteristics stratified by the four clinical recovery clusters.

Table S7. Multinomial logistic regression of clinical characteristics against cluster membership (mild cluster as reference) for participants with plasma proteomic data.

Table S8. Comparison of plasma proteomics between a) cluster 1 (very severe) and cluster 4 (mild), b) cluster 2 (severe) and cluster 4 (mild) and c) cluster 3 (moderate/cognitive) and cluster 4 (mild). Multinomial regression results with unadjusted P<0·05 are presented.

Table S9. Ongoing symptoms recorded at five-months and one-year post-discharge from hospital

Table S10. Patient reported outcomes, physiological and biochemical tests stratified by patient perceived recovery outcomes at five-months and one-year visits a) paired data and b) unpaired data.

Table S11. A comparison between the four clinical recovery phenotypes of health-related quality of life, disability, fatigue and breathlessness across pre-hospitalisation, and five-months and one-year post-discharge

Figure S1. A comparison of participant-perceived recovery between five months and one-year

Figure S2. Clusters of mental, cognitive, and physical health impairments at five-months

Figure S3. Characteristics associated with the four ‘recovery’ clusters

Figure S4. Estimation plots for the features significantly upregulated when comparing cluster 1 (very severe) to cluster 4 (mild) (panels a-m) and when comparing cluster 3 (moderate/cognitive) to cluster 4 (mild) (panels n and o).

Figure S5. Health-related quality of life, disability, and symptoms across the four ‘recovery’ clusters assessed for pre-hospitalisation (patient estimated), and at five-months and one-year post-discharge (individual complete data for all three time-points)

**Table S1. Tier 2 outcome measures.**

| **Module** | **Tier 2 outcome measures reported in the current analysis** | **Other Tier 2 outcome measures not analysed** |
| --- | --- | --- |
| **Symptoms** | Patient symptom questionnaire (PHOSP-COVID study specific questionnaire)  Dyspnoea12 Questionnaire  The Functional Assessment of Chronic Illness Therapy (FACIT)  Brief Pain Inventory Questionnaire (BPI) | MRC dyspnoea scale grade  Nottingham activities of daily living Questionnaire |
| **Health-related Quality of life and Disability** | Euroqol EQ5D-5L  Washington Short Set of Functioning |  |
| **Respiratory** | Pulmonary Function Tests Including: Spirometry (FEV_1_, FVC) and Transfer Factor (TLCO, KCO) |  |
| **Cardiac** | Blood tests: BNP / NT-Pro-BNP | ECG Image Collection  Blood tests: Troponin I /Troponin T & Lipid Profile |
| **Renal** | Blood tests: eGFR | Urine tests: Albumin:Creatinine Ratio, Protein:Creatinine Ratio, Bedside urinalysis |
| **Pre-diabetes/diabetes** | Blood tests: HbA1C |  |
| **Haematological** | Blood tests: D Dimer, Ferritin | Blood tests: Full Blood Count, INR |
| **Systemic inflammation** | Blood test: CRP | Blood tests: Fibrinogen |
| **Other organ function** |  | Blood tests: Liver function tests, 25-Hydroxyvitamin D, Bone Profile |
| **Physical performance** | Incremental Shuttle Walk Test (ISWT) to assess exercise capacity  Short Physical Performance Battery (SPPB) | Daily physical activity by wearable technology (Geneactive)  Handgrip Strength  General Practice Physical Activity Questionnaire (GPPAQ) |
| **Frailty** | Rockwood Clinical Frailty Scale (CFS) | Fried’s frailty definition  SARC-F Questionnaire |
| **Body composition** | Body Mass Index (BMI) calculation from Height and Weight Measurement | Body composition estimation via: Bio-Electrical Impedance Analysis (BIA) or Duel Energy X-ray Analysis (DXA)  Waist Circumference Measurement |
| **Mental Health** | Generalised Anxiety Disorder Questionnaire (GAD-7)  Patient Health Questionnaire (PHQ-9)  Post-Traumatic Stress Disorder Checklist for DSM-5 Questionnaire (PCL-5) |  |
| **Cognition** | Montreal Cognitive Assessment (MoCA) |  |

FEV_1_ = Forced Expiratory Volume in 1 second, FVC = Forced Vital Capacity, TLCO = Transfer Capacity of the Lung for Carbon Monoxide, KCO = carbon monoxide transfer coefficient, BNP = Brain Natriuretic Peptide or NT-BNP N-Terminal Brain Natriuretic Peptide, ECG = electrocardiogram, HbA1C = glycosylated haemoglobin, INR = international normalized ratio, eGFR = estimated Glomerular Filtration Rate, CRP = C - reactive protein

**Table S2. Methods and thresholds for processing of variables and outcome measures used in the current analysis.**

|  | **Method** |
| --- | --- |
| **Table 1** |  |
| Indices of Multiple Deprivation | Obtained using postcode.^2^ |
| Comorbidities | A pre-existing comorbidity was considered absent if not indicated by a ‘yes’ on the case report form. |
| Admission duration | Calculated using the hospital discharge date and the earliest admission date to the same or different hospital for the participant’s COVID-19 episode. |
| **Table 2 and 3** |  |
| Symptoms at five months and one year | The total number of current symptoms reported were from the following list which were answered as binary Yes/No questions: Aching in your muscles (pain), Physical slowing down, Slowing down in your thinking, Joint pain or swelling, Limb weakness, Difficulty with concentration, Short term memory loss, Headache, Tingling feeling/pins and needles, Confusion/fuzzy head, Dizziness or light headedness, Chest tightness, Problems with balance, Altered personality/ behaviour, Chest pain, Palpitations, Leg/ankle swelling, Difficulty with communication, Skin rash, Diarrhoea, Problems seeing, Pain on breathing, Weight loss, Tremor/shakiness, Constipation, Erectile Dysfunction, Loss of sense of smell, Can’t fully move or control movement, Abdominal pain, Stomach pain, Loss of control of passing urine, Loss of appetite, Loss of taste, Nausea/vomiting, Bleeding, Can’t move and/or feel one side of your body or face, Loss of control of opening bowels, Lumpy lesions on toes, Fainting / blackouts, Seizures  Symptom severity was rated using a 0-10 visual analogue scale for Breathlessness, Cough, Fatigue, Sleep quality and Pain before COVID-19 illness and worst in last 24 hours. The results were dichotomised using cut off of ≤2 for no and ≥3 for Yes to combine the analysis with the longer list of symptoms.  An amendment was made Feb 2021 whereby ‘Breathlessness, Fatigue, Sleep Problems, Pain, and Cough’ were added to the symptom questionnaire with binary Y/N in addition to the VAS. These are therefore not comparable between five months and one year and were symbolised with not applicable ‘NA’ on Table S4. |
| Generalised Anxiety Disorder Questionnaire (GAD-7) (Anxiety) | The Generalised Anxiety Disorder (GAD-7) questionnaire is a patient reported outcome measure consists of 7 questions with total scores ranging from 0 to 21. We used a GAD7 threshold score of > 8 to suggest at least mild-moderate anxiety.^3^ |
| Patient Health Questionnaire (PHQ-9) (Depression) | The Patient Health Questionnaire (PHQ-9) is a patient reported outcome measure consisting of 9 questions with total scores ranging from 0 to 27. We used a PHQ-9 threshold score of ≥10 to suggest at least moderate depression.^4^ |
| Post-Traumatic Stress Disorder Checklist for DSM V (PCL-5) Questionnaire | The Post-Traumatic Stress Disorder Checklist for DSM V (PCL-5) questionnaire is a patient reported outcome measure consisting of 20 questions assessing evidence of post-traumatic stress disorder according to the DSM V criteria. Total scores range from 0-80. We used a PCL-5 threshold score of ≥38 suggestive of a provisional diagnosis of post-traumatic stress disorder.^5,6^ |
| Dyspnoea-12 | The Dyspnoea-12 questionnaire is a patient reported outcome measure consisting of 12 questions assessing breathlessness severity incorporating both “physical” and “affective” aspects.^7^ Scores range from 0 to 36 with higher scores correspond to greater severity of breathlessness. |
| FACIT fatigue subscale score (FACIT) | The Functional Assessment of Chronic Illness Therapy – Fatigue (FACIT-Fatigue) scale is a patient reported outcome measure consisting of 13 questions to assess self-reported fatigue and its impact on daily activities and function.^8^ Total scores range from 0-52, with lower scores corresponding to an increased burden of fatigue.^9^ |
| Brief Pain Inventory (BPI) severity and interference | The Brief Pain Inventory (BPI) is a patient reported outcome questionnaire consisting of 15 questions across domains of pain severity and pain interference. We have reported the BPI Severity score as the mean score from the 4 severity questions each with a range 0 – 10 anchored at 0 = “No Pain” and 10 = “Pain as bad as you can imagine”.^10,11^ |
| Short Physical Performance Battery (SPPB) | The Short Physical Performance Battery (SPPB) test is a researcher administer assessment of physical performance and frailty. It comprises 3 components; balance, gait speed and sit to stand tests. Tests were completed according to recommended standards and training was provided to site staff by the central study team via a recorded demonstration video. SPPB total scores range from 0-12. We have reported a total SPPB score of ≤10 suggestive of underlying frailty.^12-14^ |
| Incremental Shuttle Walk Test (ISWT) | The Incremental Shuttle Walk Test (ISWT) is a researcher administered assessment of maximal physical performance and was performed according to standardised instructions with two attempts performed by participants on the same day with a 20 minutes rest between them.^15^ Training was provided to site staff by the central study team via a recorded demonstration video. The best effort was reported in metres and the percent predicted value was calculated using the following reference formula accounting for gender, age and BMI.^16^ (ISWT predicted = 1449·701 − (11·735 × age) + (241·897 × gender) − (5·686 × BMI), where male gender = 1 and female gender = 0) |
| Rockwood Clinical Frailty Scale (CFS) | The Rockwood Clinical Frailty Scale (CFS) is a researcher assessed scale of clinical frailty with scores ranging from 1-9 where lower scores correspond to increased frailty. We have reported CFS scores of <5 suggestive of frailty.^17^ |
| Montreal Cognitive Assessment (MoCA) | The Montreal Cognitive Assessment (MoCA) is a researcher administered cognitive function questionnaire across 8 domains. Training was provided to site staff using standardised resources supplied online by MoCA TEST Inc.^18^ The assessment was conducted in English with researchers applying their discretion to exclude participants whose command of English was insufficient to complete the test accurately. Total scores range from 0 to 30. We report total MoCA scores of <23 suggestive of at least Mild Cognitive Impairment.^19^ |
| Spirometry and Pulmonary Function Testing | Due to COVID-19 related restrictions on aerosol-generating procedures during the study period, access to spirometry and lung function was limited. Spirometry and Pulmonary function testing was completed as per ERS/ATS recommendations.^20^ Spirometry were converted to SI units if not reported as such by sites. ERS Reference values were used to calculate % predicted values.^21^ FEV_1_/FVC <0·7 was used to define airflow obstruction.^22^ |
| BNP / NT-pro BNP | Brain Natriuretic Peptide (BNP) or N-terminal pro B-type Natriuretic Peptide (NT-pro BNP) were collected by according to each site’s routine clinically available assay as a biomarker of heart failure. Three sites submitted BNP results with all of the remaining sites submitting NT-pro BNP results. The threshold values used for BNP was ≥ 100 ng/litre ^23^ and for NT-pro BNP ≥ 400ng/litre ^24^ as suggestive of heart failure. |
| Glycated haemoglobin (HbA1c) | Glycated haemoglobin (HbA1c) was collected as a biomarker of current glycaemic control. We have reported HbA1c levels ≥ 6·5% as suggestive of a diagnosis of diabetes.^25^ |
| C-Reactive Protein (CRP) | C-Reactive Protein (CRP) levels were collected as a biomarker of current systemic inflammation. Values reported as below the lower or upper limit reportable range for the assay used at the site have been included at the stated less than or more than cut off value for calculation of mean (SD) results. We have reported CRP levels > 5mg/L as suggestive of systemic inflammation. |
| **Figure 3 and Table S9** |  |
| EQ5D-5L VAS | The EQ5D Visual Analogue Scale is a patient reported outcome questionnaire recording the patient’s self-rated health and was completed for “before your COVID-19 illness” and “your own health state today.” Scores are presented as mean and standard deviation.^26^ |
| EQ5D-5L Utility Index | The EQ5D-5L is a five-dimension patient reported outcome questionnaire recording a patient’s self-rated health state for mobility, self-care, usual activities, pain/discomfort and anxiety/depression. These scores are then mapped to a United Kingdom specific Utility Index anchored at 1 for “perfect health” and 0 for “dead” calculated from reported EQ5D-5L scores across the five dimensions.^27^ |
| Washington Group Short Set of Functioning Severity Continuum | The Washington Group Short Set of Functioning (WG-SS) is a patient reported outcome questionnaire using six questions to assess disability and function. Participant responses were transformed to the “Severity Continuum” by assigning scores of zero to responses “no difficulty”, one to responses “some difficulty”, six to responses “a lot of difficulty” and 36 to responses “cannot do at all”. ^28^ |

**Table S3. Comparison of individual and admission characteristics, and patient-perceived recovery between the one-year visit attendees and non-attendees for participants discharged between February 2020 – June 2020.**

**a) Comparison of individual and admission characteristics between the one-year visit attendees and non-attendees for participants discharged between February 2020 – June 2020 (n=1053)**

|  | Participants who attended the five-year visit but did not attend the one-year visit | Participants who attended both the five-month and one-year visits |
| --- | --- | --- |
| Total N (%) | 350 (32·9%) | 713 (62·7%) |
| Age, years† | 55·8 (13·3) | 58·3 (12·7) |
| Sex at birth |  |  |
| Male | 204 (59·8) | 445 (64·1) |
| Female | 137 (40·2) | 249 (35·9) |
| Missing | 9 | 19 |
| **Ethnicity** |  |  |
| White | 218 (64·5) | 518 (73·3) |
| South Asian | 71 (21·0) | 84 (11·9) |
| Black | 32 (9·5) | 55 (7·8) |
| Mixed | < 5 | 16 (2·3) |
| Other | 13 (3·8) | 34 (4·8) |
| Missing | 12 | 6 |
| **Health-care worker** |  |  |
| Yes | 86 (27·4) | 142 (21·5) |
| Missing | 36 | 54 |
| **IMD** |  |  |
| 1 - most deprived | 73 (21·9) | 136 (19·3) |
| 2 | 81 (24·3) | 147 (20·9) |
| 3 | 62 (18·6) | 135 (19·2) |
| 4 | 60 (18·0) | 125 (17·8) |
| 5 - least deprived | 58 (17·4) | 161 (22·9) |
| Missing | 16 | 9 |
| **BMI** †† |  |  |
| BMI < 30 kg/m^2^ | 142 (48·8) | 284 (42·2) |
| BMI ≥ 30 kg/m^2^ | 149 (51·2) | 389 (57·8) |
| Missing | 59 | 40 |
| **Smoking status** |  |  |
| Never | 177 (61·9) | 345 (54·8) |
| Ex-smoker | 103 (36·0) | 278 (44·1) |
| Current smoker | 6 (2·1) | 7 (1·1) |
| Missing | 64 | 83 |
| **WHO clinical progression scale** |  |  |
| WHO – class 3-4 | 89 (25·6) | 128 (18·0) |
| WHO – class 5 | 102 (29·3) | 260 (36·5) |
| WHO – class 6 | 56 (16·1) | 121 (17·0) |
| WHO – class 7-9 | 101 (29·0) | 204 (28·6) |
| Missing | < 5 | 0 |
| **Comorbidities** |  |  |
| No comorbidity | 101 (28·9) | 195 (27·3) |
| 1 comorbidity | 86 (24·6) | 139 (19·5) |
| 2+ comorbidities | 163 (46·6) | 379 (53·2) |
| Cardiovascular | 137 (39·1) | 324 (45·4) |
| Metabolic/endocrine/renal | 31 (8·9) | 74 (10·4) |
| Respiratory | 88 (25·1) | 186 (26·1) |
| Type 2 Diabetes | 73 (20·9) | 143 (20·1) |
| Neurological and psychiatric | 61 (17·4) | 124 (17·4) |
| **Hospital admission characteristics** |  |  |
| Admission duration, days † | 15·4 (18·2) | 16·6 (18·3) |
| SARS-CoV-2 swab |  |  |
| Negative | 41 (12·6) | 59 (8·6) |
| Positive | 285 (87·4) | 620 (90·6) |
| Indeterminate | 0 (0·0) | 5 (0·7) |
| Missing | 24 | 29 |
| Systemic steroids | 57 (18·0) | 167 (25·4) |
| Missing | 33 | 56 |
| Antibiotic therapy | 288 (84·0) | 583 (84·5) |
| Missing | 7 | 23 |
| Therapeutic dose anti-coagulation | 112 (34·3) | 244 (36·5) |
| Missing | 23 | 44 |

**b) Comparison of patient-perceived recovery at the five-month visit between attendees and non-attendees at the one-year visit for participants discharged between February – June 2020.**

|  |  | Participants who attended the five-year visit but did not attend the one-year visit | Participants who attended both the five-month and one-year visits |
| --- | --- | --- | --- |
| Total N (%) |  | 350 (32·9%) | 713 (62·7%) |
| Do you feel fully recovered from COVID-19? | Yes (Recovered) | 86 (31·6) | 152 (26·4) |
|  | Not sure | 56 (20·6) | 110 (19·1) |
|  | No (Not recovered) | 130 (47·8) | 313 (54·4) |
|  | (Missing) | 78 | 138 |

**Table S4. Comparison between imputed and non-imputed logistic regression of predictors of failure to recover at 1-year (multi-variable and multi-level).**

| **Dependent: ‘Fully recovered’** | **No** | **Yes** | **OR (univariable)** | **OR (multivariable)** | **OR (multivariable imputation)** | **OR (multilevel imputation)** |
| --- | --- | --- | --- | --- | --- | --- |
| **Age at admission, years** |  |  |  |  |  |  |
| 50-59 | 190 (82·6) | 40 (17·4) | - | - | - | - |
| <30 | 6 (54·5) | 5 (45·5) | 3·96 (1·09-13·77, p=0·029) | 2·13 (0·32-12·58, p=0·406) | 4·10 (1·04-16·21, p=0·045) | 3·65 (0·87-15·27, p=0·076) |
| 30-39 | 30 (60·0) | 20 (40·0) | 3·17 (1·62-6·12, p=0·001) | 2·46 (1·03-5·80, p=0·040) | 3·30 (1·63-6·67, p=0·001) | 3·09 (1·48-6·47, p=0·003) |
| 40-49 | 70 (68·0) | 33 (32·0) | 2·24 (1·31-3·83, p=0·003) | 1·53 (0·76-3·07, p=0·227) | 2·22 (1·24-3·96, p=0·007) | 1·88 (1·02-3·45, p=0·041) |
| 60-69 | 169 (70·7) | 70 (29·3) | 1·97 (1·27-3·08, p=0·003) | 1·62 (0·93-2·87, p=0·091) | 2·06 (1·29-3·29, p=0·003) | 1·93 (1·19-3·12, p=0·007) |
| 70-79 | 86 (65·2) | 46 (34·8) | 2·54 (1·55-4·18, p<0·001) | 2·15 (1·15-4·08, p=0·017) | 2·40 (1·41-4·08, p=0·001) | 2·08 (1·20-3·61, p=0·009) |
| 80+ | 16 (48·5) | 17 (51·5) | 5·05 (2·35-10·93, p<0·001) | 2·13 (0·79-5·70, p=0·131) | 4·26 (1·87-9·67, p=0·001) | 4·10 (1·77-9·48, p=0·001) |
| **Sex at birth** |  |  |  |  |  |  |
| Male | 341 (67·8) | 162 (32·2) | - | - | - | - |
| Female | 204 (75·6) | 66 (24·4) | 0·68 (0·49-0·95, p=0·024) | 0·73 (0·47-1·13, p=0·163) | 0·64 (0·44-0·93, p=0·019) | 0·68 (0·46-0·99, p=0·047) |
| **Ethnicity** |  |  |  |  |  |  |
| White | 441 (73·5) | 159 (26·5) | - | - | - | - |
| South Asian | 55 (64·7) | 30 (35·3) | 1·51 (0·93-2·43, p=0·091) | 1·36 (0·73-2·53, p=0·327) | 1·41 (0·82-2·43, p=0·209) | 1·45 (0·82-2·54, p=0·201) |
| Black | 35 (63·6) | 20 (36·4) | 1·58 (0·88-2·80, p=0·119) | 1·68 (0·74-3·69, p=0·202) | 1·98 (1·03-3·79, p=0·040) | 2·44 (1·23-4·82, p=0·011) |
| Mixed | 10 (52·6) | 9 (47·4) | 2·50 (0·98-6·30, p=0·051) | 2·24 (0·70-7·26, p=0·169) | 2·46 (0·91-6·66, p=0·077) | 2·95 (1·02-8·52, p=0·045) |
| Other | 23 (65·7) | 12 (34·3) | 1·45 (0·68-2·93, p=0·315) | 1·82 (0·61-5·14, p=0·268) | 2·15 (0·96-4·82, p=0·062) | 1·77 (0·74-4·25, p=0·201) |
| **IMD** |  |  |  |  |  |  |
| 1 - most deprived | 119 (74·4) | 41 (25·6) | - | - | - | - |
| 2 | 116 (75·8) | 37 (24·2) | 0·93 (0·55-1·55, p=0·768) | 0·80 (0·41-1·53, p=0·500) | 0·88 (0·51-1·54, p=0·659) | 1·00 (0·56-1·81, p=0·992) |
| 3 | 110 (70·5) | 46 (29·5) | 1·21 (0·74-1·99, p=0·443) | 1·29 (0·69-2·44, p=0·429) | 1·33 (0·77-2·27, p=0·306) | 1·34 (0·75-2·38, p=0·317) |
| 4 | 94 (66·7) | 47 (33·3) | 1·45 (0·88-2·40, p=0·143) | 1·29 (0·67-2·51, p=0·442) | 1·50 (0·87-2·56, p=0·141) | 1·60 (0·91-2·83, p=0·105) |
| 5 - least deprived | 128 (69·2) | 57 (30·8) | 1·29 (0·81-2·08, p=0·287) | 1·00 (0·54-1·89, p=0·990) | 1·21 (0·72-2·04, p=0·471) | 1·33 (0·76-2·32, p=0·318) |
| **No· of comorbidities**¶ |  |  |  |  |  |  |
| No comorbidity | 151 (68·6) | 69 (31·4) | - | - | - | - |
| 1 comorbidity | 93 (63·7) | 53 (36·3) | 1·25 (0·80-1·94, p=0·327) | 1·43 (0·79-2·57, p=0·233) | 1·36 (0·84-2·20, p=0·214) | 1·37 (0·83-2·27, p=0·216) |
| 2+ comorbidities | 328 (74·9) | 110 (25·1) | 0·73 (0·51-1·05, p=0·090) | 0·71 (0·43-1·19, p=0·194) | 0·77 (0·51-1·15, p=0·204) | 0·75 (0·49-1·16, p=0·197) |
| **BMI**¶ |  |  |  |  |  |  |
| BMI < 30 kg/m^2 | 177 (61·2) | 112 (38·8) | - | - | - | - |
| BMI ≥ 30 kg/m^2 | 254 (77·2) | 75 (22·8) | 0·47 (0·33-0·66, p<0·001) | 0·52 (0·34-0·79, p=0·002) | 0·53 (0·36-0·77, p=0·001) | 0·50 (0·34-0·74, p=0·001) |
| **WHO Class**¶ |  |  |  |  |  |  |
| WHO – class 3-4 | 100 (68·0) | 47 (32·0) | - | - | - | - |
| WHO – class 5 | 196 (66·0) | 101 (34·0) | 1·10 (0·72-1·68, p=0·669) | 1·31 (0·76-2·29, p=0·342) | 1·18 (0·74-1·87, p=0·486) | 1·23 (0·77-1·98, p=0·391) |
| WHO – class 6 | 95 (69·3) | 42 (30·7) | 0·94 (0·57-1·55, p=0·811) | 1·44 (0·73-2·83, p=0·292) | 1·23 (0·70-2·16, p=0·478) | 1·27 (0·71-2·26, p=0·428) |
| WHO – class 7-9 | 179 (81·0) | 42 (19·0) | 0·50 (0·31-0·81, p=0·005) | 0·40 (0·19-0·80, p=0·011) | 0·45 (0·25-0·80, p=0·006) | 0·42 (0·23-0·76, p=0·005) |
| **Steroids**¶ |  |  |  |  |  |  |
| No | 355 (68·8) | 161 (31·2) | - | - | - | - |
| Yes | 164 (75·6) | 53 (24·4) | 0·71 (0·49-1·02, p=0·066) | 1·18 (0·71-1·95, p=0·525) | 1·09 (0·71-1·67, p=0·707) | 1·05 (0·66-1·65, p=0·839) |
| **Anticoagulation**¶ |  |  |  |  |  |  |
| No | 314 (68·9) | 142 (31·1) | - | - | - | - |
| Yes | 210 (73·7) | 75 (26·3) | 0·79 (0·57-1·10, p=0·161) | 0·94 (0·58-1·50, p=0·783) | 0·95 (0·64-1·42, p=0·817) | 0·94 (0·62-1·44, p=0·785) |
| **Time from discharge to review (days)** |  |  |  |  |  |  |
| Mean (SD) | 382·2 (35·2) | 386·2 (32·0) | 1·00 (1·00-1·01, p=0·132) | 1·00 (1·00-1·01, p=0·163) | 1·00 (1·00-1·01, p=0·052) | 1·00 (1·00-1·01, p=0·118) |

OR = Odds Ratio, Data are n (%) unless otherwise stated. ¶ = % of category with positive response BMI = Body Mass Index, IMD = Indices of Multiple Deprivation, WHO = World Health Organisation. Category 3-4 = no continuous supplemental oxygen needed, 5= continuous supplemental oxygen only, 6= Continuous or Bi-level Positive Airway Pressure ventilation or High Flow Nasal O2, 7-9 = Invasive Mechanical Ventilation or other organ support.

**Table S5. Cluster medoids and characteristics**

### Cluster medoids (z-scores)

| Cluster | Anxiety (GAD7) | Depression (PHQ9) | PTSD (PCL5) | Breathlessness (Dyspnoea-12) | Fatigue (FACIT) | Physical Performance (SPPB) | Cognition (MoCA) |
| --- | --- | --- | --- | --- | --- | --- | --- |
| 1 | 1·5667731 | 1·5539296 | 1·1471037 | 1·2572720 | 1·3207526 | 0·39292536 | -0·05886651 |
| 2 | 0·1412077 | -0·1375875 | 0·1472313 | 0·2470335 | 0·3144553 | -0·04334305 | -0·05886651 |
| 3 | -0·7497707 | -0·9064589 | -0·7350089 | -0·5106454 | -0·4596196 | 0·82919376 | 1·40071221 |
| 4 | -0·7497707 | -0·9064589 | -0·7350089 | -0·7632050 | -0·9240646 | -0·47961145 | -0·35078225 |

### Cluster characteristics

| Cluster | Size | Maximal dissimilarity | Average dissimilarity | Isolation |
| --- | --- | --- | --- | --- |
| 1 | 330 | 6·751081 | 2·465918 | 2·369638 |
| 2 | 513 | 4·339742 | 1·800711 | 1·942071 |
| 3 | 184 | 6·444791 | 1·801180 | 2·865002 |
| 4 | 664 | 2·457693 | 1·059864 | 1·099839 |

GAD-7 = General Anxiety Disorder 7 Questionnaire, PHQ-9 = Patient Health Questionnaire-9, PCL-5 = Post

Traumatic Stress Disorder Checklist, Dyspnoea-12 Questionnaire, FACIT Fatigue Scale - Functional Assessment of Chronic Illness Therapy – Fatigue, BPI =Brief Pain Inventory, SPPB = Short Physical Performance Battery, MoCA = Montreal Cognitive Assessment.

**Table S6. Comparison of participant demographics, clinical characteristics, and admission characteristics stratified by the four clinical recovery clusters.**

|  | **1: Very severe** | **2: Severe** | **3: Moderate / Cognitive** | **4: Mild** | **Total** | **p** |
| --- | --- | --- | --- | --- | --- | --- |
| Total N (%) | 319 (19·5) | 493 (30·1) | 179 (10·9) | 645 (39·4) | 1636 |  |
| Age, years† | 53·9 (11·3) | 56·5 (11·6) | 67·8 (11·4) | 57·7 (12·9) | 57·7 (12·6) | <0·0001 |
| **Sex at birth** |  |  |  |  |  |  |
| Male | 141 (46·1) | 249 (53·4) | 123 (71·9) | 447 (71·6) | 960 (61·3) | <0·0001 |
| Female | 165 (53·9) | 217 (46·6) | 48 (28·1) | 177 (28·4) | 607 (38·7) |  |
| Missing | 13 | 27 | 8 | 21 | 69 |  |
| **Ethnicity** |  |  |  |  |  |  |
| White | 234 (77·0) | 384 (79·8) | 129 (72·5) | 483 (77·0) | 1230 (77·4) | 0·21 |
| South Asian | 32 (10·5) | 53 (11·0) | 22 (12·4) | 82 (13·1) | 189 (11·9) |  |
| Black | 21 (6·9) | 21 (4·4) | 19 (10·7) | 30 (4·8) | 91 (5·7) |  |
| Mixed | 5 (1·6) | 9 (1·9) | < 5 | 9 (1·4) | 24 (1·5) |  |
| Other | 12 (3·9) | 14 (2·9) | 7 (3·9) | 23 (3·7) | 56 (3·5) |  |
| Missing | 15 | 12 | < 5 | 18 | 46 |  |
| **Health-care worker** |  |  |  |  |  |  |
| Yes | 51 (17·5) | 92 (20·0) | 10 (5·8) | 96 (16·1) | 249 (16·4) | 0·0019 |
| No | 236 (80·8) | 363 (78·7) | 161 (93·6) | 494 (83·0) | 1254 (82·5) |  |
| Prefer not to say | 5 (1·7) | 6 (1·3) | < 5 | 5 (0·8) | 17 (1·1) |  |
| Missing | 27 | 32 | 7 | 50 | 116 |  |
| **IMD** |  |  |  |  |  |  |
| 1 - most deprived | 87 (27·8) | 98 (20·0) | 46 (26·3) | 94 (14·6) | 325 (20·0) | <0·0001 |
| 2 | 87 (27·8) | 114 (23·2) | 54 (30·9) | 126 (19·6) | 381 (23·5) |  |
| 3 | 47 (15·0) | 89 (18·1) | 26 (14·9) | 127 (19·8) | 289 (17·8) |  |
| 4 | 44 (14·1) | 88 (17·9) | 21 (12·0) | 141 (22·0) | 294 (18·1) |  |
| 5 - least deprived | 48 (15·3) | 102 (20·8) | 28 (16·0) | 154 (24·0) | 332 (20·5) |  |
| Missing | 6 | < 5 | < 5 | < 5 | 15 |  |
| **BMI** †† | 33·9 (29·2 - 40·2) | 31·9 (28·1 - 37·1) | 30·3 (27·4 - 34·1) | 30·0 (26·9 - 34·1) | 31·3 (27·7 - 36·0) | <0·0001 |
| BMI < 30 kg/m^2^ | 84 (29·2) | 159 (36·0) | 72 (47·1) | 286 (49·8) | 601 (41·2) | <0·0001 |
| BMI ≥ 30 kg/m^2^ | 204 (70·8) | 283 (64·0) | 81 (52·9) | 288 (50·2) | 856 (58·8) |  |
| Missing | 31 | 51 | 26 | 71 | 179 |  |
| **Smoking status** |  |  |  |  |  |  |
| Never | 155 (52·5) | 234 (50·8) | 88 (53·3) | 353 (60·8) | 830 (55·3) | 0·0056 |
| Ex-smoker | 132 (44·7) | 216 (46·9) | 76 (46·1) | 224 (38·6) | 648 (43·1) |  |
| Current smoker | 8 (2·7) | 11 (2·4) | < 5 | < 5 | 24 (1·6) |  |
| Missing | 24 | 32 | 14 | 64 | 134 |  |
| **WHO clinical progression scale** |  |  |  |  |  |  |
| WHO – class 3-4 | 57 (18·2) | 103 (21·4) | 22 (12·6) | 96 (15·1) | 278 (17·3) | 0·0040 |
| WHO – class 5 | 109 (34·7) | 191 (39·7) | 87 (49·7) | 278 (43·8) | 665 (41·4) |  |
| WHO – class 6 | 75 (23·9) | 107 (22·2) | 30 (17·1) | 148 (23·3) | 360 (22·4) |  |
| WHO – class 7-9 | 73 (23·2) | 80 (16·6) | 36 (20·6) | 113 (17·8) | 302 (18·8) |  |
| Missing | 5 | 12 | < 5 | 10 | 31 |  |
| **Comorbidities** |  |  |  |  |  |  |
| Number of comorbidities †† | 2 (1 - 4) | 2 (0 - 3) | 2 (1 - 4) | 1 (0 - 2) | 2 (0 - 3) | <0·0001 |
| No comorbidity | 64 (20·1) | 124 (25·2) | 42 (23·5) | 228 (35·3) | 458 (28·0) | <0·0001 |
| 1 comorbidity | 45 (14·1) | 111 (22·5) | 34 (19·0) | 158 (24·5) | 348 (21·3) |  |
| 2+ comorbidities | 210 (65·8) | 258 (52·3) | 103 (57·5) | 259 (40·2) | 830 (50·7) |  |
| Cardiovascular | 141 (44·2) | 207 (42·0) | 103 (57·5) | 259 (40·2) | 710 (43·4) | 0·00050 |
| Metabolic/endocrine/renal | 44 (13·8) | 53 (10·8) | 26 (14·5) | 46 (7·1) | 169 (10·3) | 0·0020 |
| Respiratory | 127 (39·8) | 129 (26·2) | 40 (22·3) | 122 (18·9) | 418 (25·6) | <0·0001 |
| Type 2 Diabetes | 62 (19·4) | 97 (19·7) | 46 (25·7) | 98 (15·2) | 303 (18·5) | 0·0095 |
| Rheumatological | 54 (16·9) | 58 (11·8) | 33 (18·4) | 43 (6·7) | 188 (11·5) | <0·0001 |
| Gastrointestinal | 55 (17·2) | 82 (16·6) | 21 (11·7) | 53 (8·2) | 211 (12·9) | <0·0001 |
| Neurological and psychiatric | 137 (42·9) | 108 (21·9) | 17 (9·5) | 45 (7·0) | 307 (18·8) | <0·0001 |
| Other comorbidities | 18 (5·6) | 24 (4·9) | 15 (8·4) | 30 (4·7) | 87 (5·3) | 0·24 |
| **Hospital admission characteristics** |  |  |  |  |  |  |
| Admission duration, days † | 14·6 (17·3) | 12·7 (16·7) | 15·9 (21·5) | 12·6 (15·9) | 13·4 (17·1) | 0·055 |
| **SARS-CoV-2 PCR¶** |  |  |  |  |  |  |
| Negative | 25 (8·7) | 35 (8·1) | 9 (5·4) | 44 (7·4) | 113 (7·6) | 0·68 |
| Positive | 263 (91·3) | 397 (91·5) | 157 (94·6) | 546 (92·1) | 1363 (92·0) |  |
| Indeterminate | 0 (0·0) | < 5 | 0 (0·0) | 3 (0·5) | 5 (0·3) |  |
| Missing | 31 | 59 | 13 | 52 | 155 |  |
| Systemic steroids | 177 (60·8) | 229 (49·7) | 85 (50·9) | 314 (52·2) | 805 (53·0) | 0·022 |
| Missing | 28 | 32 | 12 | 44 | 116 |  |
| Antibiotic therapy | 240 (77·9) | 369 (77·7) | 136 (78·6) | 498 (80·6) | 1243 (79·0) | 0·65 |
| Missing | 11 | 18 | 6 | 27 | 62 |  |
| Therapeutic dose anti-coagulation**§** | 136 (45·6) | 174 (37·9) | 68 (40·5) | 260 (43·0) | 638 (41·7) | 0·16 |
| Missing | 21 | 34 | 11 | 41 | 107 |  |

Data presented as n (%) were % is a positive response unless † mean [SD], †† median [IQR]. Percentages are calculated for each category after exclusion of missing data for that variable. P values across recovery clusters were calculated using a chi-squared test when testing for differences between proportions, ANOVA F-test for normally distributed continuous data and Kruskal Wallis for non-normally distributed continuous data. Non-normally distributed continuous variables summarised as median [IQR] and normally distributed continuous variables are presented. as mean [SD]. WHO classes are as follows: 3–4 = no continuous supplemental oxygen needed; 5 = continuous supplemental oxygen only; 6 = continuous or bi-level positive airway pressure ventilation or high-flow nasal oxygen; and 7–9 = invasive mechanical ventilation or other organ support. IMD = Index of Multiple Deprivation. BMI = body-mass index. PCR= Polymerase Chain Reaction. §Therapeutic dose anticoagulation; does not include intermediate doses which were not recorded.

**Table S7. Multinomial logistic regression of clinical characteristics against cluster membership (mild cluster as reference) for participants with plasma proteomic data. ***P<0·001, **P<0·01, * P<0·05.**

|  | Relative Risk Ratio | coefficient | Standard Error | Z | P |  |
| --- | --- | --- | --- | --- | --- | --- |
| **1: Very severe cluster vs mild cluster** | | | | | | |
| (Intercept) | 0·290 | -1·236 | 0·881 | -1·403 | 0·1605 |  |
| BMI | 1·059 | 0·057 | 0·017 | 3·387 | 0·0007 | *** |
| Age at admission | 0·973 | -0·027 | 0·011 | -2·437 | 0·0148 | * |
| Ethnicity: Black, mixed or other vs white | 0·892 | -0·114 | 0·381 | -0·299 | 0·7652 |  |
| Ethnicity: South Asian vs white | 0·544 | -0·608 | 0·420 | -1·446 | 0·1481 |  |
| Sex: Male vs Female | 0·435 | -0·833 | 0·254 | -3·276 | 0·0011 | ** |
| Comorbidity: 1 vs 0 | 0·973 | -0·028 | 0·401 | -0·069 | 0·9448 |  |
| Comorbidity: 2+ vs 0 | 3·703 | 1·309 | 0·318 | 4·122 | 3·76E-05 | *** |
| WHO: class 5 vs 3-4 | 0·679 | -0·387 | 0·339 | -1·142 | 0·2534 |  |
| WHO: class 6 vs 3-4 | 0·670 | -0·400 | 0·400 | -1·001 | 0·3170 |  |
| WHO: class 7-9 vs 3-4 | 1·278 | 0·245 | 0·359 | 0·683 | 0·4944 |  |
| **2: Severe cluster vs mild cluster** | | | | | | |
| (Intercept) | 0·341 | -1·077 | 0·774 | -1·391 | 0·1641 |  |
| BMI | 1·054 | 0·052 | 0·015 | 3·473 | 0·0005 | *** |
| Age at admission | 0·989 | -0·011 | 0·010 | -1·141 | 0·2539 |  |
| Ethnicity: Black, mixed or other vs white | 0·553 | -0·592 | 0·380 | -1·557 | 0·1194 |  |
| Ethnicity: South Asian vs white | 0·958 | -0·043 | 0·305 | -0·142 | 0·8870 |  |
| Sex: Male vs Female | 0·502 | -0·689 | 0·215 | -3·201 | 0·0014 | ** |
| Comorbidity: 1 vs 0 | 0·989 | -0·011 | 0·295 | -0·037 | 0·9705 |  |
| Comorbidity: 2+ vs 0 | 1·905 | 0·644 | 0·255 | 2·528 | 0·0115 | * |
| WHO: class 5 vs 3-4 | 0·560 | -0·580 | 0·281 | -2·060 | 0·0394 | * |
| WHO: class 6 vs 3-4 | 0·812 | -0·209 | 0·320 | -0·653 | 0·5139 |  |
| WHO: class 7-9 vs 3-4 | 0·846 | -0·167 | 0·309 | -0·540 | 0·5891 |  |
| **3: Moderate/cognitive cluster vs mild cluster** | | | | | | |
| (Intercept) | 0·000 | -9·593 | 1·462 | -6·563 | 5·27E-11 | *** |
| BMI | 1·032 | 0·032 | 0·022 | 1·443 | 0·1490 |  |
| Age at admission | 1·125 | 0·118 | 0·017 | 6·904 | 5·06E-12 | *** |
| Ethnicity: Black, mixed or other vs white | 1·265 | 0·235 | 0·520 | 0·452 | 0·6509 |  |
| Ethnicity: South Asian vs white | 1·445 | 0·368 | 0·512 | 0·719 | 0·4722 |  |
| Sex: Male vs Female | 0·728 | -0·317 | 0·315 | -1·007 | 0·3138 |  |
| Comorbidity: 1 vs 0 | 0·488 | -0·717 | 0·465 | -1·542 | 0·1230 |  |
| Comorbidity: 2+ vs 0 | 0·940 | -0·062 | 0·362 | -0·171 | 0·8639 |  |
| WHO: class 5 vs 3-4 | 0·913 | -0·091 | 0·432 | -0·210 | 0·8337 |  |
| WHO: class 6 vs 3-4 | 0·865 | -0·146 | 0·513 | -0·284 | 0·7764 |  |
| WHO: class 7-9 vs 3-4 | 2·173 | 0·776 | 0·456 | 1·702 | 0·0887 |  |

WHO classes are as follows: 3–4 = no continuous supplemental oxygen needed; 5 = continuous supplemental oxygen only; 6 = continuous or bi-level positive airway pressure ventilation or high-flow nasal oxygen; and 7–9 = invasive mechanical ventilation or other organ support. BMI = body-mass index.

**Table S8. Comparison of plasma proteomics between a) cluster 1 (very severe) and cluster 4 (mild), b) cluster 2 (severe) and cluster 4 (mild) and c) cluster 3 (moderate/cognitive) and cluster 4 (mild). Multinomial regression results with unadjusted P<0·05 are presented. **FDR<5% *FDR<10%**

**a) Cluster 1 (very severe) and cluster 4 (mild)**

| Protein name | coefficient | Lower CI | Upper CI | Standard Error | Z value | Unadjusted P | FDR adjusted P |
| --- | --- | --- | --- | --- | --- | --- | --- |
| TFF2 | 0·677 | 0·399 | 0·955 | 0·142 | 4·771 | 1·83E-06 | 5·43E-04** |
| TGFA | 0·489 | 0·239 | 0·739 | 0·127 | 3·840 | 1·23E-04 | 1·82E-02** |
| LAMP3 | 0·478 | 0·223 | 0·734 | 0·130 | 3·672 | 2·41E-04 | 2·38E-02** |
| CD83 | 0·468 | 0·209 | 0·727 | 0·132 | 3·542 | 3·98E-04 | 2·58E-02** |
| LGALS9 | 0·464 | 0·205 | 0·722 | 0·132 | 3·517 | 4·36E-04 | 2·58E-02** |
| PLAUR | 0·451 | 0·189 | 0·714 | 0·134 | 3·372 | 7·47E-04 | 3·69E-02** |
| IL6 | 0·432 | 0·170 | 0·694 | 0·134 | 3·233 | 1·23E-03 | 5·19E-02* |
| EPO | 0·369 | 0·129 | 0·609 | 0·122 | 3·013 | 2·59E-03 | 7·66E-02* |
| FLT3LG | 0·400 | 0·145 | 0·655 | 0·130 | 3·074 | 2·12E-03 | 7·66E-02* |
| AGRN | 0·432 | 0·151 | 0·712 | 0·143 | 3·017 | 2·55E-03 | 7·66E-02* |
| SCGB3A2 | 0·376 | 0·127 | 0·625 | 0·127 | 2·956 | 3·12E-03 | 7·69E-02* |
| FST | 0·353 | 0·121 | 0·585 | 0·118 | 2·980 | 2·88E-03 | 7·69E-02* |
| CLEC4D | 0·353 | 0·112 | 0·593 | 0·123 | 2·875 | 4·04E-03 | 9·21E-02* |
| CXCL17 | 0·420 | 0·126 | 0·715 | 0·150 | 2·797 | 5·16E-03 | 1·09E-01 |
| FCAR | 0·317 | 0·088 | 0·546 | 0·117 | 2·714 | 6·66E-03 | 1·19E-01 |
| SCG3 | 0·341 | 0·097 | 0·586 | 0·125 | 2·734 | 6·25E-03 | 1·19E-01 |
| CTRC | -0·316 | -0·546 | -0·087 | 0·117 | -2·705 | 6·83E-03 | 1·19E-01 |
| TNFRSF11B | 0·357 | 0·091 | 0·624 | 0·136 | 2·630 | 8·54E-03 | 1·40E-01 |
| CSF3 | 0·332 | 0·081 | 0·583 | 0·128 | 2·592 | 9·55E-03 | 1·49E-01 |
| VEGFA | 0·304 | 0·071 | 0·536 | 0·119 | 2·562 | 1·04E-02 | 1·54E-01 |
| ISM1 | 0·302 | 0·069 | 0·534 | 0·119 | 2·544 | 1·10E-02 | 1·54E-01 |
| LTBR | 0·330 | 0·074 | 0·587 | 0·131 | 2·526 | 1·15E-02 | 1·55E-01 |
| CLEC4C | 0·303 | 0·064 | 0·542 | 0·122 | 2·483 | 1·30E-02 | 1·68E-01 |
| CXCL8 | 0·292 | 0·058 | 0·525 | 0·119 | 2·447 | 1·44E-02 | 1·78E-01 |
| HGF | 0·295 | 0·054 | 0·537 | 0·123 | 2·395 | 1·66E-02 | 1·92E-01 |
| IL1RN | 0·294 | 0·053 | 0·535 | 0·123 | 2·390 | 1·69E-02 | 1·92E-01 |
| CXADR | 0·278 | 0·048 | 0·507 | 0·117 | 2·372 | 1·77E-02 | 1·94E-01 |
| OSM | 0·273 | 0·041 | 0·504 | 0·118 | 2·307 | 2·11E-02 | 2·23E-01 |
| PSPN | -0·253 | -0·476 | -0·029 | 0·114 | -2·218 | 2·65E-02 | 2·71E-01 |
| CD70 | 0·278 | 0·030 | 0·526 | 0·127 | 2·196 | 2·81E-02 | 2·77E-01 |
| LRRN1 | -0·271 | -0·517 | -0·025 | 0·126 | -2·158 | 3·09E-02 | 2·83E-01 |
| DPP10 | 0·245 | 0·023 | 0·468 | 0·114 | 2·160 | 3·07E-02 | 2·83E-01 |
| CNTNAP2 | 0·250 | 0·022 | 0·479 | 0·116 | 2·150 | 3·16E-02 | 2·83E-01 |
| TANK | 0·239 | 0·013 | 0·465 | 0·115 | 2·075 | 3·80E-02 | 3·12E-01 |
| ENAH | 0·236 | 0·011 | 0·460 | 0·115 | 2·055 | 3·98E-02 | 3·12E-01 |
| PROK1 | -0·238 | -0·466 | -0·010 | 0·116 | -2·045 | 4·09E-02 | 3·12E-01 |
| CCL20 | 0·233 | 0·009 | 0·457 | 0·114 | 2·042 | 4·11E-02 | 3·12E-01 |
| CD276 | 0·256 | 0·014 | 0·499 | 0·124 | 2·071 | 3·84E-02 | 3·12E-01 |
| LGALS4 | 0·279 | 0·016 | 0·542 | 0·134 | 2·079 | 3·76E-02 | 3·12E-01 |
| CCL7 | 0·223 | 0·001 | 0·444 | 0·113 | 1·971 | 4·87E-02 | 3·61E-01 |

**b) Cluster 2 (severe) and cluster 4 (mild)**

| Protein name | coefficient | Lower CI | Upper CI | Standard Error | Z value | Unadjusted P | FDR adjusted P |
| --- | --- | --- | --- | --- | --- | --- | --- |
| ICAM4 | -0·319 | -0·522 | -0·116 | 0·104 | -3·079 | 2·08E-03 | 6·15E-01 |
| TFF2 | 0·351 | 0·109 | 0·593 | 0·124 | 2·841 | 4·50E-03 | 6·66E-01 |

**c) Cluster 3 (moderate/cognitive) and cluster 4 (mild)**

| Protein name | coefficient | Lower CI | Upper CI | Standard Error | Z value | Unadjusted P | FDR adjusted P |
| --- | --- | --- | --- | --- | --- | --- | --- |
| IL6 | 0·573 | -0·184 | 0·291 | 0·158 | 3·630 | 2·84E-04 | 4·64E-02** |
| CD70 | 0·523 | 0·064 | 0·485 | 0·145 | 3·604 | 3·13E-04 | 4·64E-02** |
| LRRN1 | -0·372 | -0·403 | 0·018 | 0·150 | -2·488 | 1·29E-02 | 9·28E-01 |
| CSF3 | 0·406 | 0·050 | 0·477 | 0·165 | 2·466 | 1·37E-02 | 9·28E-01 |
| CD276 | 0·351 | -0·127 | 0·290 | 0·145 | 2·417 | 1·57E-02 | 9·28E-01 |

**Table S9: Ongoing symptoms recorded at five-months and one-year post-discharge from hospital**

|  | **N pairs** | **Five-month visit (paired)** | **One-year visit (paired)** | **p** | **One-year visit (all participants) Total n =924** | |
| --- | --- | --- | --- | --- | --- | --- |
|  |  |  |  |  | N with available data | n (%) |
| Any symptom n (%) | 619 | 580 (93·7) | 584 (94·3) | 0·67 | 817 | 773 (94·6) |
| Symptom count † | 619 | 9 (4 - 16) | 9 (4 - 17) | 0·010 | 817 | 10 (4 - 16) |
| Symptoms measured by VAS 0-10 scale (within the PSQ) |  |  |  |  |  |  |
| a) Breathlessness † | 524 | 2·0 (0·0 - 5·0) | 2·0 (0·0 - 5·0) | 0·052 | 747 | 2·0 (0·0 - 5·0) |
| b) Fatigue † | 521 | 3·0 (0·0 - 6·0) | 3·0 (0·0 - 6·0) | 0·090 | 752 | 3·0 (0·0 - 6·0) |
| c) Cough † | 518 | 0·0 (0·0 - 2·0) | 0·0 (0·0 - 2·0) | 0·43 | 751 | 0·0 (0·0 - 2·0) |
| d) Pain † | 514 | 1·0 (0·0 - 5·0) | 1·0 (0·0 - 5·0) | 0·24 | 751 | 1·0 (0·0 - 5·0) |
| e) Sleep quality † | 520 | 2·0 (0·0 - 6·0) | 2·0 (0·0 - 5·0) | 0·21 | 754 | 2·0 (0·0 - 5·0) |
| Fatigue | 533 | NA | 322 (60·4) | NA | 770 | 463 (60·1) |
| Aching in your muscles (pain) | 603 | 334 (55·4) | 324 (53·7) | 0·47 | 809 | 442 (54·6) |
| Physical slowing down | 609 | 313 (51·4) | 314 (51·6) | 1·0 | 811 | 429 (52·9) |
| Sleep disturbance | 530 | NA | 275 (51·9) | NA | 769 | 402 (52·3) |
| Breathlessness | 537 | NA | 281 (52·3) | NA | 769 | 395 (51·4) |
| Joint pain or swelling | 589 | 289 (49·1) | 287 (48·7) | 0·94 | 803 | 382 (47·6) |
| Slowing down in your thinking | 602 | 263 (43·7) | 279 (46·3) | 0·22 | 808 | 377 (46·7) |
| Pain | 527 | NA | 248 (47·1) | NA | 770 | 359 (46·6) |
| Short term memory loss | 595 | 247 (41·5) | 263 (44·2) | 0·23 | 808 | 360 (44·6) |
| Limb weakness | 607 | 289 (47·6) | 253 (41·7) | 0·010 | 813 | 341 (41·9) |
| Difficulty with concentration | 592 | 242 (40·9) | 243 (41·0) | 1·0 | 807 | 337 (41·8) |
| Tingling feeling/pins and needles | 599 | 243 (40·6) | 211 (35·2) | 0·014 | 813 | 285 (35·1) |
| Headache | 603 | 184 (30·5) | 195 (32·3) | 0·42 | 808 | 253 (31·3) |
| Confusion/fuzzy head | 606 | 181 (29·9) | 186 (30·7) | 0·73 | 811 | 250 (30·8) |
| Problems with balance | 601 | 210 (34·9) | 180 (30·0) | 0·0076 | 811 | 250 (30·8) |
| Dizziness or lightheaded | 594 | 175 (29·5) | 182 (30·6) | 0·60 | 810 | 243 (30·0) |
| Erectile Dysfunction | 328 | 87 (26·5) | 96 (29·3) | 0·25 | 491 | 144 (29·3) |
| Cough | 530 | NA | 148 (27·9) | NA | 771 | 215 (27·9) |
| Leg/ankle swelling | 602 | 156 (25·9) | 160 (26·6) | 0·79 | 810 | 223 (27·5) |
| Chest tightness | 599 | 157 (26·2) | 145 (24·2) | 0·32 | 807 | 198 (24·5) |
| Altered personality/behaviour (‘not the same person’) | 608 | 131 (21·5) | 123 (20·2) | 0·53 | 812 | 171 (21·1) |
| Difficulty with communication | 604 | 106 (17·5) | 124 (20·5) | 0·092 | 810 | 168 (20·7) |
| Palpitations | 588 | 122 (20·7) | 117 (19·9) | 0·69 | 803 | 165 (20·5) |
| Constipation | 600 | 106 (17·7) | 104 (17·3) | 0·91 | 811 | 141 (17·4) |
| Diarrhoea | 599 | 99 (16·5) | 89 (14·9) | 0·39 | 804 | 113 (16·5) |
| Skin rash | 563 | 100 (17·8) | 89 (15·8) | 0·32 | 776 | 127 (16·4) |
| Chest pain | 600 | 105 (17·5) | 87 (14·5) | 0·080 | 804 | 124 (15·4) |
| Abdominal pain | 604 | 91 (15·1) | 75 (12·4) | 0·11 | 808 | 119 (14·7) |
| Problems seeing | 590 | 100 (16·9) | 85 (14·4) | 0·14 | 807 | 115 (14·3) |
| Stomach pain | 584 | 81 (13·9) | 73 (12·5) | 0·45 | 802 | 108 (13·5) |
| Tremor/shakiness | 599 | 76 (12·7) | 76 (12·7) | 1·0 | 812 | 106 (13·1) |
| Pain on breathing | 593 | 65 (11·0) | 72 (12·1) | 0·47 | 807 | 106 (13·1) |
| Loss of appetite | 605 | 74 (12·2) | 69 (11·4) | 0·61 | 808 | 97 (12·0) |
| Loss of control of passing urine | 605 | 65 (10·7) | 68 (11·2) | 0·80 | 807 | 96 (11·9) |
| Loss of sense of smell | 601 | 67 (11·1) | 60 (10·0) | 0·46 | 808 | 86 (10·6) |
| Can’t fully move or control movement | 593 | 57 (9·6) | 56 (9·4) | 1·0 | 810 | 83 (10·2) |
| Loss of taste | 606 | 70 (11·6) | 53 (8·7) | 0·037 | 812 | 79 (9·7) |
| Nausea/vomiting | 597 | 62 (10·4) | 47 (7·9) | 0·078 | 809 | 71 (8·8) |
| Loss of control of opening your bowels | 600 | 35 (5·8) | 40 (6·7) | 0·54 | 807 | 58 (7·2) |
| Weight loss | 593 | 59 (9·9) | 42 (7·1) | 0·072 | 809 | 56 (6·9) |
| Lumpy lesions (purple/pink/bluish) on toes | 544 | 21 (3·9) | 27 (5·0) | 0·44 | 776 | 35 (4·5) |
| Bleeding | 551 | 26 (4·7) | 24 (4·4) | 0·88 | 781 | 34 (4·4) |
| Can’t move and / or feel one side of your body or face | 597 | 25 (4·2) | 17 (2·8) | 0·20 | 811 | 29 (3·6) |
| Fainting / blackouts | 588 | 9 (1·5) | 8 (1·4) | 1·0 | 808 | 15 (1·9) |
| Seizures | 592 | < 5 | < 5 | 1·0 | 808 | < 5 |

Data are n (%) where percentage indicates each category with positive response unless * mean (SD) and

† median (IQR). Symptoms listed in order of prevalence at 1-year for all participants with a 1-year visit. PSQ: Patient Symptom Questionnaire – bespoke questionnaire for PHOSP-COVID

**Table S10. Patient reported outcomes, physiological and biochemical tests stratified by patient perceived recovery outcomes at five-months and one-year visits**

a) **Paired data**

|  |  | **Five-month visit** | | | | **One-year visit** | | | |
| --- | --- | --- | --- | --- | --- | --- | --- | --- | --- |
|  | **Total N (Pairs)** | **Recovered** | **Not sure** | **Not recovered** | **p** | **Recovered** | **Not sure** | **Not**  **recovered** | **p** |
| Total N (%) | 590 | 151 (25·6) | 113 (19·2) | 326 (55·3) |  | 168 (28·5) | 138 (23·4) | 284 (48·1) |  |
| Time from discharge, days † | 590 | 182 (164 -199·5) | 180 (160 - 200) | 176 (151·2 - 197) | 0·14 | 392 (363 - 415) | 386 (350·8 - 406) | 383 (361·8 - 410) | 0·26 |
| **PROMS** |  |  |  |  |  |  |  |  |  |
| Any symptom at time point (%) ¶ | 588 | 122 (81·3) | 110 (97·3) | 320 (98·5) | <0·0001 | 141 (84·4) | 135 (97·8) | 279 (98·6) | <0·0001 |
| Median symptom count † | 588 | 3 (1 - 7) | 8 (4 - 13) | 13 (8 - 18) | <0·0001 | 3 (1 - 7) | 9 (5 - 15) | 14 (8 - 20·5) | <0·0001 |
| Fatigue VAS † | 496 | 0·0 (0·0 - 1·0) | 3·0 (0·0 - 5·0) | 5·0 (2·0 - 7·0) | <0·0001 | 0·0 (0·0 - 2·0) | 2·0 (0·0 - 5·0) | 5·0 (2·0 - 7·0) | <0·0001 |
| Breathlessness VAS † | 499 | 0·0 (0·0 - 1·0) | 1·0 (0·0 - 3·0) | 4·0 (1·0 - 6·0) | <0·0001 | 0·0 (0·0 - 1·0) | 2·0 (0·0 - 4·0) | 4·0 (1·0 - 6·0) | <0·0001 |
| GAD-7 total score* | 563 | 2·5 (4·0) | 4·7 (5·2) | 6·5 (6·1) | <0·0001 | 2·4 (3·8) | 4·9 (5·8) | 6·6 (6·0) | <0·0001 |
| Anxiety (GAD-7 >8)¶ | 527 | 15 (11·1) | 21 (20·8) | 96 (33·0) | <0·0001 | 12 (8·2) | 25 (20·0) | 82 (32·2) | <0·0001 |
| PHQ9 total score* | 561 | 3·0 (4·4) | 5·8 (5·6) | 8·3 (6·6) | <0·0001 | 2·7 (3·9) | 5·7 (5·8) | 8·6 (6·8) | <0·0001 |
| Depression (PHQ-9 ≥10)¶ | 525 | 13 (9·7) | 21 (21·0) | 110 (37·8) | <0·0001 | 10 (6·8) | 26 (21·0) | 97 (38·2) | <0·0001 |
| PCL-5 Total Severity Score* | 558 | 5·9 (9·6) | 14·1 (14·6) | 20·6 (19·1) | <0·0001 | 6·0 (9·3) | 13·0 (14·9) | 18·9 (18·3) | <0·0001 |
| PTSD (PCL-5 ≥38)¶ | 523 | < 5 (2·3) | 8 (7·9) | 54 (18·6) | <0·0001 | < 5 (2·0) | 11 (9·0) | 37 (14·6) | 0·00020 |
| Dyspnoea-12 score* | 534 | 1·5 (3·3) | 4·9 (7·0) | 8·5 (9·1) | <0·0001 | 1·7 (4·1) | 4·6 (6·4) | 8·4 (8·8) | <0·0001 |
| FACIT fatigue Subscale Score* | 521 | 44·5 (8·8) | 36·8 (11·1) | 30·5 (12·7) | <0·0001 | 44·1 (8·2) | 37·8 (10·3) | 30·1 (12·4) | <0·0001 |
| BPI severity* | 356 | 9·5 (10·2) | 11·2 (8·7) | 14·3 (9·6) | 0·00040 | 6·6 (7·1) | 13·5 (10·2) | 14·4 (9·0) | <0·0001 |
| BPI interference* | 341 | 13·1 (18·1) | 16·3 (15·8) | 23·5 (19·5) | 0·00010 | 9·6 (13·0) | 18·9 (17·5) | 22·0 (17·4) | <0·0001 |
| **Health-related quality of life and disability** |  |  |  |  |  |  |  |  |  |
| EQ5DL utility index † | 450 | 0·88 (0·75 - 1·00) | 0·74 (0·64 - 0·86) | 0·72 (0·55 - 0·80) | <0·0001 | 0·88 (0·77 - 1·00) | 0·77 (0·66 - 0·88) | 0·65 (0·49 - 0·77) | <0·0001 |
| EQ5D-5L VAS † | 451 | 90·0 (75·8 - 95·0) | 70·0 (60·0 - 85·0) | 70·0 (50·0 - 80·0) | <0·0001 | 82·5 (70·0 - 90·0) | 75·0 (70·0 - 90·0) | 70·0 (50·0 - 80·0) | <0·0001 |
| WG-SS-SCo † | 531 | 0·0 (0·0 - 1·0) | 1·5 (1·0 - 3·0) | 3·0 (1·0 - 8·0) | <0·0001 | 0·0 (0·0 - 2·0) | 1·0 (0·0 - 3·0) | 3·0 (1·0 - 8·0) | <0·0001 |
| **Physical performance** |  |  |  |  |  |  |  |  |  |
| SPPB total score (0-12)* | 519 | 10·5 (2·1) | 10·0 (2·0) | 9·6 (2·3) | 0·00050 | 10·6 (2·1) | 10·1 (2·2) | 9·5 (2·5) | 0·00010 |
| SPPB ≤10 (mobility disability)¶ | 519 | 47 (34·8) | 51 (50·5) | 160 (56·5) | 0·00020 | 50 (32·9) | 57 (47·5) | 142 (57·5) | <0·0001 |
| ISWT distance, m* | 381 | 531·1 (283·3) | 434·4 (219·8) | 417·7 (251·0) | 0·002 | 525·4 (285·2) | 454·2 (251·5) | 420·0 (252·9) | 0·0058 |
| ISWT % predicted* | 324 | 67·9 (31·4) | 58·6 (25·1) | 56·4 (28·7) | 0·013 | 67·3 (29·1) | 58·4 (27·8) | 56·9 (28·7) | 0·026 |
| **Frailty and Cognition** |  |  |  |  |  |  |  |  |  |
| Rockwood CSF ≥5¶ | 498 | < 5 (3·2) | < 5 (3·0) | 26 (9·5) | 0·017 | 8 (5·7) | 5 (4·1) | 21 (8·9) | 0·19 |
| MoCA total score* | 466 | 25·5 (3·9) | 26·7 (2·3) | 25·6 (3·5) | 0·020 | 26·8 (3·0) | 26·7 (2·8) | 26·5 (3·1) | 0·73 |
| MoCA <23¶ | 466 | 23 (19·3) | 4 (4·8) | 39 (14·8) | 0·012 | 12 (9·2) | 9 (8·5) | 21 (9·2) | 0·98 |
| MoCA (Adjusted) total score* | 466 | 25·8 (3·7) | 27·1 (2·2) | 26·0 (3·5) | 0·013 | 27·1 (2·9) | 27·1 (2·8) | 26·9 (3·1) | 0·82 |
| MOCA (adjusted) <23¶ | 466 | 17 (14·3) | < 5 (4·8) | 32 (12·2) | 0·090 | 10 (7·6) | 6 (5·7) | 20 (8·7) | 0·62 |
| **Lung physiology** |  |  |  |  |  |  |  |  |  |
| FEV1 (L)* | 245 | 2·9 (0·9) | 2·8 (0·7) | 2·9 (0·8) | 0·56 | 3·0 (0·8) | 2·9 (0·8) | 2·8 (0·9) | 0·33 |
| FEV1 % predicted* | 225 | 97·9 (30·5) | 94·7 (18·4) | 90·4 (19·3) | 0·099 | 98·2 (17·1) | 94·7 (29·7) | 91·5 (26·0) | 0·25 |
| FEV1 % predicted <80%¶ | 225 | 10 (18·2) | 8 (19·0) | 36 (28·1) | 0·25 | 10 (15·4) | 15 (24·6) | 24 (24·2) | 0·33 |
| FVC (L)* | 239 | 3·8 (1·0) | 3·7 (1·1) | 3·6 (1·1) | 0·61 | 4·0 (1·0) | 3·8 (1·0) | 3·5 (1·0) | 0·012 |
| FVC % predicted* | 219 | 99·0 (31·3) | 98·6 (17·5) | 88·1 (20·4) | 0·0037 | 101·6 (16·9) | 94·6 (30·2) | 89·1 (18·5) | 0·0023 |
| FVC % predicted <80%¶ | 219 | 11 (20·0) | < 5 | 46 (37·1) | 0·00050 | 7 (10·9) | 13 (22·4) | 28 (28·9) | 0·027 |
| FEV1/FVC* | 234 | 0·8 (0·1) | 0·8 (0·1) | 0·8 (0·1) | 0·096 | 0·8 (0·1) | 0·8 (0·1) | 0·8 (0·1) | 0·11 |
| FEV1/FVC <0·7¶ | 234 | 6 (10·2) | 5 (11·9) | 8 (6·0) | 0·38 | 6 (8·6) | 7 (11·5) | 9 (8·7) | 0·81 |
| **Biochemical Tests** |  |  |  |  |  |  |  |  |  |
| Pro-NT-BNP (ng/L)* | 224 | 247·7 (460·7) | 230·4 (560·5) | 136·6 (340·7) | 0·19 | 210·4 (435·1) | 249·4 (605·4) | 152·4 (376·4) | 0·41 |
| BNP/NT-Pro-BNP above threshold¶ | 242 | 10 (16·9) | 6 (13·3) | 10 (7·2) | 0·11 | 10 (14·9) | 7 (11·9) | 8 (6·9) | 0·21 |
| HbA1C % (DCCT/NGSP)* | 289 | 6·1 (1·0) | 6·2 (1·3) | 6·0 (1·0) | 0·63 | 6·1 (1·1) | 6·1 (1·3) | 6·0 (1·0) | 0·83 |
| HbA1C ≥6·0¶ | 289 | 26 (34·7) | 18 (34·6) | 52 (32·1) | 0·90 | 27 (30·7) | 19 (27·1) | 37 (28·2) | 0·88 |
| eGFR Result (ml/min/1·73 m^2^)* | 394 | 74·4 (15·8) | 76·1 (16·0) | 74·1 (15·2) | 0·63 | 75·1 (13·7) | 72·1 (17·3) | 74·4 (16·2) | 0·36 |
| eGFR < 60 ml/min/1·73 m^2^¶ | 397 | 16 (16·0) | 8 (10·7) | 35 (15·8) | 0·53 | 16 (15·2) | 16 (16·8) | 34 (17·3) | 0·90 |
| **Systemic Inflammation** |  |  |  |  |  |  |  |  |  |
| CRP (mg/L)* | 404 | 4·8 (12·5) | 4·1 (4·5) | 5·8 (8·1) | 0·34 | 3·5 (4·0) | 4·6 (4·9) | 5·4 (7·3) | 0·033 |
| CRP >5 mg/L¶ | 404 | 15 (15·3) | 17 (21·5) | 57 (25·1) | 0·15 | 20 (18·2) | 21 (23·3) | 55 (27·0) | 0·22 |
| Fibrinogen (g/L)* | 270 | 3·3 (0·8) | 3·5 (0·7) | 3·6 (0·9) | 0·036 | 3·4 (0·9) | 3·5 (0·8) | 3·6 (0·9) | 0·27 |
| Ferritin (µg/L)* | 311 | 110·1 (117·4) | 153·5 (146·3) | 153·7 (169·8) | 0·11 | 124·8 (113·1) | 157·0 (141·0) | 131·7 (162·5) | 0·37 |

**b) Unpaired data**

|  | **Five-month visit** | | | | | **One-year visit** | | | | |
| --- | --- | --- | --- | --- | --- | --- | --- | --- | --- | --- |
|  | **Total N**  **(five-month)** | **Recovered** | **Not sure** | **Not recovered** | **p** | **Total N (one-year)** | **Recovered** | **Not sure** | **Not recovered** | **p** |
| Total N (%) | 1965 (100·0) | 501 (25·5) | 385 (19·6) | 1079 (54·9) | - | 804 (100·0) | 232 (28·9) | 180 (22·4) | 392 (48·8) | - |
| Time from discharge, days † | 1965 (100·0) | 166 (12 -191) | 165 (122-191) | 157 (119-189) | 0·040 | 804 (100·0) | 390 (364-410) | 383 (353-405) | 383 (361-408) | 0·20 |
| **PROMS** |  |  |  |  |  |  |  |  |  |  |
| Any symptom at time point (%)¶ | 1962 (99·8) | 415 (83·0) | 369 (95·8) | 1066 (99·0) | <0·0001 | 803 (99·9) | 197 (84·9) | 175 (97·2) | 387 (99·0) | <0·0001 |
| Median number of symptoms | 1962 (99·8) | 3 (1 - 7) | 8 (4 - 15) | 14 (8 - 20) | <0·0001 | 803 (99·9) | 3 (1 - 7) | 10 (5 - 15) | 14 (8 - 21) | <0·0001 |
| Fatigue VAS | 1772 | 0·0 (0·0 - 2·0) | 2·0 (0·0 - 5·0) | 5·0 (2·0 - 8·0) | <0·0001 | 740 | 0·0 (0·0 - 2·0) | 3·0 (0·0 - 5·0) | 5·0 (3·0 - 7·0) | <0·0001 |
| Breathlessness VAS | 1782 | 0·0 (0·0 - 1·0) | 1·5 (0·0 - 4·0) | 4·0 (1·0 - 6·0) | <0·0001 | 735 | 0·0 (0·0 - 1·0) | 1·0 (0·0 - 4·0) | 4·0 (1·0 - 6·0) | <0·0001 |
| GAD7 total score* | 1882 (95·8) | 2·8 (4·2) | 4·9 (5·1) | 6·7 (6·1) | <0·0001 | 777 (96·6) | 2·4 (3·9) | 5·1 (5·8) | 6·9 (6·2) | <0·0001 |
| Anxiety (GAD7 >8)¶ | 1845 (93·9) | 53 (11·3) | 81 (22·6) | 339 (33·4) | <0·0001 | 732 (91·0) | 17 (7·9) | 36 (21·8) | 120 (34·0) | <0·0001 |
| PHQ9 total score* | 1883 (95·8) | 3·4 (4·6) | 6·2 (5·9) | 9·2 (6·7) | <0·0001 | 776 (96·5) | 2·7 (3·7) | 5·8 (5·9) | 8·9 (6·9) | <0·0001 |
| Depression (PHQ-9 ≥10)¶ | 1846 (93·9) | 47 (9·9) | 96 (26·7) | 426 (42·0) | <0·0001 | 731 (90·9) | 13 (6·1) | 37 (22·6) | 143 (40·5) | <0·0001 |
| PCL-5 Total Severity Score* | 1879 (95·6) | 7·4 (11·3) | 14·1 (15·1) | 20·7 (18·8) | <0·0001 | 770 (95·8) | 5·7 (9·3) | 13·8 (16·1) | 19·8 (18·8) | <0·0001 |
| PTSD (PCL-5 ≥38)¶ | 1844 (93·8) | 18 (3·8) | 34 (9·5) | 202 (20·0) | <0·0001 | 727 (90·4) | < 5 | 16 (9·9) | 61 (17·3) | <0·0001 |
| Dyspnoea-12 score* | 1847 (94·0) | 2·1 (4·8) | 5·1 (7·1) | 8·9 (8·8) | <0·0001 | 751 (93·4) | 1·7 (4·2) | 4·9 (6·7) | 8·7 (8·8) | <0·0001 |
| FACIT fatigue Subscale Score* | 1839 (93·6) | 43·6 (8·8) | 36·5 (11·2) | 29·1 (12·8) | <0·0001 | 722 (89·8) | 44·1 (7·9) | 37·7 (10·5) | 29·8 (12·6) | <0·0001 |
| BPI severity* | 1478 (75·2) | 9·2 (10·1) | 12·0 (9·5) | 15·2 (10·0) | <0·0001 | 610 (75·9) | 6·7 (8·1) | 11·9 (10·1) | 14·8 (9·4) | <0·0001 |
| BPI interference* | 1434 (73·0) | 11·2 (16·4) | 17·6 (17·3) | 24·8 (19·8) | <0·0001 | 583 (72·5) | 8·6 (13·4) | 17·6 (17·5) | 22·2 (18·2) | <0·0001 |
| **Health-related quality of life and disability** |  |  |  |  |  |  |  |  |  |  |
| EQ5DL utility index † | 1683 | 0·88 (0·75 - 1·00) | 0·77 (0·65 - 0·88) | 0·69 (0·52 - 0·80) | <0·0001 | 706 | 0·88 (0·77 - 1·00) | 0·75 (0·66 - 0·88) | 0·66 (0·43 - 0·77) | <0·0001 |
| EQ5D-5L VAS † | 1678 | 85·0 (72·2 - 91·2) | 75·0 (60·0 - 85·0) | 70·0 (50·0 - 80·0) | <0·0001 | 699 | 85·0 (70·0 - 90·0) | 78·5 (68·5 - 89·5) | 70·0 (50·0 - 80·0) | <0·0001 |
| WG-SS-SCo † | 1861 | 0·0 (0·0 - 2·0) | 2·0 (0·5 - 3·0) | 3·0 (1·0 - 8·0) | <0·0001 | 760 | 0·0 (0·0 - 2·0) | 2·0 (1·0 - 3·0) | 3·0 (2·0 - 8·0) | <0·0001 |
| **Physical performance** |  |  |  |  |  |  |  |  |  |  |
| SPPB total score (0-12)* | 1815 (92·4) | 10·3 (2·2) | 10·1 (2·2) | 9·4 (2·5) | <0·0001 | 746 (92·8) | 10·5 (1·9) | 10·0 (2·4) | 9·5 (2·4) | <0·0001 |
| SPPB (mobility disability ≤10¶ | 1815 (92·4) | 181 (38·9) | 166 (46·0) | 582 (58·8) | <0·0001 | 746 (92·8) | 81 (36·8) | 75 (45·5) | 205 (56·8) | <0·0001 |
| ISWT, m* | 1465 (74·6) | 487·6 (274·7) | 431·4 (242·3) | 384·6 (249·4) | <0·0001 | 549 (68·3) | 528·3 (271·0) | 458·2 (250·2) | 408·6 (249·2) | <0·0001 |
| ISWT % predicted* | 968 (49·3) | 63·5 (30·7) | 57·8 (28·1) | 52·5 (28·7) | <0·0001 | 452 (56·2) | 67·5 (27·3) | 60·9 (28·2) | 55·5 (28·9) | 0·00080 |
| **Frailty and Cognition** |  |  |  |  |  |  |  |  |  |  |
| Rockwood CSF ≥5¶ | 1779 (90·5) | 9 (2·0) | 17 (4·9) | 81 (8·2) | <0·0001 | 743 (92·4) | 13 (6·1) | 8 (4·7) | 36 (10·0) | 0·058 |
| MoCA total score* | 1645 (83·7) | 25·6 (3·4) | 25·8 (3·6) | 25·6 (3·5) | 0·55 | 694 (86·3) | 26·0 (4·1) | 26·4 (3·0) | 26·4 (3·1) | 0·30 |
| MoCA <23¶ | 1645 (83·7) | 66 (16·5) | 39 (12·4) | 147 (15·8) | 0·26 | 694 (86·3) | 31 (15·0) | 17 (11·3) | 38 (11·2) | 0·39 |
| MoCA (Adjusted) total score* | 1645 (83·7) | 25·9 (3·3) | 26·2 (3·6) | 25·9 (3·4) | 0·53 | 694 (86·3) | 26·3 (3·9) | 26·8 (3·0) | 26·8 (3·1) | 0·20 |
| MoCA (Adjusted) <23¶ | 1645 (83·7) | 57 (14·3) | 36 (11·4) | 125 (13·4) | 0·52 | 694 (86·3) | 27 (13·1) | 14 (9·3) | 35 (10·4) | 0·47 |
| **Lung physiology** |  |  |  |  |  |  |  |  |  |  |
| FEV1 (L)* | 1168 (59·4) | 2·9 (0·8) | 2·7 (0·8) | 2·7 (0·8) | 0·027 | 408 (50·7) | 3·0 (0·8) | 2·8 (0·9) | 2·8 (0·9) | 0·13 |
| FEV1 % predicted* | 826 (42·0) | 94·4 (22·7) | 91·2 (21·8) | 89·8 (19·7) | 0·030 | 354 (44·0) | 98·9 (24·2) | 92·8 (28·0) | 92·3 (23·0) | 0·090 |
| FEV1 % <80% predicted¶ | 826 (42·0) | 43 (21·4) | 43 (27·7) | 131 (27·9) | 0·20 | 354 (44·0) | 15 (15·0) | 24 (27·9) | 39 (23·2) | 0·093 |
| FVC (L)* | 1161 (59·1) | 3·6 (1·0) | 3·4 (1·0) | 3·4 (1·0) | 0·018 | 403 (50·1) | 3·9 (1·0) | 3·6 (1·1) | 3·5 (1·0) | 0·0061 |
| FVC % predicted* | 822 (41·8) | 94·7 (23·1) | 92·1 (22·2) | 88·1 (20·1) | 0·0007 | 349 (43·4) | 101·8 (26·2) | 92·1 (27·8) | 90·9 (17·9) | 0·00070 |
| FVC % <80% predicted¶ | 822 (41·8) | 40 (19·9) | 33 (21·4) | 155 (33·2) | 0·0003 | 349 (43·4) | 12 (12·1) | 23 (27·1) | 42 (25·5) | 0·018 |
| FEV1/FVC* | 1152 (58·6) | 0·8 (0·1) | 0·8 (0·1) | 0·8 (0·1) | 0·039 | 399 (49·6) | 0·8 (0·1) | 0·8 (0·1) | 0·8 (0·1) | 0·40 |
| FEV1/FVC <0·7¶ | 1152 (58·6) | 25 (8·8) | 23 (10·6) | 49 (7·5) | 0·35 | 399 (49·6) | 9 (8·3) | 11 (10·9) | 18 (9·5) | 0·82 |
| **Biochemical Tests** |  |  |  |  |  |  |  |  |  |  |
| BNP (ng/L)* | 93 (4·7) | 82·7 (222·5) | 98·4 (134·0) | 57·2 (64·8) | 0·60 | 24 (3·0) | 26·2 (28·6) | 54·0 (55·1) | 136·3 (178·3) | 0·090 |
| Pro-NT-BNP (ng/L)* | 1134 (57·7) | 199·3 (537·3) | 163·4 (375·6) | 149·1 (919·6) | 0·65 | 437 (54·4) | 212·5 (448·4) | 468·2 (2409·4) | 174·6 (395·2) | 0·12 |
| BNP/Pro-NT-BNP above threshold¶ | 1224 (62·3) | 27 (8·7) | 24 (10·3) | 35 (5·2) | 0·014 | 461 (57·3) | 18 (13·5) | 11 (11·0) | 22 (9·6) | 0·53 |
| HbA1C % (DCCT/NGSP)* | 1272 (64·7) | 6·1 (1·2) | 6·2 (1·3) | 6·2 (1·3) | 0·60 | 498 (61·9) | 6·1 (1·1) | 6·0 (1·2) | 6·2 (1·4) | 0·65 |
| HbA1C ≥6·0¶ | 1272 (64·7) | 113 (35·3) | 93 (37·2) | 267 (38·0) | 0·71 | 498 (61·9) | 49 (33·8) | 29 (26·4) | 82 (33·7) | 0·34 |
| eGFR (ml/min/1·73 m^2^)* | 1596 (81·2) | 76·7 (16·7) | 76·3 (15·5) | 75·7 (15·4) | 0·54 | 633 (78·7) | 74·1 (17·7) | 74·0 (17·6) | 74·6 (16·0) | 0·94 |
| eGFR <60 (ml/min/1·73 m^2^)¶ | 1607 (81·8) | 49 (12·0) | 35 (11·3) | 101 (11·4) | 0·94 | 635 (79·0) | 30 (17·4) | 21 (14·6) | 44 (13·8) | 0·55 |
| **Systemic Inflammation** |  |  |  |  |  |  |  |  |  |  |
| CRP (mg/L)* | 1594 (81·1) | 5·6 (10·3) | 5·2 (10·0) | 5·7 (7·5) | 0·77 | 641 (79·7) | 4·0 (5·0) | 4·6 (4·5) | 5·3 (6·3) | 0·045 |
| CRP >5 mg/L ¶ | 1594 (81·1) | 83 (20·9) | 75 (23·9) | 239 (27·1) | 0·052 | 641 (79·7) | 32 (18·1) | 36 (25·5) | 89 (27·6) | 0·059 |
| Fibrinogen (g/L)* | 1258 (64·0) | 3·5 (0·9) | 3·6 (0·8) | 3·6 (0·9) | 0·34 | 508 (63·2) | 3·4 (0·9) | 3·5 (0·8) | 3·6 (0·9) | 0·099 |
| Ferritin (µg/L)* | 1407 (71·6) | 136·3 (149·7) | 145·3 (150·6) | 140·1 (166·4) | 0·79 | 525 (65·3) | 145·1 (196·9) | 176·6 (232·5) | 130·3 (158·9) | 0·087 |

Data presented as n (%) unless † median [IQR], *mean [SD]. ¶ = % of category with positive response. P values across patient perceived recovery were calculated using a chi-squared test when testing for differences between proportions, ANOVA F-test for normally distributed continuous data and Kruskal Wallis for non-normally distributed continuous data. Non-normally distributed continuous variables summarised as median [IQR] and normally distributed continuous variables are presented as mean [SD]. Threshold of BNP ≥100ng/L or NT-BNP ≥400ng/L. DCCT/NGSP - Diabetes Control and Complications Trial / National Glycohemoglobin Standardization Programme, PROMS = Patient reported outcome measures, VAS = Visual Analogue Scale, GAD7 = General Anxiety Disorder 7 Questionnaire, PHQ-9 = Patient Health Questionnaire-9, PCL-5 = Post Traumatic Stress Disorder Checklist, Dyspnoea-12 Questionnaire, FACIT Fatigue Scale (FACIT), BPI =Brief Pain Inventory, WG-SS-SCo = Washington Group Short Set of Functioning Severity Continuum, SPPB = Short Physical Performance Battery, ISWT = Incremental Shuttle Walking Test, CFS = Clinical Frailty Scale, MoCA = Montreal Cognitive Assessment. MoCA adjusted = adjusted for education. FEV1 = Forced Expiratory Volume in 1 second, FVC = Forced Vital Capacity, TLCO = Transfer Capacity of the Lung for Carbon Monoxide, KCO = carbon monoxide transfer coefficient, BNP = Brain Natriuretic Peptide. Pro-NT-BNP = N-Terminal Brain Natriuretic Peptide, HbA1C = glycosylated haemoglobin, eGFR = estimated Glomerular Filtration Rate, CRP = C-reactive protein.

**Table S11. A comparison between the four clinical recovery phenotypes of health-related quality of life, disability, fatigue and breathlessness across pre-hospitalisation, and five-months and one-year post-discharge**

a) Data for participants with data at all three time points and for the whole cohort at the individual time-points

|  | Participants with complete data for all three time points | | | All Data | | |
| --- | --- | --- | --- | --- | --- | --- |
|  | Pre-COVID | 5-month | 1-year | Pre-COVID | 5-month | 1-year |
| EQ5D-5L Utility Index† | 0·88 (0·77-1·0) | 0·74 (0·63-0·88) | 0·75 (0·62-0·88) | 0·88 (0·74-1·0) | 0·75 (0·62-0·88) | 0·74 (0·59-0·88) |
| N | 505 | 505 | 505 | 1979 | 1923 | 778 |
| EQ5D-5L VAS† | 90 (75-95) | 75 (60-90) | 75 (60-90) | 85 (70-90) | 75 (60-85) | 75 (60-87·5) |
| N | 487 | 487 | 487 | 1849 | 1913 | 771 |
| WG-SS-SCo † | 2 (0-4) | 2 (0-4) | 2 (0-4) | 0 (0-2) | 2 (0-4) | 2 (0-3·5) |
| N | 541 | 541 | 541 | 1453 | 1386 | 511 |
| Breathlessness VAS† | 0 (0-1·25) | 2 (0-5) | 2 (0-5) | 0 (0-2) | 2 (0-5) | 2 (0-5) |
| N | 524 | 524 | 524 | 2130 | 1829 | 747 |
| Fatigue VAS† | 0 (0-2) | 3 (0-6) | 3 (0-6) | 0 (0-2) | 3 (0-7) | 3 (0-6) |
| N | 521 | 521 | 521 | 2123 | 1819 | 752 |

†Median [IQR]

b) Participants with complete data for all three time points stratified by the severity clusters

|  | **Cluster 1** | | | **Cluster 2** | | | **Cluster 3** | | | **Cluster 4** | | | **All Clusters** | | |
| --- | --- | --- | --- | --- | --- | --- | --- | --- | --- | --- | --- | --- | --- | --- | --- |
|  | Pre-COVID | 5-month | 1-year | Pre-COVID | 5-month | 1-year | Pre-COVID | 5-month | 1-year | Pre-COVID | 5-month | 1-year | Pre-COVID | 5-month | 1-year |
| EQ5D-5L Utility Index† | 0·84 (0·61-1·0) | 0·53 (0·27-0·63) | 0·43 (0·31-0·63) | 0·85 (0·77-1·0) | 0·73 (0·65-0·77) | 0·72 (0·64-0·82) | 0·88 (0·76-1·0) | 0·77 (0·70-0·87) | 0·77 (0·66-0·85) | 1·0 (0·84-1·0) | 0·88 (0·77-1·0) | 0·88 (0·77-1·0) | 0·89 (0·77-1·0) | 0·75 (0·65-0·88) | 0·77 (0·64-0·88) |
| N | 66 | 66 | 66 | 115 | 115 | 115 | 42 | 42 | 42 | 169 | 169 | 169 | 392 | 392 | 392 |
| EQ5D-5L VAS† | 80 (70-90) | 50 (35-66·25) | 50 (40-66·25) | 85( 75-90) | 70 (60-82·75) | 70 (60-80) | 90 (80-95) | 80 (70-90) | 80 (70-87·5) | 90 (80-95) | 85 (75-90) | 88 (75-92) | 90 (75·25-95) | 79 (60-90) | 80 (60-90) |
| N | 64 | 64 | 64 | 106 | 106 | 106 | 43 | 43 | 43 | 165 | 165 | 165 | 378 | 378 | 378 |
| WG-SS-SCo† | 1 (0-4) | 8 (2-14) | 8 (3-14) | 1 (0-2) | 2 (1-4) | 2 (1-4) | 1 (0-1) | 1 (0-2) | 2 (1-4) | 0 (0-0) | 0 (0-1) | 1 (0-2) | 0 (0-1) | 2 (0-3) | 2 (0-4) |
| N | 69 | 69 | 69 | 125 | 125 | 125 | 56 | 56 | 56 | 168 | 168 | 168 | 418 | 418 | 418 |
| Breathlessness VAS† | 0·5 (0-3) | 6 (4-8) | 5 (2·75-7) | 0 (0-1) | 3 (1-5) | 3 (0-5) | 0 (0-1) | 2 (0-5) | 2 (0-5) | 0 (0-0) | 0 (0-1·25) | 0 (0-1) | 0 (0-1) | 2 (0-5) | 2 (0-5) |
| N | 64 | 64 | 64 | 120 | 120 | 120 | 49 | 49 | 49 | 160 | 160 | 160 | 393 | 393 | 393 |
| Fatigue VAS† | 2 (0-4) | 7 (5-8) | 6 (4-8) | 1 (0-2) | 5 (2-7) | 5 (3-6) | 0 (0-1·25) | 2·5 (1-5) | 2·5 (0-5) | 0 (0-1) | 0 (0-2) | 0 (0-2) | 0 (0-2) | 3 (0-6) | 3 (0-5) |
| N | 63 | 63 | 63 | 121 | 121 | 121 | 48 | 48 | 48 | 158 | 158 | 158 | 390 | 390 | 390 |

†Median [IQR]

c) Data for the whole cohort at the individual time-points stratified by the severity clusters

| Unpaired measurements stratified by the severity clusters | | | | | | | | | | | | | | | |
| --- | --- | --- | --- | --- | --- | --- | --- | --- | --- | --- | --- | --- | --- | --- | --- |
|  | **Cluster 1** | | | **Cluster 2** | | | **Cluster 3** | | | **Cluster 4** | | | **All Clusters** | | |
|  | Pre-COVID | 5-month | 1-year | Pre-COVID | 5- month | 1-year | Pre-COVID | 5-month | 1-year | Pre-COVID | 5-month | 1-year | Pre-COVID | 5-month | 1-year |
| EQ5D-5L Utility Index† | 0·74 (0·52-0·88) | 0·49 (0·26-0·64) | 0·42 (0·28-0·63) | 0·84 (0·74-1·00) | 0·72 (0·65-0·78) | 0·72 (0·64-0·80) | 0·88 (0·77-1·00) | 0·80 (0·71-0·88) | 0·75 (0·65-0·85) | 1·00 (0·84-1·00) | 0·88 (0·77-1·00) | 0·88 (0·77-1·00) | 0·88 (0·77-1·00) | 0·77 (0·64-0·88) | 0·77 (0·64-0·88) |
| N | 265 | 278 | 86 | 409 | 438 | 150 | 143 | 161 | 62 | 546 | 573 | 222 | 1363 | 1450 | 520 |
| EQ5D-5L VAS† | 75 (60-87·25) | 50 (40-65·75) | 50 (40-70) | 80 (70-90) | 70 (60-80) | 70 (55-80) | 90 (80-95) | 80 (70-90) | 80 (70-90) | 90 (80-95) | 85 (75-90) | 87 (78-90) | 85 (75-90) | 75 (60-88) | 80 (60-100) |
| N | 250 | 282 | 86 | 384 | 435 | 149 | 135 | 165 | 64 | 519 | 574 | 220 | 1288 | 1456 | 519 |
| WG-SS-SCo † | 1 (0-4) | 8 (3-13) | 8 (3-13) | 1 (0-2) | 2 (1-4) | 2 (1-4) | 1 (0-2) | 1 (0-3) | 2 (1-4) | 0 (0-1) | 1 (0-1) | 1 (0-2) | 0 (0-2) | 2 (0-4) | 2 (0-3·5) |
| N | 284 | 276 | 81 | 445 | 421 | 154 | 161 | 156 | 66 | 563 | 533 | 210 | 1453 | 1386 | 511 |
| Breathlessness VAS† | 1 (0-3) | 5 (3-7) | 5 (3-7) | 0 (0-2) | 3 (1-5) | 3 (0-5) | 0 (0-2) | 1 (0-3) | 2 (0-5) | 0 (0-0) | 0 (0-2) | 0 (0-1·75) | 0 (0-2) | 2 (0-5) | 1 (0-5) |
| N | 291 | 265 | 77 | 449 | 411 | 142 | 167 | 145 | 59 | 578 | 511 | 206 | 1485 | 1332 | 484 |
| Fatigue VAS† | 2 (0-4) | 7 (5-8) | 6 (4-8) | 1 (0-2) | 5 (2-7) | 4 (2-6) | 0 (0-2) | 2 (0-4) | 2·5 (0-5) | 0 (0-1) | 0 (0-2) | 0 (0-2) | 0 (0-2) | 3 (0-6) | 3 (0-5) |
| N | 291 | 264 | 77 | 448 | 410 | 143 | 167 | 141 | 60 | 574 | 508 | 206 | 1480 | 1323 | 486 |

†Median [IQR], WG-SS-SCo = Washington Group Short Set of Functioning Severity Continuum, VAS = Visual Analogue Scale.

Supplement Figures

**Figure S1. A comparison of participant-perceived recovery between five months and one-year**

a) Paired data (n = 590)

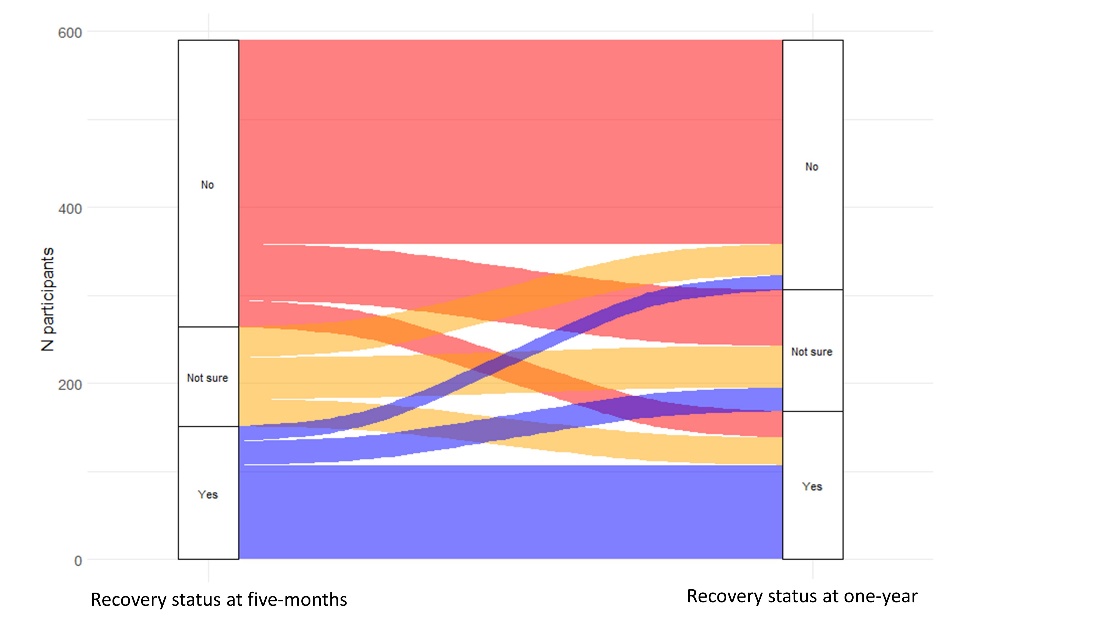

Figure S1a. Legend

Recovery status

| Five-month visit | One-year visit | Total (N = 590) |
| --- | --- | --- |
| No | No | 232 (39·3%) |
| Yes | Yes | 107 (18·1%) |
| Not sure | Not sure | 47 (8·0%) |
| No | Not sure | 64 (10·8%) |
| No | Yes | 30 (5·1%) |
| Not sure | No | 35 (5·9%) |
| Not sure | Yes | 31 (5·3%) |
| Yes | No | 17 (2·9%) |
| Yes | Not sure | 27 (4·6%) |

b) All patients who had had research visits at the time of writing.

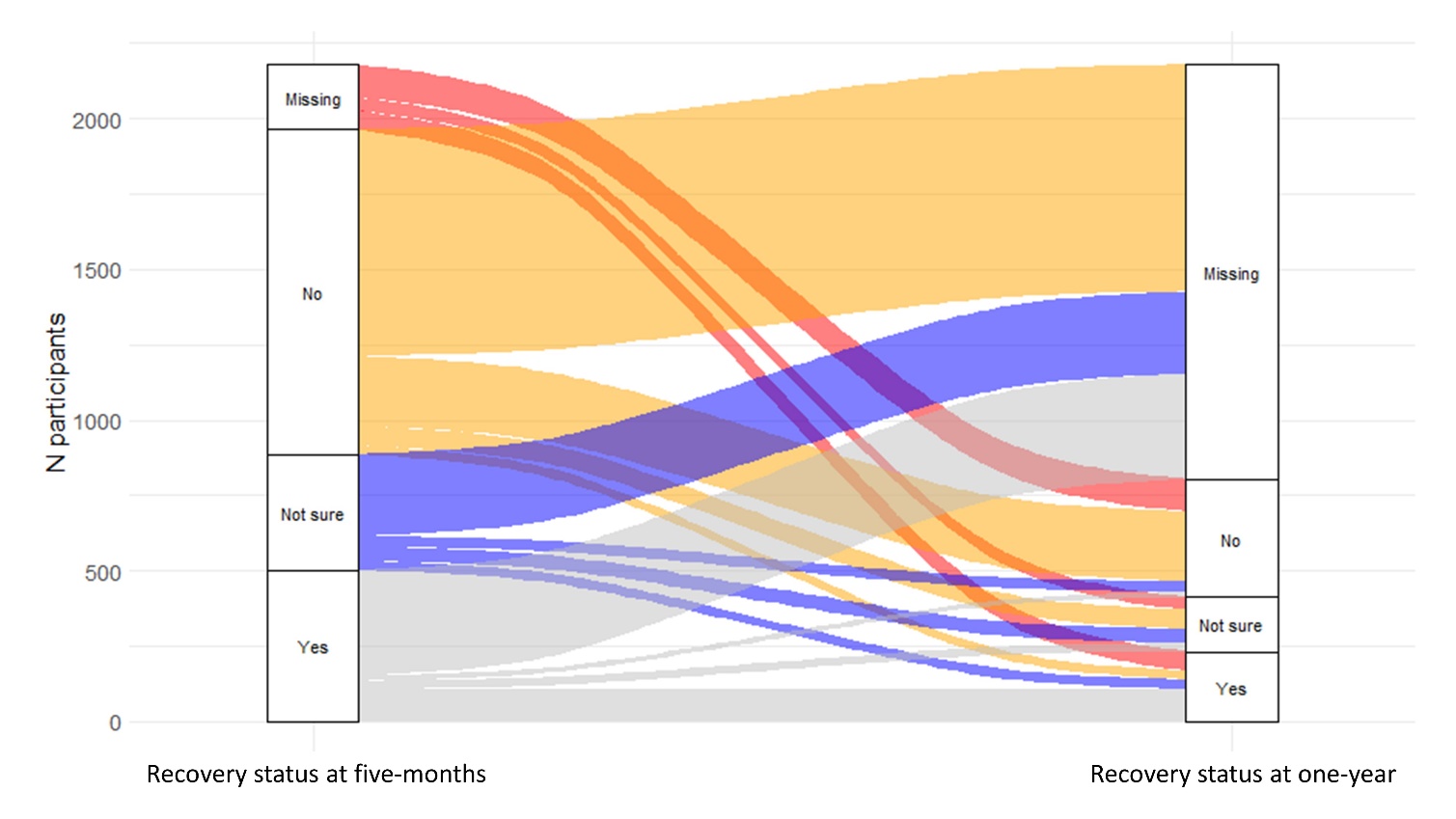

Figure S1b. Legend

| Five-month visit | One-year visit | Total (N = 2,179) |
| --- | --- | --- |
| No | No | 232 (10·6 %) |
| Yes | Yes | 107 (4·9%) |
| Not sure | Not sure | 47 (2·2%) |
| No | Not sure | 64 (2·9%) |
| No | Yes | 30 (1·4%) |
| No | Not yet complete* | 753 (34·6%) |
| Not sure | No | 35 (1·6%) |
| Not sure | Yes | 31 (1·4%) |
| Not sure | Not yet complete* | 272 (12·5%) |
| Yes | No | 17 (0·8%) |
| Yes | Not sure | 27 (1·2%) |
| Yes | Not yet complete* | 350 (16·1%) |
| NA or missing** | No | 108 (5·0%) |
| NA or missing** | Yes | 64 (2·9%) |
| NA or missing** | Not sure | 42 (1·9%) |

*not yet complete = are not eligible for their one-year visit yet

**NA or missing = not applicable as did not have a five-month visit (one-year visit only) or missing where the question was not completed at the five-month visit.

**Figure S2. Clusters of mental, cognitive, and physical health impairments at five-months**

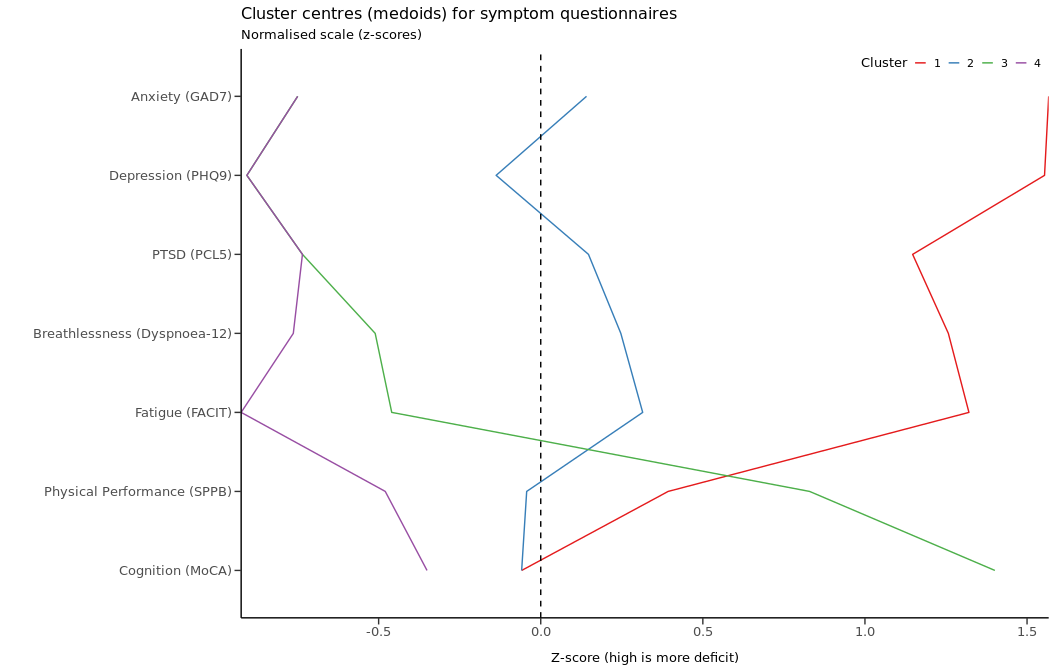

Figure S2 legend. Four recovery clusters by Z scores, where a higher Z score indicates a higher deficit. GAD-7 = Generalized Anxiety Disorder 7-item scale. PHQ-9 = Patient Health Questionnaire-9. PCL-5 Post Traumatic Stress Disorder Checklist. FACIT Fatigue Scale = Functional Assessment of Chronic Illness Therapy Fatigue Scale. SPPB=short physical performance battery. MoCA = Montreal Cognitive Assessment.

Cluster 1 Red - ‘very severe’ physical and mental health impairment

Cluster 2 Blue - ‘severe’ physical and mental health impairment

Cluster 3 Green - ‘moderate/cognitive’ physical impairment and cognitive impairment

Cluster 4 Purple - ‘mild’ physical and mental health impairment

**Figure S3. Characteristics associated with the four clusters**

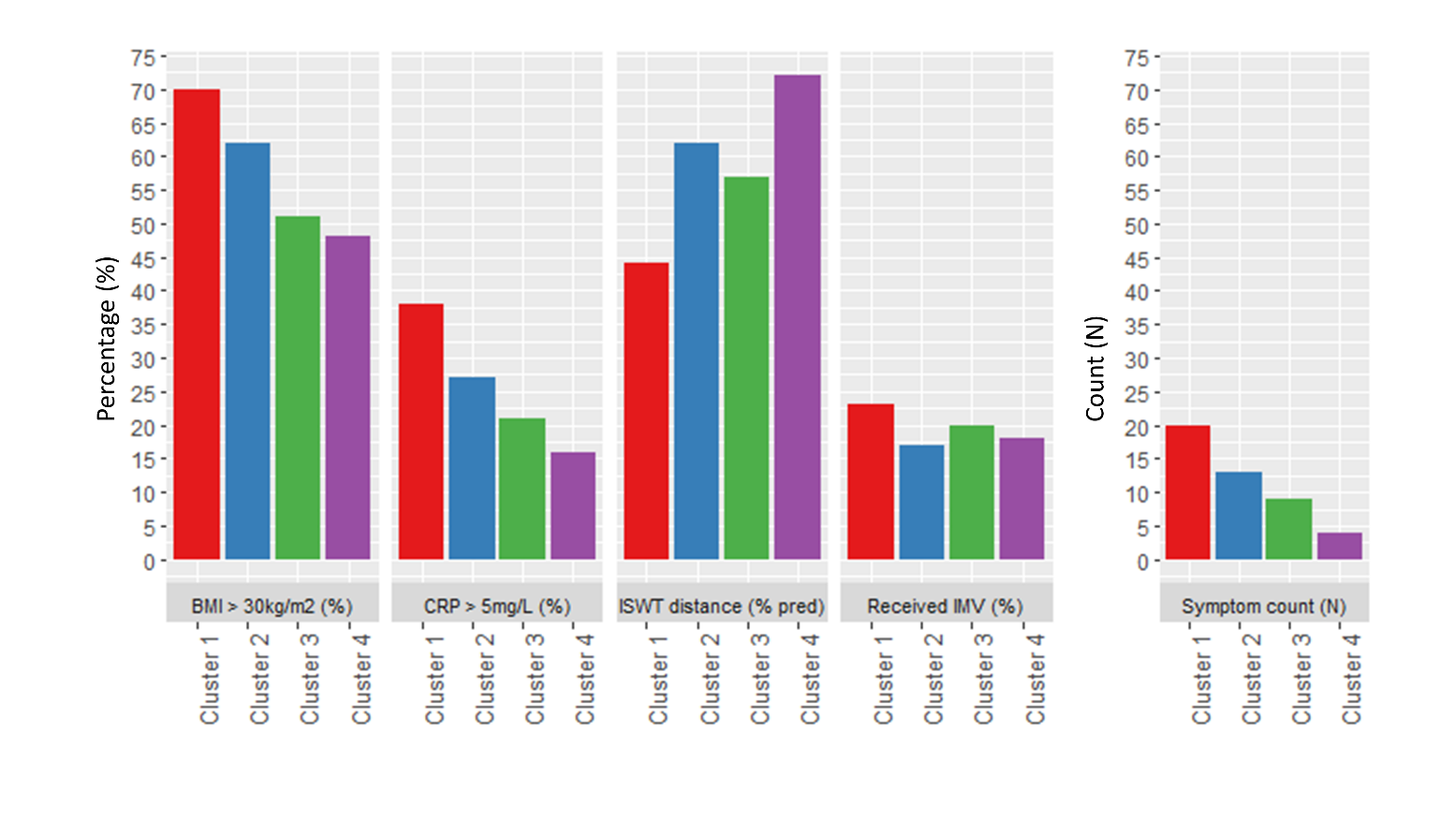

Cluster 1 Red - ‘very severe’ physical and mental health impairment

Cluster 2 Blue - ‘severe’ physical and mental health impairment

Cluster 3 Green - ‘moderate/cognitive’ physical impairment and cognitive impairment

Cluster 4 Purple - ‘mild’ physical and mental health impairment

BMI: Body Mass Index, CRP: C-reactive protein assessed at one year, ISWT: Incremental Shuttle Walk Test distance (metres) assessed at one year, IMV: invasive mechanical ventilation, symptom count at one year.

**Figure S4. Estimation plots for the features significantly upregulated when comparing cluster 1 (very severe) to cluster 4 (mild) (panels a to m) and when comparing cluster 3 (moderate/cognitive) to cluster 4 (mild) (panels n and o).**

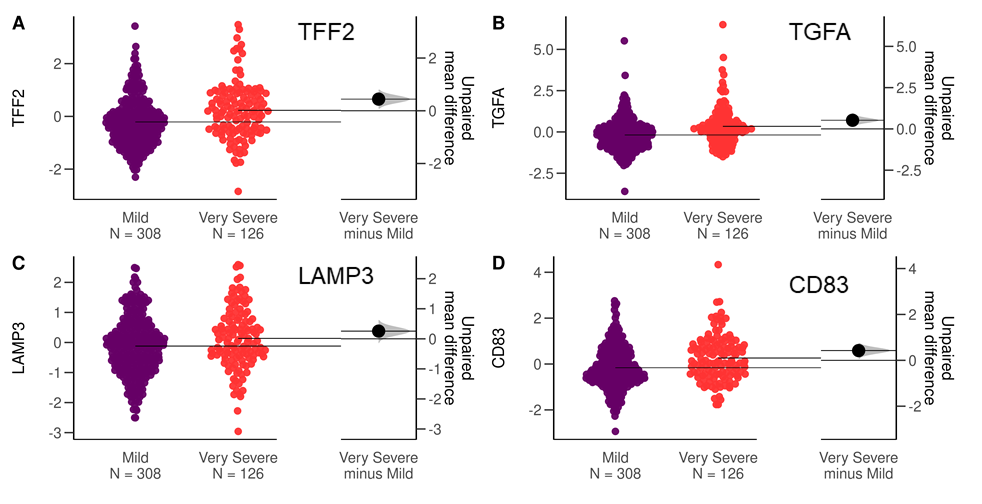

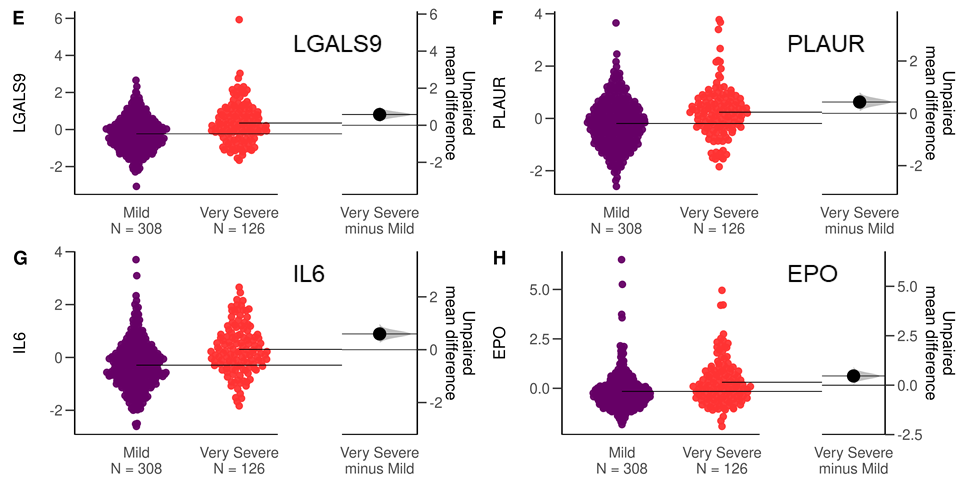

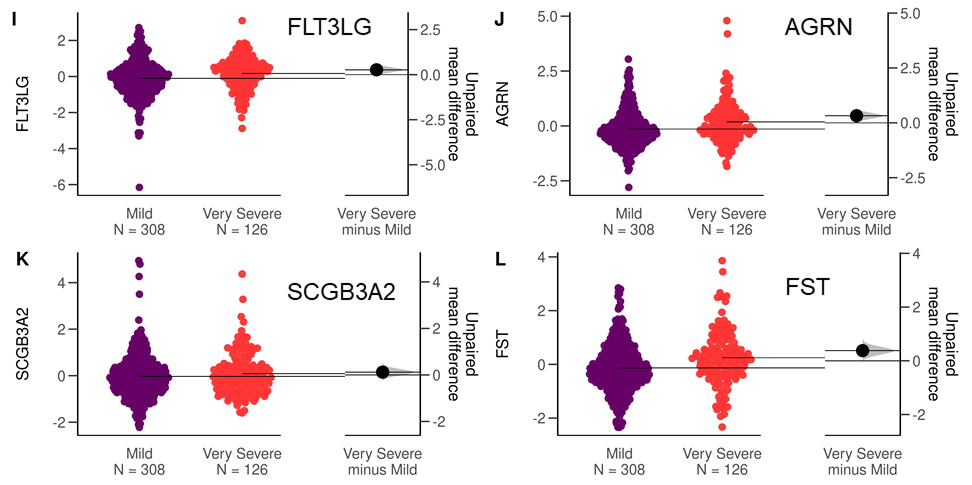

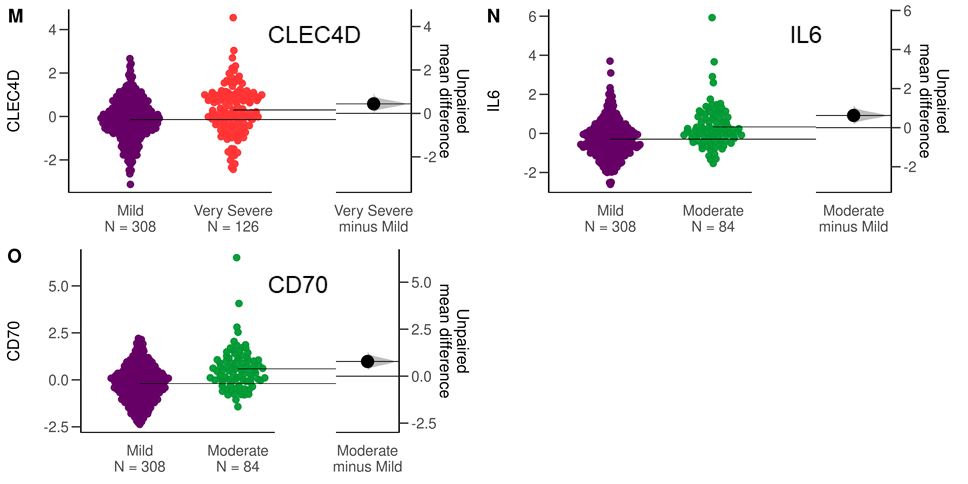

Figure S4. Legend: For each significantly associated feature, left-hand side shows distribution of normalised protein expression in cluster 4 (mild, purple, n=308) and cluster 1 (very severe, n=126, red, panels a to m) or cluster 3 (moderate/cognitive, green, n=84, panels n and o) with mean expression indicated by horizontal bars. Right hand side shows unpaired mean difference with confidence interval indicated by vertical bar. Plots include all participants with available measures for each feature, without exclusion of participants with missing clinical data for age, BMI and number of comorbidities).

**Figure S5. Health-related quality of life, disability, and symptoms across the four ‘severity’ clusters assessed for pre-hospitalisation (patient estimated), and at five-months and one-year post-discharge (individual complete data for all three time-points)**

a) Health-related quality of life assessed by the EQ5D-5L utility index

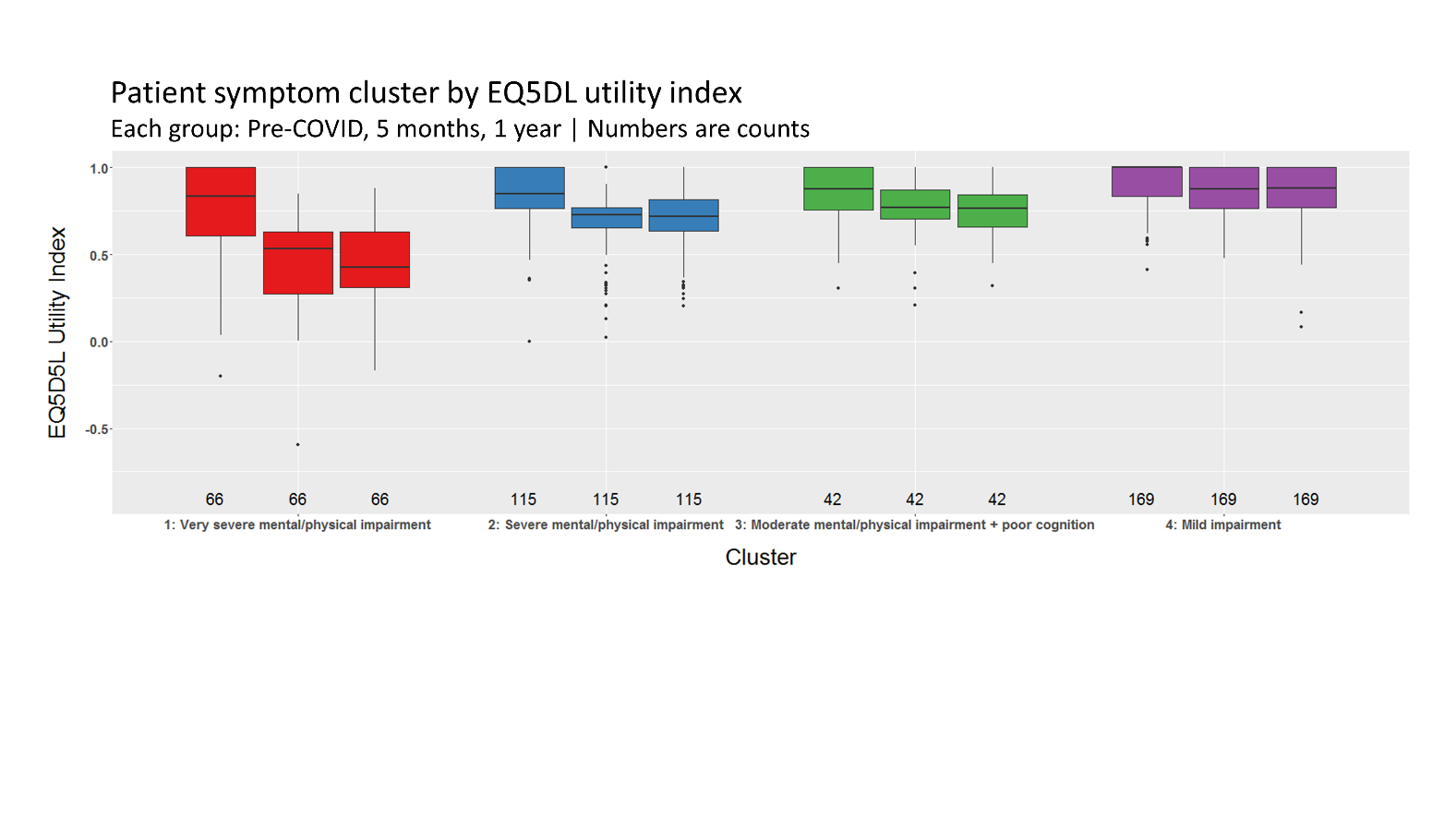

b) Disability assessed by the Washington Group Short Set of Functioning Severity Continuum (WG-SS-SCO) tool

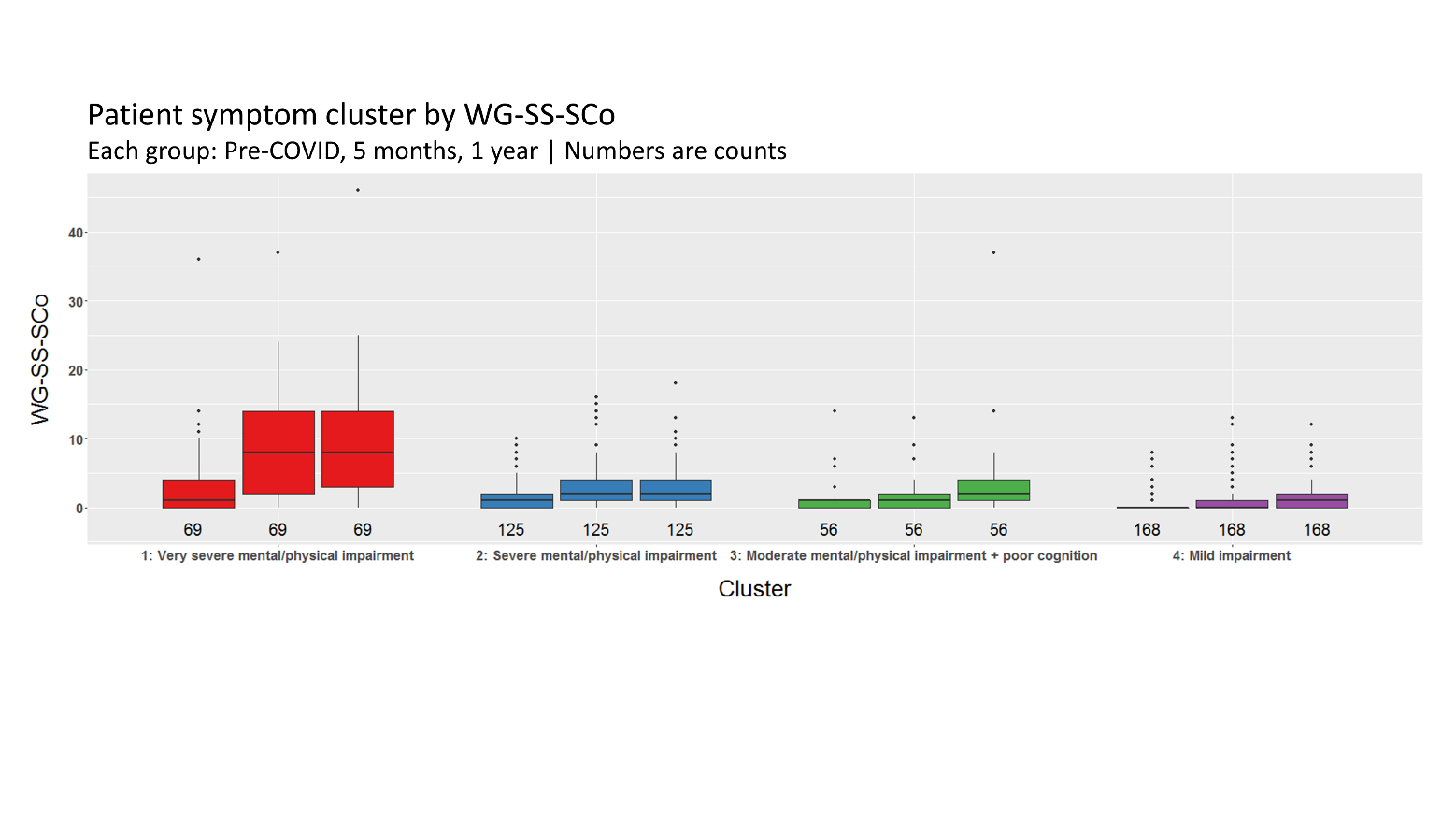

c) Fatigue severity in the last 24 hrs assessed using a Visual Analogue Scale 0-10

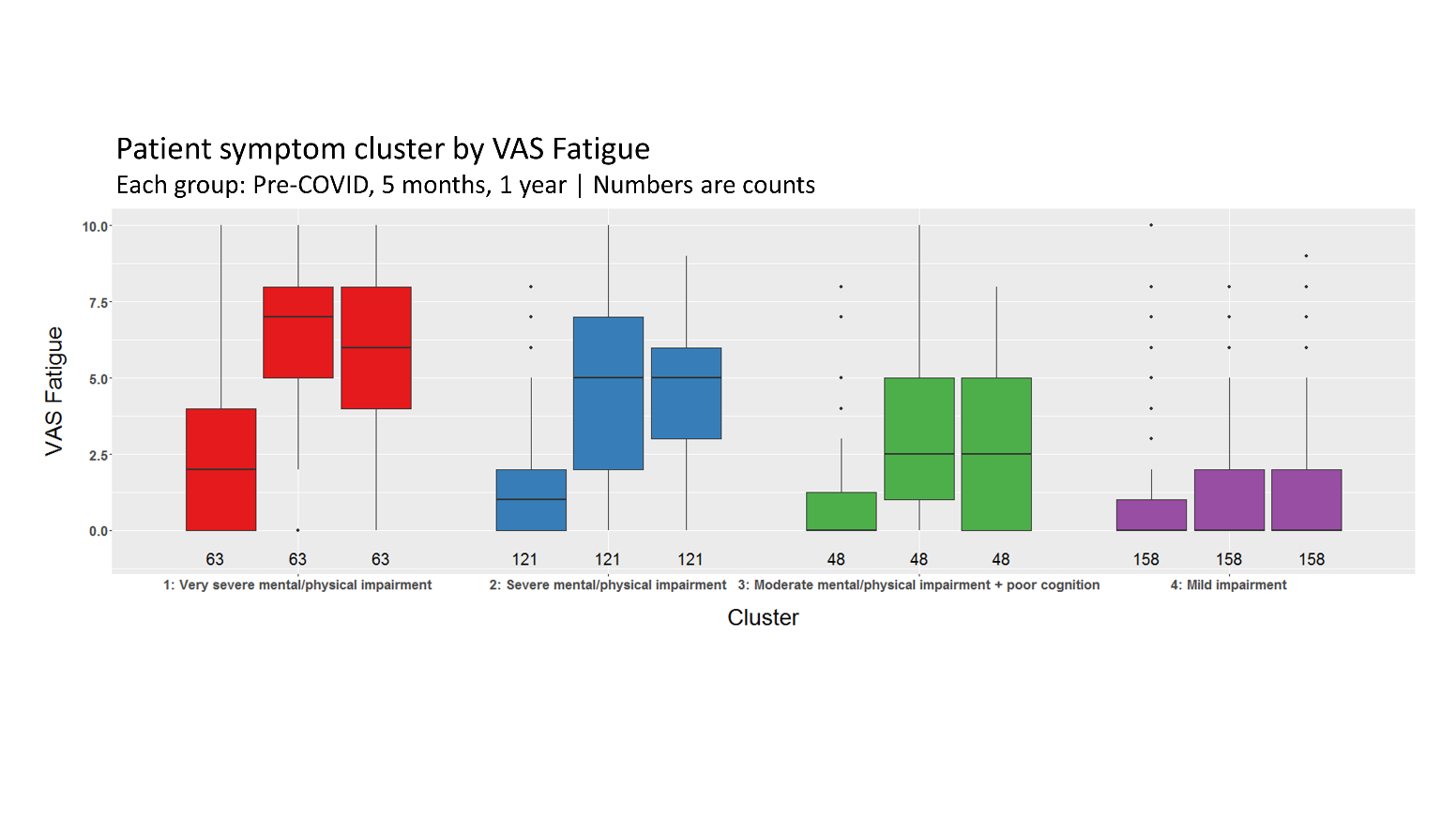

d) Breathlessness severity in the last 24 hrs assessed using a Visual Analogue Scale 0-10

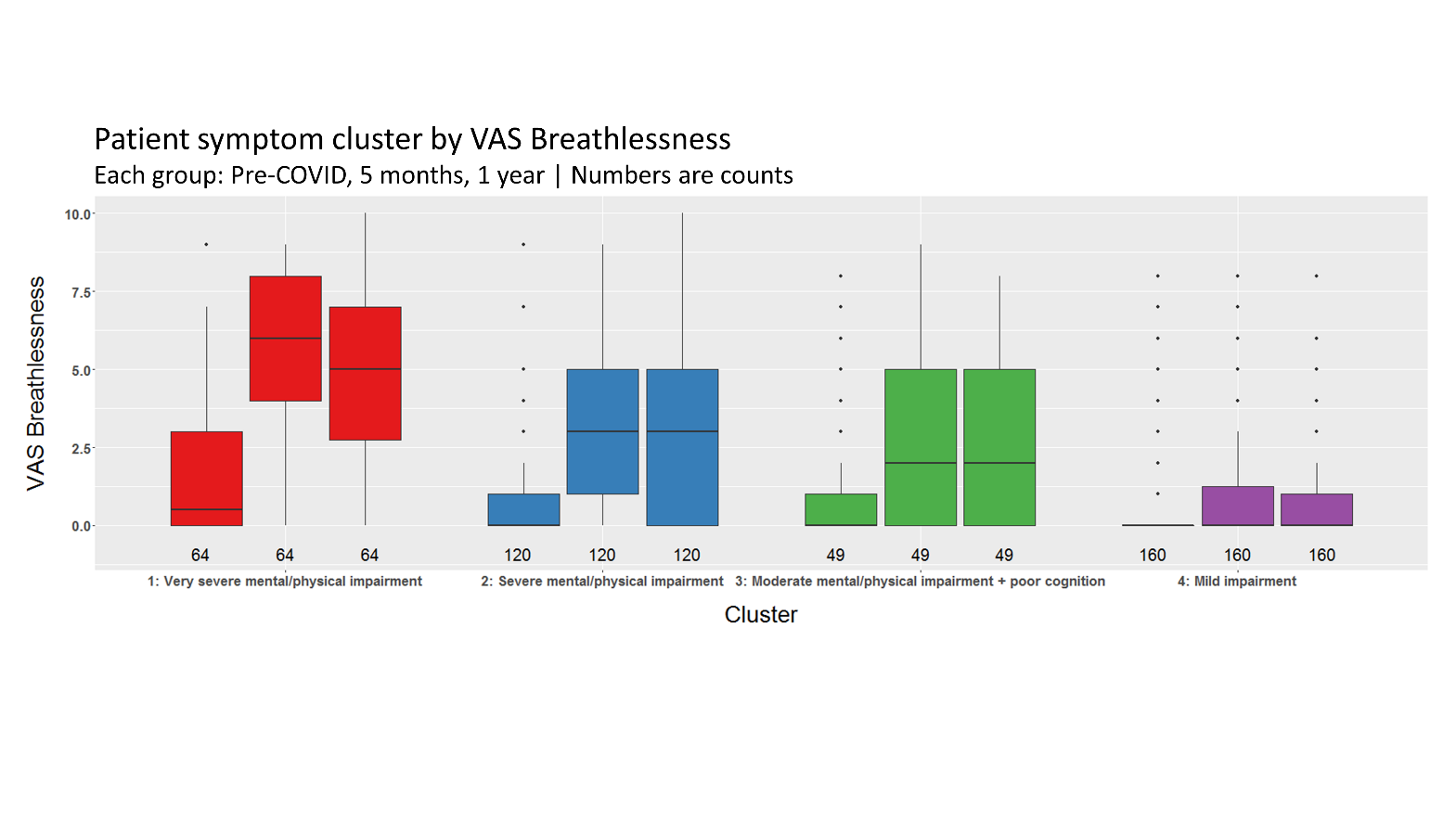

Figure S5. Legend

Cluster 1 Red - ‘very severe’ physical and mental health impairment

Cluster 2 Blue - ‘severe’ physical and mental health impairment

Cluster 3 Green - ‘moderate/cognitive’ physical impairment and cognitive impairment

Cluster 4 Purple - ‘mild’ physical and mental health impairment

Pre-hospital health status assessed retrospectively
