## Supplementary material for "Clinical characteristics with inflammation profiling of Long-COVID and association with one-year recovery following hospitalisation in the UK: a prospective observational study": PHOSP-COVID Collaborative Group

**Core Management Group**

*Chief Investigator* C E Brightling, *Members* R A Evans (Lead Co-I), L V Wain (Lead Co-I), J D Chalmers, V C Harris, L P Ho, A Horsley, M Marks, K Poinasamy, B Raman, A Shikotra, A Singapuri

**PHOSP-COVID Study Central Coordinating Team**

C E Brightling (Chief Investigator), R A Evans (*Lead Co-I*), L V Wain (*Lead Co-I*), R Dowling, C Edwardson, O Elneima, S Finney, B Hargadon, V Harris, L Houchen, O C Leavy, H McAuley, C Overton, T Plekhanova, R Saunders, M Sereno, A Singapuri, A Shikotra, C Taylor, S Terry, C Tong, B Zhao

**Steering Committee**

*Co-chairs* D Lomas, E Sapey*, Institution representatives* C Berry, C E Bolton, N Brunskill, E R Chilvers, R Djukanovic, Y Ellis, D Forton, N French, J George, N A Hanley, N Hart, L McGarvey, N Maskell, H McShane, M Parkes, D Peckham, P Pfeffer, A Sayer, A Sheikh, A A R Thompson, N Williams and core management group representation

**Executive Board**

*Chair* C E Brightling, representation from the core management group, each working group and platforms

**Platforms**

**Bioresource**

W Greenhalf (*Co-Lead*), M G Semple (*Co-Lead*), M Ashworth, H E Hardwick, M Sereno, R Saunders, A Singapuri, V Shaw, A Shikotra, L V Wain

**Data Hub**

A B Docherty (*Co-Lead*), E M Harrison (*Co-Lead*), A Sheikh (*Co-Lead*), J K Baillie, C E Brightling, L Daines, R A Evans, S Kerr, O C Leavy, N I Lone, H J C McAuley, R Pius, J Quint, M Richardson, M Sereno, M Thorpe, L V Wain

**Imaging Alliance**

M Halling-Brown (*Co-Lead*), F Gleeson (*Co-Lead*), J Jacob (*Co-Lead*), S Neubauer (*Co-Lead*) B Raman (*Co-Lead*) S Siddiqui (*Co-Lead*) J Wild (*Co-Lead*), S Aslani, P Jezzard, H Lamlum, E Tunnicliffe

**Omics**

L V Wain (*Co-Lead)*, J K Baillie (*Co-Lead*), H Baxendale, C E Brightling, M Brown, J D Chalmers, R A Evans, B Gooptu, W Greenhalf, H E Hardwick, R G Jenkins, D Jones, I Koychev, C Langenberg, A Lawrie, P L Molyneaux, A Shikotra, J Pearl, M Ralser, N Sattar, R Saunders, J Scott, T Shaw, D Thomas, D Wilkinson

**Working Groups**

**Airways**

L G Heaney (*Co-Lead*), A De Soyza (*Co-Lead*), D Adeloye, C E Brightling, J S Brown, J Busby, J D Chalmers, C Echevarria, L Daines, O Elneima, RA Evans, J Hurst, P Novotny, P Pfeffer, K Poinasamy, J Quint, I Rudan, E Sapey, A Sheikh, S Siddiqui, S Walker

**Brain**

J R Geddes (*Lead*), M Hotopf *(Co-Lead),* K Abel, R Ahmed, L Allan, C Armour, D Baguley, D Baldwin, C Ballard, J Bambrough, K Bhui, G Breen, M Broome, T Brugha, E Bullmore, D Burn, J Cavanagh, T Chalder, D Clark, A David, B Deakin, H Dobson, B Elliott, J Evans, R Francis, E Guthrie, P Harrison, M Henderson,  A Hosseini, N Huneke, M Husain, T Jackson, I Jones, T Kabir, P Kitterick, A Korszun, I Koychev, J Kwan, A Lingford-Hughes, C Mackay, P Mansoori, H McAllister-Williams, K McIvor, B Michael, L Milligan, R Morriss, E Mukaetova-Ladinska, T Nicholson, S Paddick, C Pariante, J Pimm, K Saunders, M Sharpe, G Simons, R Upthegrove, S Wessely

**Cardiac**

G P McCann (*Lead*), C Antoniades, R Bell, A Bularga, C Berry, K M Channon, P Chowienczyk, J P Greenwood, A Hingorani, A D Hughes, C Jolley, K Khunti, C Kotanidis, J Mayet, N L Mills, A J Moss, S Neubauer, D E Newby, B Raman, N Samani, A Saratzis, N Sattar, A M Shah, C L Sudlow, M Toshner, R Touyz, B Williams, C Xie

**Immunology**

P J M Openshaw (*Lead*), D Altmann, J K Baillie, R Batterham, H Baxendale, N Bishop, C E Brightling, P C Calder, R A Evans, J L Heeney, T Hussell, P Klenerman, J M Lord, P Moss, S L Rowland-Jones, W Schwaeble, M G Semple, R S Thwaites, L Turtle, L V Wain, S Walmsley, D Wraith

**Intensive Care**

M J Rowland (*Lead*), A Rostron (*Co-Lead*), J K Baillie, B Connolly, A B Docherty, N I Lone, D F McAuley, D Parekh, J Simpson, C Summers

**Lung Fibrosis**

R G Jenkins (*Co-Lead*), J Porter (*Co-Lead*), R J Allen, R Aul, J K Baillie, S Barratt, P Beirne, J Blaikley, R C Chambers, N Chaudhuri, C Coleman, L Fabbri, P M George, M Gibbons, F Gleeson, B Gooptu, I Hall, N A Hanley, L P Ho, E Hufton, J Jacob, S Johnson, M Jones, S Jones, F Khan, J Mitchell, P L Molyneaux, J E Pearl, K Piper Hanley, K Poinasamy, J Quint, P Rivera-Ortega, M G Semple, J Simpson, M Spears, L G Spencer, S Stanel, I Stewart, D Thickett, R Thwaites, L V Wain, S Walker, S Walsh, J Wild, D G Wootton, L Wright

**Metabolic**

S Heller (*Co-Lead*), M Davies (*Co-Lead*), H Atkins, S Bain, J Dennis, K Ismail, D Johnston, P Kar, K Khunti, C Langenberg, P McArdle, A McGovern, T Peto, J Petrie, E Robertson, N Sattar, K Shah, J Valabhji, B Young

**Pulmonary and Systematic Vasculature**

L Howard (*Co-Lead*), Mark Toshner (*Co-Lead*), C Berry, P Chowienczyk, D Lasserson, A Lawrie, O C Leavy, J Mitchell, L Price, J Quint, J Rossdale, N Sattar, C Sudlow, A A R Thompson, J M Wild, M Wilkins

**Rehabilitation, Sarcopenia and Fatigue**

S J Singh (*Co-Lead*), W D-C Man (*Co-Lead*), J M Lord (*Co-Lead*), N J Greening (*Co-Lead*), T Chalder (*Co-Lead*), J Scott (*Co-Lead*), N Armstrong, E Baldry, N Basu, M Beadsworth, L Bishop, C E Bolton, A Briggs, M Buch, G Carson, J Cavanagh, H Chinoy, E Daynes, S Defres, R A Evans, P Greenhaff, S Greenwood, M Harvie, M Husain, A McArdle, A McMahon, M McNarry, C Nolan, J Pimm, J Sargent, L Sigfrid, M Steiner, D Stensel, A L Tan, J Whitney, D Wilkinson, D Wilson, M Witham, D G Wootton, T Yates

**Renal**

D Thomas (*Lead*), N Brunskill (*Co-Lead*), S Francis (*Co-Lead*), S Greenwood (*Co-Lead*), C Laing (*Co-Lead*), K Bramham, P Chowdhury, A Frankel, L Lightstone, S McAdoo, K McCafferty, M Ostermann, N Selby, C Sharpe, M Willicombe

**Local Clinical Centre PHOSP-COVID trial staff**

(listed in alphabetical order)

**Airedale NHS Foundation Trust**

A Shaw (PI), L Armstrong, B Hairsine, H Henson, C Kurasz, L Shenton

**Aneurin Bevan University Health Board**

S Fairbairn (PI), A Dell, N Hawkings, J Haworth, M Hoare, A Lucey, V Lewis, G Mallison, H Nassa, C Pennington, A Price, C Price, A Storrie, G Willis, S Young

**Barts Health NHS Trust &** **Queen Mary University of London**

P Pfeffer (PI), K Chong-James, C David, W Y James, A Martineau, O Zongo

**Barnsley Hospital NHS Foundation Trust**

A Sanderson (PI)

**Belfast Health and Social Care Trust & Queen's University Belfast**

L G Heaney (PI), C Armour, V Brown, T Craig, S Drain, B King, N Magee, D McAulay, E Major, L McGarvey, J McGinness, R Stone

**Betsi Cadwaladr University Health Board**

A Haggar (PI), A Bolger, F Davies, J Lewis, A Lloyd, R Manley, E McIvor, D Menzies, K Roberts, W Saxon, D Southern, C Subbe, V Whitehead

**Borders General Hospital, NHS Borders**

H El-Taweel (PI), J Dawson, L Robinson

**Bradford Teaching Hospitals NHS Foundation Trust**

D Saralaya (PI), L Brear, K Regan, K Storton

**Cambridge University Hospitals NHS Foundation Trust, NIHR Cambridge Clinical Research Facility & University of Cambridge**

J Fuld (PI), A Bermperi, I Cruz, K Dempsey, A Elmer, H Jones, S Jose, S Marciniak, M Parkes, C Ribeiro, J Taylor, M Toshner, L Watson, J Worsley

**Cardiff and Vale University Health Board**

R Sabit (PI), L Broad, A Buttress, T Evans, M Haynes, L Jones, L Knibbs, A McQueen, C Oliver, K Paradowski, J Williams

**Chesterfield Royal Hospital NHS Trust**

E Harris (PI), C Sampson

**Cwm Taf Morgannwg University Health Board**

C Lynch (PI), E Davies, C Evenden , A Hancock, K Hancock, M Rees , L Roche, N Stroud, T Thomas-Woods

**East Cheshire NHS Trust**

M Babores (PI), J Bradley-Potts, M Holland, N Keenan, S Shashaa, H Wassall

**East Kent Hospitals University NHS Foundation Trust**

E Beranova (PI), H Weston (PI), T Cosier, L Austin, J Deery, T Hazelton, C Price, H Ramos, R Solly, S Turney

**Gateshead NHS Trust**

L Pearce (PI), W McCormack, S Pugmire, W Stoker, A Wilson

**Guy’s and St Thomas’ NHS Foundation Trust**

N Hart (PI), LA Aguilar Jimenez, G Arbane, S Betts, K Bisnauthsing, A Dewar, P Chowdhury, A Dewar, G Kaltsakas, H Kerslake, MM Magtoto, P Marino, LM Martinez, M Ostermann, J Rossdale, TS Solano, E Wynn

**Hampshire Hospitals NHS Foundation Trust**

N Williams (PI), W Storrar (PI), M Alvarez Corral, A Arias, E Bevan, D Griffin, J Martin, J Owen,

S Payne, A Prabhu, A Reed, C Wrey Brown

**Harrogate and District NHD Foundation Trust**

C Lawson (PI), T Burdett, J Featherstone, A Layton, C Mills, L Stephenson,

**Hull University Teaching Hospitals NHS Trust & University of Hull**

N Easom (PI), P Atkin, K Brindle, M G Crooks, K Drury, R Flockton, L Holdsworth, A Richards, D L Sykes, S Thackray-Nocera, C Wright

**Hywel Dda University Health Board**

K E Lewis (PI), A Mohamed (PI), G Ross (PI), K Davies, R Hughes, R Loosley, L O’Brien, Z Omar, H McGuinness, E Perkins, J Phipps, A Taylor, H Tench, R Wolf-Roberts

**Imperial College Healthcare NHS Trust & Imperial College London**

L Howard (PI), O Kon (PI), D C Thomas (PI), S Anifowose, L Burden, E Calvelo, B Card, C Carr, E R Chilvers, D Copeland, P Cullinan, P Daly, L Evison, T Fayzan, H Gordon, S Haq, R G Jenkins, C King, K March, M Mariveles, L McLeavey, N Mohamed, S Moriera, U Munawar, J Nunag, U Nwanguma, L Orriss- Dib, A Ross, M Roy, E Russell, K Samuel, J Schronce, N Simpson, L Tarusan, C Wood, N Yasmin

**Kettering General Hospital NHS Trust**

R Reddy (PI), A-M, Guerdette, M Hewitt, K Warwick, S White

**King’s College Hospital NHS Foundation Trust & Kings College London**

A M Shah (PI), C J Jolley (PI), O Adeyemi, R Adrego, H Assefa-Kebede, J Breeze, M Brown, S Byrne, T Chalder, P Dulawan, N Hart, A Hayday, A Hoare, A Knighton, M Malim, S Patale, I Peralta, N Powell, A Ramos, K Shevket, F Speranza, A Te

**Leeds Teaching Hospitals & University of Leeds**

P Beirne (PI), A Ashworth, J Clarke, C Coupland, M Dalton, E Wade, C Favager, J Greenwood, J Glossop, L Hall, T Hardy, A Humphries, J Murira, D Peckham, S Plein, J Rangeley, G Saalmink, A L Tan, B Whittam, N Window, J Woods,

**Lewisham & Greenwich NHS Trust**

G Coakley (PI)

**Liverpool University Hospitals NHS Foundation Trust & University of Liverpool**

D G Wootton (PI), L Turtle (PI), L Allerton, AM All, M Beadsworth, A Berridge, J Brown, S Cooper, A Cross, S Defres, S L Dobson, J Earley, N French, W Greenhalf, H E Hardwick, K Hainey, J Hawkes, V Highett, S Kaprowska, AL Key, L Lavelle-Langham, N Lewis-Burke, G Madzamba, F Malein, S Marsh, C Mears, L Melling, M J Noonan, L Poll, J Pratt, E Richardson, A Rowe, M G Semple, V Shaw, K A Tripp, L O Wajero, S A Williams-Howard, J Wyles,

**London North West University Healthcare NHS Trust**

S N Diwanji (PI), P Papineni (PI), S Gurram, S Quaid, G F Tiongson, E Watson

**Manchester University NHS Foundation Trust & University of Manchester**

B Al-Sheklly (PI), A Horsley (PI), C Avram, J Blaikely, M Buch, N Choudhury, D Faluyi, T Felton, T Gorsuch, N A Hanley, T Hussell, Z Kausar, N Odell, R Osbourne, K Piper Hanley, K Radhakrishnan, S Stockdale

**Newcastle upon Tyne Hospitals NHS Foundation Trust & University of Newcastle**

A De Soyza (PI), C Echevarria (PI), A Ayoub, J Brown, G Burns, G Davies, H Fisher, C Francis, A Greenhalgh, P Hogarth, J Hughes, K Jiwa, G Jones, G MacGowan, D Price, A Sayer, J Simpson, H Tedd, S Thomas, S West, M Witham, S Wright, A Young

**NHS Dumfries and Galloway**

M J McMahon (PI), P Neill

**NHS Greater Glasgow and Clyde Health Board & University of Glasgow**

D Anderson (PI), H Bayes (PI), C Berry (PI), D Grieve (PI), I B McInnes (PI), N Basu, A Brown, A Dougherty, K Fallon, L Gilmour, K Mangion, A Morrow, K Scott, R Sykes

**NHS Highland**

E K Sage (PI), F Barrett, A Donaldson

**NHS Lanarkshire**

M Patel (PI), D Bell, A Brown, M Brown, R Hamil, K Leitch, L Macliver, J Quigley, A Smith, B Welsh

**NHS Lothian & University of Edinburgh**

G Choudhury (PI), J K Baillie, S Clohisey, A Deans, A B Docherty, J Furniss, E M Harrison, S Kelly, N I Lone, A Sheikh

**NHS Tayside & University of Dundee**

J D Chalmers (PI), D Connell, A Elliott, C Deas, J George, S Mohammed, J Rowland, A R Solstice, D Sutherland, C J Tee

**North Bristol NHS Trust & University of Bristol**

N Maskell (PI), D Arnold, S Barrett, H Adamali, A Dipper, S Dunn, A Morley, L Morrison, L Stadon, S Waterson, H Welch

**North Middlesex Hospital NHS Trust**

B Jayaraman (PI), T Light

**Nottingham University Hospitals NHS Trust & University of Nottingham**

C E Bolton (PI), P Almeida, J Bonnington, M Chrystal, P Greenhaff, A Gupta, L Howard, S Linford, L Matthews, R Needham, K Shaw, A K Thomas

**Oxford University Hospitals NHS Foundation Trust & University of Oxford**

L P Ho (PI), N M Rahman (PI), M Ainsworth, A Alamoudi, A Bloss, P Carter, J Chen, F Conneh, T Dong, R I Evans, E Fraser, J R Geddes, F Gleeson, P Harrison, M Havinden-Williams, P Jezzard, I Koychev, P Kurupati, H McShane, C Megson, S Neubauer, D Nicoll, G Ogg, E Pacpaco, M Pavlides, Y Peng, N Petousi, N Rahman, B Raman, M J Rowland, K Saunders, M Sharpe, N Talbot, E Tunnicliffe

**Royal Brompton and Harefield Clinical Group, Guy’s and St Thomas’ NHS Foundation Trust.**

W Man (PI), B Patel (PI), R E Barker, D Cristiano, N Dormand, M Gummadi, S Kon, K Liyanage, C M Nolan, S Patel, O Polgar, P Shah, S J Singh, J A Walsh

**Royal Free London NHS Foundation Trust**

J Hurst (PI), H Jarvis (PI), S Mandal (PI), S Ahmad, S Brill, L Lim, D Matila, O Olaosebikan, C Singh

**Royal Papworth Hospital NHS Foundation Trust**

M Toshner (PI), H Baxendale, L Garner, C Johnson, J Mackie, A Michael, J Pack, K Paques, H Parfrey, J Parmar

**Salford Royal NHS Foundation Trust**

N Diar Bakerly (PI), P Dark, D Evans, E Hardy, A Harvey, D Holgate, S Knight, N Mairs, N Majeed, L McMorrow, J Oxton, J Pendlebury, C Summersgill, R Ugwuoke, S Whittaker

**Salisbury NHS Foundation Trust**

W Matimba-Mupaya (PI), S Strong-Sheldrake

**Sheffield Teaching NHS Foundation Trust & University of Sheffield**

S Rowland-Jones (PI), A A R Thompson (Co PI), A Alli, A Angyal, L Armstrong, M Bayley, E Bradley, R Brown, K Chapman, L Chetham, C Clark, Z Coburn, J Cole, T De Silva, M Dixon, A Fairman, D Foote, A Ford, Z Gabriel, R Gregory, K Harrington, S Heller, L Hesselden, A Holbourn, B Holroyd-Hind, A Howell, F Ilyas, C Jarman, A Lawrie, E Lee, R Lenagh, A Lye, I Macharia, S Megson, J Meiring, H Newell, T Newman, C Norman, L Nwafor, M Plowright, J Porter, J Rodgers, I Smith, J Smith, L Smith, R Stimpston, A A R Thompson, H Turton, H Turton, P Wade, J Watson, L Watson, J Wild, I Wilson, A Zawia

**St George’s University Hospitals NHS Foundation Trust**

R Aul (PI), M Ali, A Dunleavy (PI), D Forton, N Msimanga, M Mencias, T Samakomva, S Siddique, J Teixeira, V Tavoukjian

**Sherwood Forest Hospitals NHS Foundation Trust**

J Hutchinson (PI), L Allsop, K Bennett, P Buckley, M Flynn, M Gill, C Goodwin, M Greatorex, H Gregory, C Heeley, L Holloway, M Holmes, J Kirk, W Lovegrove, TA Sewell, S Shelton, D Sissons, K Slack, S Smith, D Sowter, S Turner, V Whitworth, I Wynter

**Shropshire Community Health NHS Trust**

L Warburton (PI), S Painter, J Tomlinson

**Somerset NHS Foundation Trust**

C Vickers (PI), T Wainwright, D Redwood, J Tilley, S Palmer

**Swansea Bay University Health Board**

G A Davies (PI), L Connor, A Cook, T Rees, F Thaivalappil, C Thomas

**Tameside and Glossop Integrated Care NHS Foundation**

A Butt (PI), M Coulding, H Jones, S Kilroy, J McCormick, J McIntosh, H Savill, V Turner, J Vere

**The Great Western Hospital Foundation Trust**

E Fraile (PI), J Ugoji

**The Hillingdon Hospitals NHS Foundation Trust**

S S Kon (PI), H Lota, G Landers, M Nasseri, S Portukhay

**The Rotherham NHS Foundation Trust**

A Hormis (PI), A Daniels, J Ingham, L Zeidan

**United Lincolnshire Hospitals NHS Trust**

M Chablani (PI), L Osborne

**University College London Hospital & University College London**

M Marks (PI), J S Brown (PI), N Ahwireng, B Bang, D Basire, R C Chambers, A Checkley, R Evans, M Heightman, T Hillman, J Hurst, J Jacob, S Janes, R Jastrub, M Lipman, S Logan, D Lomas, M Merida Morillas, H Plant, J C Porter, K Roy, E Wall

**University Hospital Birmingham NHS Foundation Trust & University of Birmingham**

D Parekh (PI), N Ahmad Haider, C Atkin, R Baggott, M Bates, A Botkai, A Casey, B Cooper, J Dasgin, K Draxlbauer, N Gautam, J Hazeldine, T Hiwot, S Holden, K Isaacs, T Jackson, S Johnson, V Kamwa, D Lewis,

J M Lord, S Madathil, C McGhee, K Mcgee, A Neal, A Newton Cox, J Nyaboko, D Parekh, Z Peterkin, H Qureshi, L Ratcliffe, E Sapey, J Short, T Soulsby, J Stockley, Z Suleiman, T Thompson, M Ventura, S Walder, C Welch, D Wilson, S Yasmin, K P Yip

**University Hospitals of Derby and Burton**

P Beckett (PI) C Dickens, U Nanda

**University Hospitals of Leicester NHS Trust & University of Leicester**

C E Brightling (CI), R A Evans (PI), M Aljaroof, N Armstrong, H Arnold, H Aung, M Bakali, M Bakau, M Bourne, C Bourne, N Brunskill, P Cairns, L Carr, A Charalambou, C Christie, M Davies, S Diver, S Edwards, C Edwardson, O Elneima, H Evans, J Finch, S Glover, N Goodman, B Gootpu, N J Greening, K Hadley, P Haldar, B Hargadon, V C Harris, L Houchen, W Ibrahim, L Ingram, K Khunti, A Lea, D Lee, G P McCann, H J C McAuley, P McCourt, T Mcnally, A Moss, W Monteiro, M Pareek, S Parker, A Rowland, A Prickett, I N Qureshi, R Russell, M Sereno, A Shikotra, S Siddiqui, A Singapuri, S J Singh, J Skeemer, M Soares, E Stringer, T Thornton, M Tobin, L V Wain, T J C Ward, F Woodhead, T Yates, A Yousuf

**University Hospital Southampton NHS Foundation Trust & University of Southampton**

M Jones (PI), C Childs, R Djukanovic, S Fletcher, M Harvey, E Marouzet, B Marshall, R Samuel, T Sass, T Wallis, H Wheeler

**Whittington Health NHS**

R Dharmagunawardena (PI), E Bright, P Crisp, M Stern

**Wirral University Teaching Hospital**

A Wight (PI), L Bailey, A Reddington

**Wrightington Wigan and Leigh NHS trust**

A Ashish (PI), J Cooper, E Robinson

**Yeovil District Hospital NHS Foundation Trust**

A Broadley (PI)

**York & Scarborough NHS Foundation Trust**

K Howard (PI), L Barman, C Brookes, K Elliott. L Griffiths, Z Guy, D Ionita, H Redfearn, C Sarginson

A Turnbull

**Health and Care Research Wales**

Y Ellis

**London School of Hygiene & Tropical Medicine (LSHTM)**

M Marks, A Briggs

**NIHR Office for Clinical Research Infrastructure**

K Holmes

**Patient Public Involvement Leads**

Asthma UK and British Lung Foundation Partnership - K Poinasamy, S Walker

**Royal Surrey NHS Foundation Trust**

M Halling-Brown

**South London and Maudsley NHS Foundation Trust & Kings College London**

G Breen, M Hotopf

**Swansea University & Swansea Welsh Network**

K Lewis, N Williams
